# 24S-Hydroxycholesterol: A potential brain-derived biomarker of Huntington’s Disease

**DOI:** 10.1101/2025.10.08.25337337

**Authors:** Mohsen Ali Asgari, Douglas R Langbehn, David O F Skibinski, Ramee Lee, William J Griffiths, Yuqin Wang

## Abstract

Huntington’s disease (HD) arises from of an expanded polyglutamine-coding trinucleotide repeat in exon 1 of the huntingtin gene. Despite considerable research, a successful disease modifying therapy has yet to be confirmed. Cholesterol homeostasis in brain is considered one of the biological processes to be disturbed in HD.

In the current study we have shown, using liquid chromatography – mass spectrometry with multistage fragmentation – based sterolomics, that non-esterified 24S-hydroxycholesterol (24S-HC), the stereo-specific brain-derived cholesterol metabolite that can cross the blood brain barrier, is at a lower concentration in plasma of a group of people with manifest HD than in groups of people with premanifest HD or healthy controls.

Although our data does not indicate that 24S-HC is a prognostic biomarker at the individual level, it could potentially be used to monitor a pharmacodynamic response for groups of patients, or perhaps of disease progression in individuals. In addition, by exploiting binary classification techniques we show that a diagnostic model can be developed giving a good to very good performance according to the area under the curve (AUC) in receiver operating characteristic (ROC) curves to distinguish people with premanifest HD from people with manifest HD. Notably, the most important metabolites in the binary classification models were plasma concentrations of 24S-HC and of the cholesterol precursor 7-dehydrocholesterol, both of which showed statistical changes in the manifest HD group, reinforcing the involvement of cholesterol metabolism in HD.

## Introduction

Huntington’s disease (HD) results from of an expanded polyglutamine-coding trinucleotide (CAG) repeat in exon 1 of the huntingtin gene (*HTT*).^1^ HD is an autosomal dominant disease, and when the CAG-repeat length is greater than 39, the result is almost complete disease penetrance; inheritance of 36-39 CAG repeats is partially penetrant. Longer repeat lengths are associated with earlier disease onset.^2^ The wild type HTT protein is expressed in many tissues (https://www.proteinatlas.org/ENSG00000197386-HTT/tissue),^3^ and appears to be involved in several brain functions.^4^ In HD the polyglutamine expansion at the *N*-terminal region of the protein is thought to confer both toxic gain-of-function and loss-of-function effects that may contribute to neuron dysfunction and death.^2^ HD presents with symptoms of progressive motor, cognitive and behavioural abnormalities.^2^ Onset of symptoms is typically at ages between 35 and 40 years and disease duration is about 15 to 20 years after onset of motor dysfunction.^5^ Medium spiny neurons of the striatum are the most affected tissue in HD. Despite considerable knowledge of the disease, a successful disease-modifying therapy has yet to be confirmed.

Dysfunctional cholesterol homeostasis in brain is linked to HD.^6^ Several studies in HD mouse models have indicated reduced cholesterol synthesis in brain, but the results are on occasion contradictory,^7–10^ leading to two different ideas for HD therapeutics; one based on enhancing cholesterol levels in brain,^6,9,11,12^ the other on enhancing its metabolism to 24S-hydroxycholesterol (24S-HC).^13,14^ Since cholesterol cannot cross the blood brain barrier (BBB), essentially all cholesterol in brain is by necessity synthesised *de novo* in brain,^15^ and the majority of any excess cholesterol is removed by metabolism to 24S-HC that can cross the BBB and enter the circulation.^16^ 24S-HC is formed from cholesterol in a reaction catalysed by the enzyme cholesterol 24-hydroxylase (CYP46A1) that is mostly expressed in neurons,^17^ and in human essentially all 24S-HC found in the circulation is derived cerebrally, although this is not necessarily the case in rodents.^18^

Since circulatory 24S-HC in humans is derived from brain, it has been suggested to be a marker of brain cholesterol homeostasis and especially of the number of metabolically active neurons.^10,19,20^ However, a complication of using 24S-HC as a potential biomarker for brain health is that its levels change with age,^16^ and this may explain some of the inconsistent results in some studies of neurodegeneration.^21^

Although metabolism to 24S-HC represents the major route for cholesterol removal from brain, other cholesterol metabolites can cross the BBB and via diffusion, be exported from, or imported to, brain and could also represent markers of brain health.^22–24^ While export into plasma represents the dominant elimination route, a small proportion of 24S-HC is also removed via the CSF,^25^ hence this route could also offer information on cholesterol homeostasis and neuronal health.

There is a lack of reliable biochemical biomarkers of HD. A good biomarker can track disease progression and/or a pharmacological response, and, ideally, provide a surrogate endpoint in clinical trials. At present, the measurement of neurofilament light chain (NfL) in CSF and plasma appears promising, with increased levels of NfL found in CSF and plasma from people with HD (PwHD) compared to controls.^26^ Similarly, levels of mutant HTT protein are elevated in the plasma from PwHD compared to controls ^27^. However, the measurement of 24S-HC and other related molecules in plasma or CSF may also provide a potential biomarker.^28–30^

In the current study we have measured, using liquid chromatography – mass spectrometry with multistage fragmentation (LC-MS(MS^n^)), the concentration of 25 different oxidised cholesterol metabolites (oxysterols), cholesterol itself and four precursors in plasma from 129 samples from PwHD prior to presentation of any signs/symptoms (premanifest HD), 188 samples from PwHD who are presenting signs/symptoms (manifest HD) and 83 samples from a control cohort. From the same donor cohort, we also quantified 11 cholesterol metabolites, cholesterol and five cholesterol precursors in CSF. We tested the primary hypothesis that concentrations of 24S-HC and of cholesterol are different between the three clinical groups. When controlling for age, sex and the use of statins and treating premanifest HD and manifest HD as categorical variables, the concentration of 24S-HC in plasma was found to be significantly reduced in manifest HD compared to premanifest and control groups, although there is evidence for an age by severity interaction. There were no differences in the concentration of 24S-HC between the control and premanifest groups nor in the concentration of cholesterol between the three groups. With respect to 24S-HC and cholesterol in CSF there was no significant group differences after controlling for age, sex and statin use. Considering the cholesterol precursors and similarly controlling for age and sex and statin use, the concentrations of 7-dehydrocholesterol (7-DHC) and 8-dehydrocholesterol (8-DHC) were found to be elevated in both plasma and CSF from the manifest HD group compared to premanifest HD or controls suggesting a greater importance of the Kandutsch-Russel pathway of cholesterol biosynthesis in manifest HD compared to the premanifest and control groups or alternatively reduced activity in 7-dehydrocholesterol reductase, the final enzyme in this pathway in people with manifest HD. These results were not changed when controlling also for the antipsychotic medications haloperidol and aripiprazole, known to affect plasma levels of cholesterol precursors.^31^ Finally, we utilised several binary classification techniques, taking the different sterols measured as variables, to classify individuals into one of two alternative groups. The classification of premanifest HD and manifest HD participants was most successful with area under the receiver operating characteristic (ROC) curve, (AUC), giving good to very good or excellent for seven classification methods tested.

## Materials and methods

### Plasma and CSF

Plasma and CSF samples were from the HDClarity cohort, an ongoing international biofluid collection initiative under the Enroll-Hd platform.^32^ The HDClarity study protocol is open-access and available at http://hdclarity.net/study-information/. HDClarity is conducted in accordance with the Declaration of Helsinki; all patients provide written informed consent for participation for future analyses based on their data. The HDClarity study was approved centrally by Camberwell St Giles Research Ethics Committee (IRAS 185506) and each collection site’s ethics board. Blood and CSF were collected after overnight fast. Participants were healthy controls and PwHD across the disease spectrum. PwHD were classified into categories using the following criteria: diagnostic confidence, CAG-repeat length, disease burden score (DBS) and total functional capacity score (TFC). The categories were:

(1) <u>Early premanifest</u>; diagnostic confidence < 4 and CAG ≥ 40 and DBS < 250. (2) <u>Late</u> <u>premanifest</u>; diagnostic confidence < 4 and CAG ≥ 40 and DBS ≥ 250. (3) <u>Early Manifest</u>; diagnostic confidence = 4 and CAG ≥ 36 and TFC score of 7 to 13. (4) <u>Moderate Manifest HD</u>: diagnostic confidence = 4 and CAG ≥ 36 and TFC score of 3 to 6. (5) <u>Advanced Manifest HD</u>: diagnostic confidence = 4 and CAG ≥ 36 and TFC score of 0 to 2.

Heathy controls represented a 6^th^ group. Additional information can be found in the HDClarity Data Dictionary https://enroll-hd.org/enrollhd_documents/HDClarity/2021-04-R2/HDClarity-DD_HDClarity-2021-04-R2_2021-06-24.pdf. Demographic information is listed in Table 1.

**Table 1.**
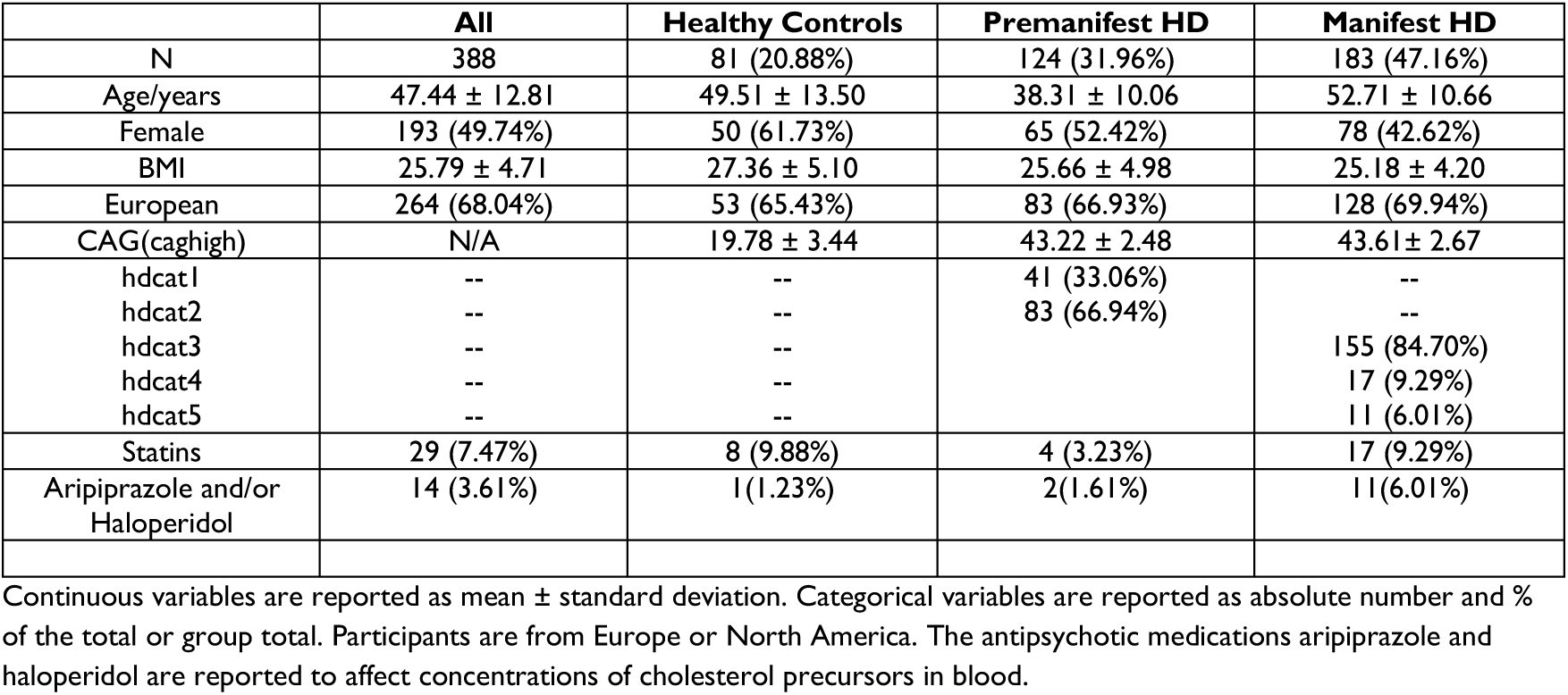
Characteristic of participants who donated samples.

### Sample preparation

Oxysterols and cholesterol precursors tend to give low-intensity signals upon LC-MS analysis.^10,33^ To overcome this problem, we have adopted a derivatisation protocol to maximise signal.^34–36^ The sample preparation protocol is described in full in Yutuc *et al*.^36^ and given in brief below.

#### Plasma

For analysis of oxysterol in plasma no hydrolysis step was performed as it is the non-esterified molecules that cross the BBB.^20^ In brief, plasma (100 µL) was added dropwise to a solution of 1.050 mL of absolute ethanol containing deuterated sterol and oxysterol standards (see Supplementary Table S1) and diluted by addition of 350 µL of water to give a solution of 70% ethanol. The solution was centrifuged at 14,000 x g at 4°C for 30 min to remove any precipitated matter. The solution (1.5 mL, 70% ethanol) was applied to a Sep-Pack tC_18_ 200 mg (Waters Corp) reversed-phase solid phase extraction column (SPE1) and the eluate combined with a column wash of 5.5 mL of 70% ethanol to give SPE1-Fr1. This fraction is devoid of cholesterol thereby avoiding the problem of *ex vivo* cholesterol autoxidation during sample preparation. After a wash with 4 mL of 70% ethanol (SPE1-FR2), cholesterol and similarly lipophilic sterols were eluted in 2 mL of absolute ethanol (SPE1-Fr3).

SPE1-Fr1 was then divided into two equal volumes, SPE1-Fr1A and SPE1-Fr1B, both sub-fractions were dried under vacuum then reconstituted into 100 µL of propan-2-ol. To fraction SPE1-Fr1A, 1 mL of 50 mM phosphate buffer (KH_2_PO_4_ pH 7) containing cholesterol oxidase enzyme (0.264 units) was added and incubated at 37 °C for 1 hr. The reaction was then quenched by the addition of 2 mL of methanol. SPE1-Fr1B was treated in an identical manner but in the absence of cholesterol oxidase. To each fraction 150 µL of glacial acetic acid was then added followed by 190 mg of [^2^H_5_]Girard P hydrazine ([^2^H_5_]GP) to fraction SPE1-Fr1A and 150 mg of [^2^H_0_]GP to SPE1-Fr1B (see Supplementary Figure 1). After a vortex mix, each fraction was left overnight at room temperature.

Each fraction (3.25 mL 69% organic) was loaded onto an Oasis HLB 60 mg (Waters Corp) SPE column (SPE2) and recycled through the column with 50% dilution with water of each eluate until the final eluate was 17.5% methanol. At this point all oxysterols are bound to the column. The column was then washed with 6 mL of 10% methanol and oxysterols eluted in 2 mL of methanol (SPE2-Fr1A and SPE2-Fr1B originating from SPE1-Fr1A and SPE1-Fr1B, respectively). Equal volumes of SPE2-Fr1A and SPE2-Fr1B were combined then diluted to 60% methanol ready for LC-MS(MS^n^) analysis.

SPE1-Fr3 was similarly divided into two equal volumes SPE1-Fr3A and SPE1-Fr3B which were treated in a similar fashion to SPE1-Fr1A and SPE-Fr1B to provide SPE2 samples (SPE2-Fr3A and SPE2-Fr3B) in 3 mL of methanol, ready for LC-MS(MS^n^) analysis after dilution to 75% methanol.

#### CSF

The slow turnover rate (about 3 – 4 times per day)^37^ and the presence of the enzyme lecithin cholesterol acyl transferase (LCAT) upon lipoprotein particles in CSF results in the almost complete esterification of hydroxysterols to their fatty acyl derivatives.^35,36^ As 24S-HC and cholesterol were the primary focus of our study a saponification step was carried out to hydrolyse these and other sterol/oxysterol esters to allow the measurement of total 24S-HC, total cholesterol and of other sterols and oxysterols. CSF (100 µL) was added dropwise to a freshly prepared solution of 1.050 mL 0.35 M ethanolic KOH containing deuterated oxysterol and sterol standards (See Supplementary Table S1). The solution was incubated at room temperature in the dark for 2 hr, after which it was neutralised by addition of 350 µL of water containing 21.1 µL of glacial acetic acid. The mixture was then centrifuged at 14,000 x g at 4°C for 30 min to remove any precipitated matter and applied to SPE1. The eluate was combined with a column wash of 5.5 mL of 70% ethanol to give SPE1-Fr1. The remainder of the protocol was followed as for plasma except SPE1-Fr3B was not dried down, instead it was used directly for LC-MS(MS^n^) analysis in the absence of derivatisation to detect sterols transparent to derivatisation.

### LC-MS(MS^n^)

LC-MS(MS^n^) was performed on an Ultimate 3000 UHPLC system linked to an Orbitrap Elite hybrid mass spectrometer (both Thermo Fisher Scientific) as described in Yutuc *et al*.^36^ For oxysterol analysis, samples were injected in 60% methanol and LC separation was achieved on a Hypersil Gold C_18_ (Thermo Fisher Scientific) reversed-phase column (50 x 2.1 mm, 1.9 µm) utilising a gradient of methanol/acetonitrile/water containing 0.1% formic acid at a flow rate of 200 µL/min. Gradients of 17 min and 37 min were employed (see Supplementary Material 1 Methods). For cholesterol and cholesterol precursor analysis, samples were injected in 75% methanol onto an ACE (Advanced Chromatography Technologies) C_18_ reversed-phase column (10 cm x 2.1 mm, 2 µm) the gradient duration was 24 min. Electrospray ionisation (ESI) in the positive-ion mode was employed and the Orbitrap operated at 120,000 resolution (FWHM at *m/z* 400) for the 17 and 24 min gradients and at 240,000 for the 37 min gradient. Mass accuracy was typically better than 5 ppm. While the Orbitrap was recording a high resolution MS^1^ scan, the linear ion trap (LIT) was simultaneously recording up to five MS^3^ scans. Quantification was performed exploiting high-resolution MS^1^ scans by isotope dilution, or by using isotope-labelled compounds of similar structure to the target analytes. Details of the gradients employed and scan events are provided in Supplementary Material 1 Methods.

The derivatisation method (depicted in Supplementary Figure S1) requires the target analyte to have a 3β-hydroxy or an oxo group. Most oxysterols and the immediate precursors of cholesterol possess such a function. However, the derivatisation fails on substrates with a 4,4-dimethyl function such as in lanosterol. Hence lanosterol was quantified without derivatisation as the [M+H-H_2_O]^+^ ion against [^2^H_7_]lanosterol in a high-resolution (120,000 at *m/z* 400) MS^1^ scan recorded on an Orbitrap IQX instrument. An aliquot of fraction SPE1-Fr3B was diluted to 80% ethanol 0.1% formic acid and injected onto a Hypersil Gold C_18_ column. The LC gradient was 17 min with the mobile phase methanol/acetonitrile/water containing 0.1% formic acid. Molecular identification was assisted by MS^2^ scans in the LIT (see Supplementary Material 1 Methods).

### Sample Batches

All samples were prepared and analysed with the analyst blinded to sample demographics. Samples were prepared in batches of 8, with one water blank (where water substituted for CSF or plasma during the sample preparation protocol) and one laboratory quality control (QC) sample of the appropriate biomaterial.^36^ In some cases, the NIST SRM 1950 (standard reference material) was substituted for one of the plasma samples.^38^ Prior to analysis of each batch of samples mass accuracy and chromatographic performance were check by injection of a derivatised oxysterol standard (19-hydroxycholesterol, 19-HC). Each sample was injected multiple times to allow the acquisition of an MS^3^ spectrum for each analyte to confirm its identity (see Supplementary Material 1 Methods). After analysis of each sample the column was washed with two injections of 50% propan-2-ol in methanol followed by one injection of derivatised 19-HC in 60% methanol which acted as a sensitivity and mass accuracy test and could also monitor any carry over. Throughout the analytical run mass accuracy was also monitored by reference to internal isotope-labelled standards.

### Statistical Analysis

Four hundred plasma and 400 CSF samples were analysed from three groups of donors consisting of control (n = 81), premanifest HD (n = 124) and manifest HD (n = 183, see Table 1). Twelve individuals donated two samples.

Due to skewness of most of the variables, the assay concentration analyses were based on log transformations. These were treated as outcome variables in linear models with HD group; manifest (motor-diagnosed) HD, premanifest HD, and non-CAG-expanded controls. The models controlled for age, sex and their interaction, regardless of significance of these covariates. In the linear models when testing potential confounding by medications, use of statins and the antipsychotics haloperidol and aripiprazole were controlled with separate indicator variables for use of each medication. Because some participants underwent repeated assay collection, the models also included a random intercept term per participant. Significance of overall group differences (group trends) is based on two degree-of-freedom maximum likelihood tests for the overall effect of HD group. Reported *P* values for difference between any pair of groups (typically manifest HD versus controls) used the Tukey adjust for multiple pairwise comparisons.

The classification analyses were carried in binary fashion for the three possible pairwise comparisons amongst control, premanifest and manifest. The purpose was to develop models to identify the most useful metabolites for predicting membership of the three groups, and for future biochemical study. Seven classification techniques were used, forward stepwise logistic regression, backwards stepwise logistic regression, the regression regularisation techniques lasso, ridge and elastic net, and the machine learning techniques random forest and gradient boosting. In regression modelling variables are assessed ‘controlling for the effects of other variables included in the model’. In random forest and gradient boosting, variable importance is evaluated in the context of all other predictors and interactions are automatically captured through the tree structures these techniques use. Model evaluation was made with the receiver operating characteristic (ROC) curve and area under the curve (AUC), histogram of probability of group membership, 2×2 confusion table, and associated statistics. In addition, performance was assessed with five-fold cross-validation supplemented by permutation analysis. Ranking of variables by the different techniques were combined to make a consensus using the Stuart rank aggregation method. R was used for analysis. An extended account of the binary classification techniques is given in Supplementary Material 2.

## Results

### Plasma

Our LC-MS(MS^n^) analysis of plasma gave quantitative data for 25 different oxysterols, cholesterol itself, four precursors and two sterols of plant or microbial origin (see Table 2, Figure 1 and Supplementary Figure S2).

**Figure 1.**
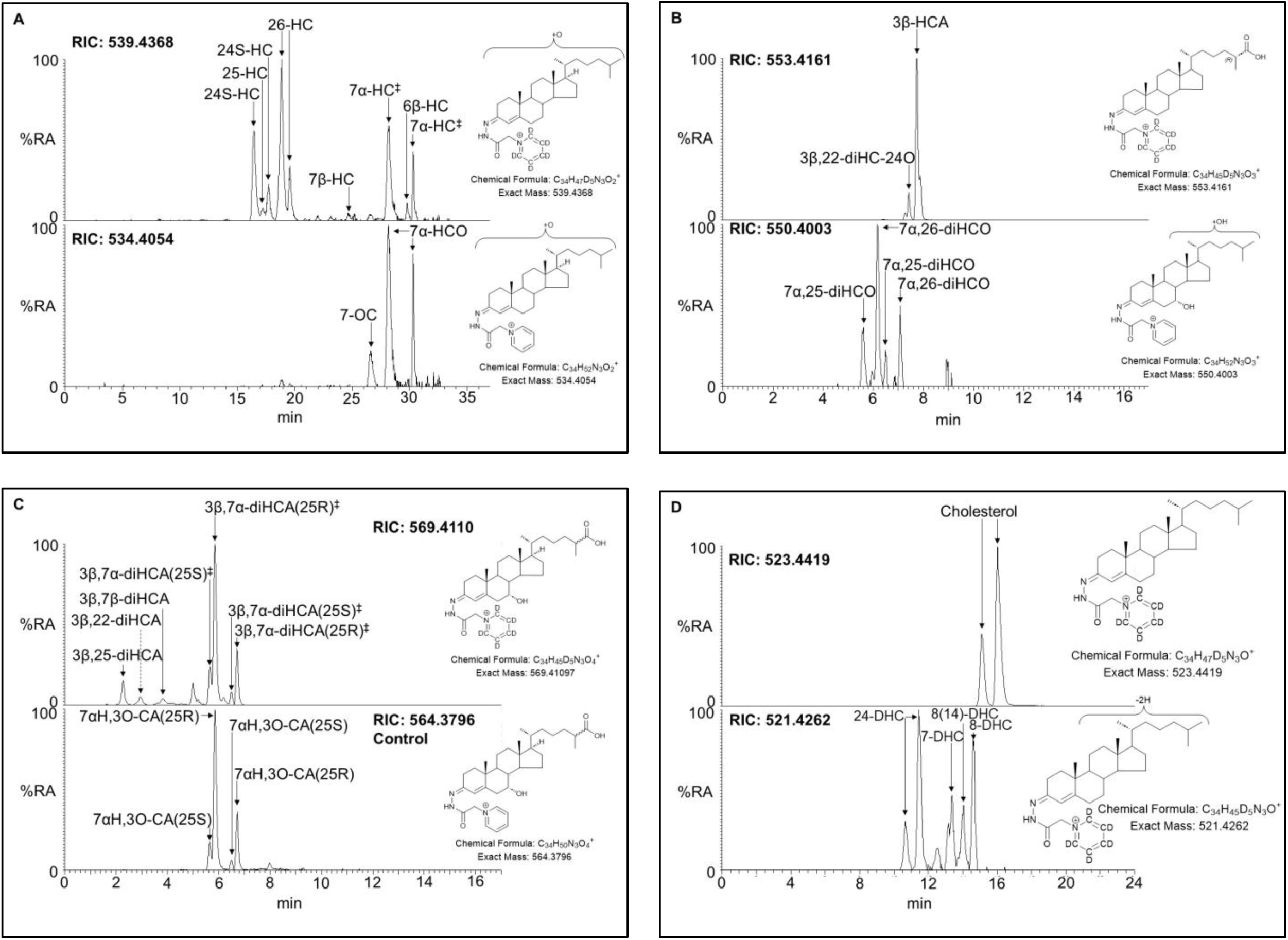
LC-MS analysis of plasma showing reconstructed ion chromatograms (RICs ± 5 ppm) of different oxysterol/sterols. (**A**) Upper panel *m/z* 539.4368 corresponding to hydroxycholesterols (HC), and lower panel *m/z* 534.4054 corresponding to hydroxycholestenones (HCO), the symbol (‡) indicates 7α-HC and 7α-HCO are measured in combination. (**B**) Upper panel *m/z* 553.4161 corresponding to hydroxycholestenoic acids (HCA) and lower panel *m/z* 550.4003 corresponding to dihydroxycholestenones (diHCO). (**C**) Upper panel *m/z* 569.4110 corresponding to dihydroxycholestenoic (diHCA) and hydroxyoxocholestenoic (HOCA) acids and lower panel *m/z* 564.3796 corresponding HOCA alone, the symbol (‡) indicates 3β,7α-diHCA and 7αH,3O-CA measured in combination. (**D**) Upper panel *m/z* 523.4419 corresponding to cholesterol, and lower panel *m/z* 521.4262 corresponding to dehydrocholesterols (DHC). Odd *m/z* values correspond to sterols/oxysterols derivatised with [^2^H_5_]GP, even *m/z* correspond to molecules derivatised with [^2^H_0_]GP. As a consequence of the derivatisation giving cis and trans configurations about the C=N double bond, some sterols/oxysterols give two chromatographic peaks which may or not be resolved. See Supplementary Table S2 for compound abbreviations.

**Table 2.**
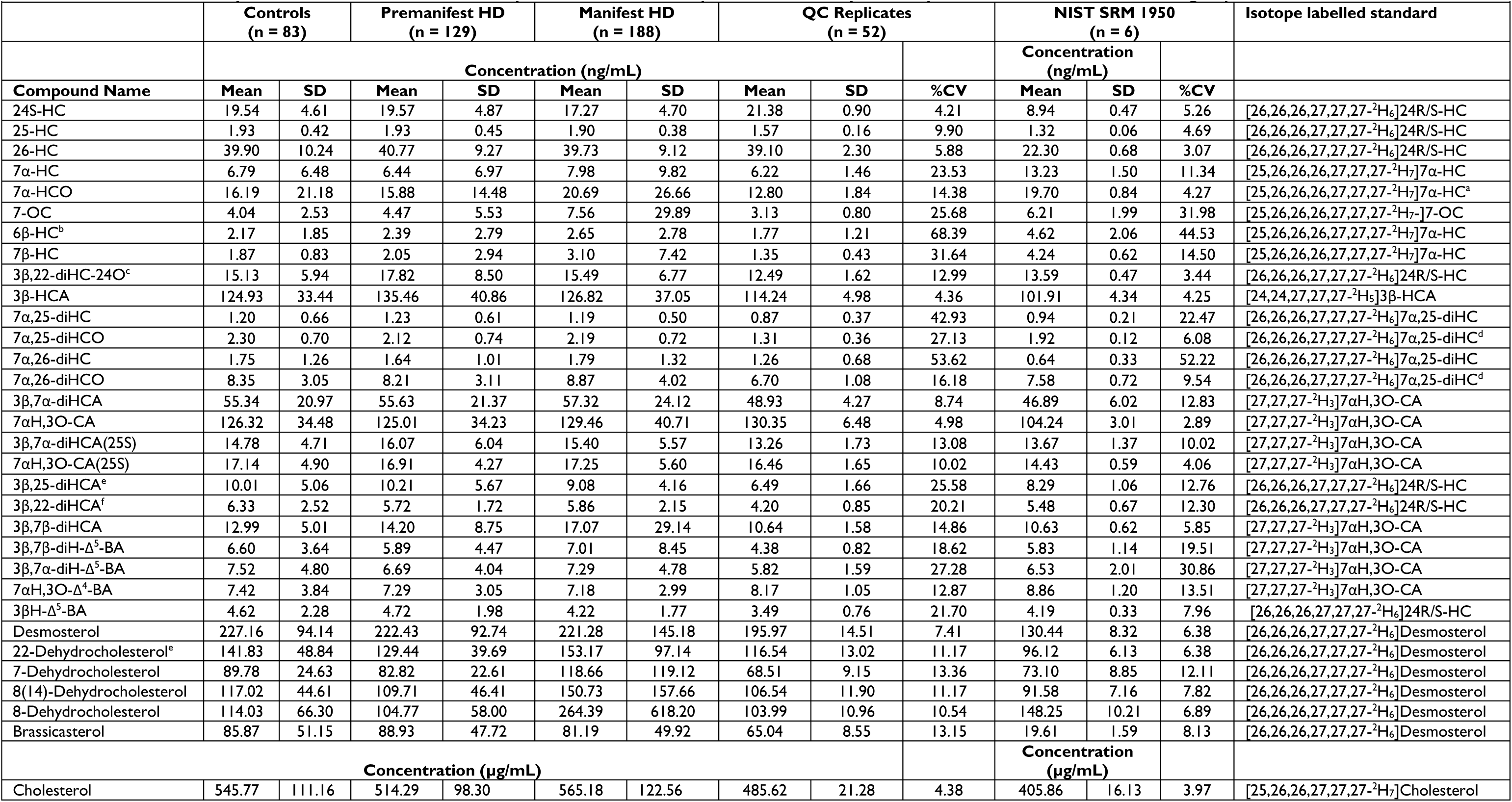
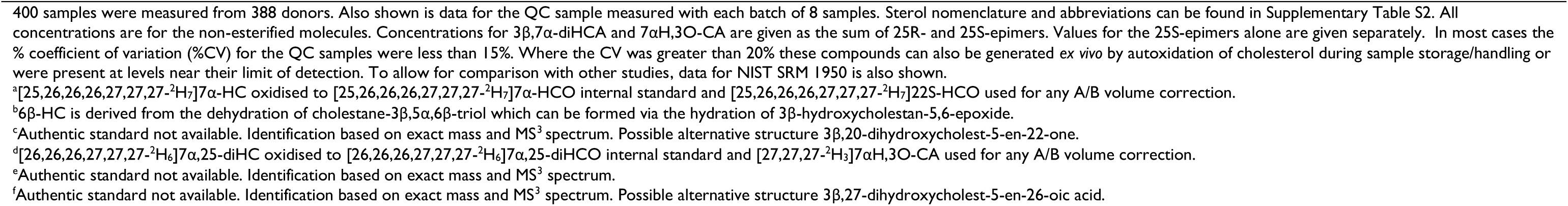
Concentrations of oxysterols, cholesterol and cholesterol precursors measured in plasma from healthy controls, people with premanifest HD and manifest HD.

Initially we investigated how 24S-HC and cholesterol varied between healthy control, premanifest HD, and manifest HD groups measured in both plasma and CSF. Shown in Figure 2 are the plasma data for 24S-HC and cholesterol. Substantial age differences between premanifest HD and the other two groups (see Table 1) dictate consideration of the confounding effects of age. For plasma, treating premanifest HD and manifest HD as categorical variables, considering age as a potential confounder, controlling for sex and performing linear regression analyses after natural log transformation to decrease skewness of data, the concentration 24S-HC was found to be lower in the manifest HD group than in the control group (*P* = 0.0072) and after controlling for statin use, 24S-HC was lower in the manifest HD group than the control (*P* = 0.0058) and premanifest groups (*P* = 0.0418, *P* values with Tukey multiple group comparisons correction). Statistical analysis for cholesterol found no overall group differences, however, an exploratory model revealed higher concentrations in the manifest HD group than controls in younger doners but the opposite for older donors (Figure 2). Unsurprisingly, statins have a strong effect on cholesterol levels (see Supplementary Figure S3), but no groupwise differences were observed after controlling for statins.

**Figure 2.**
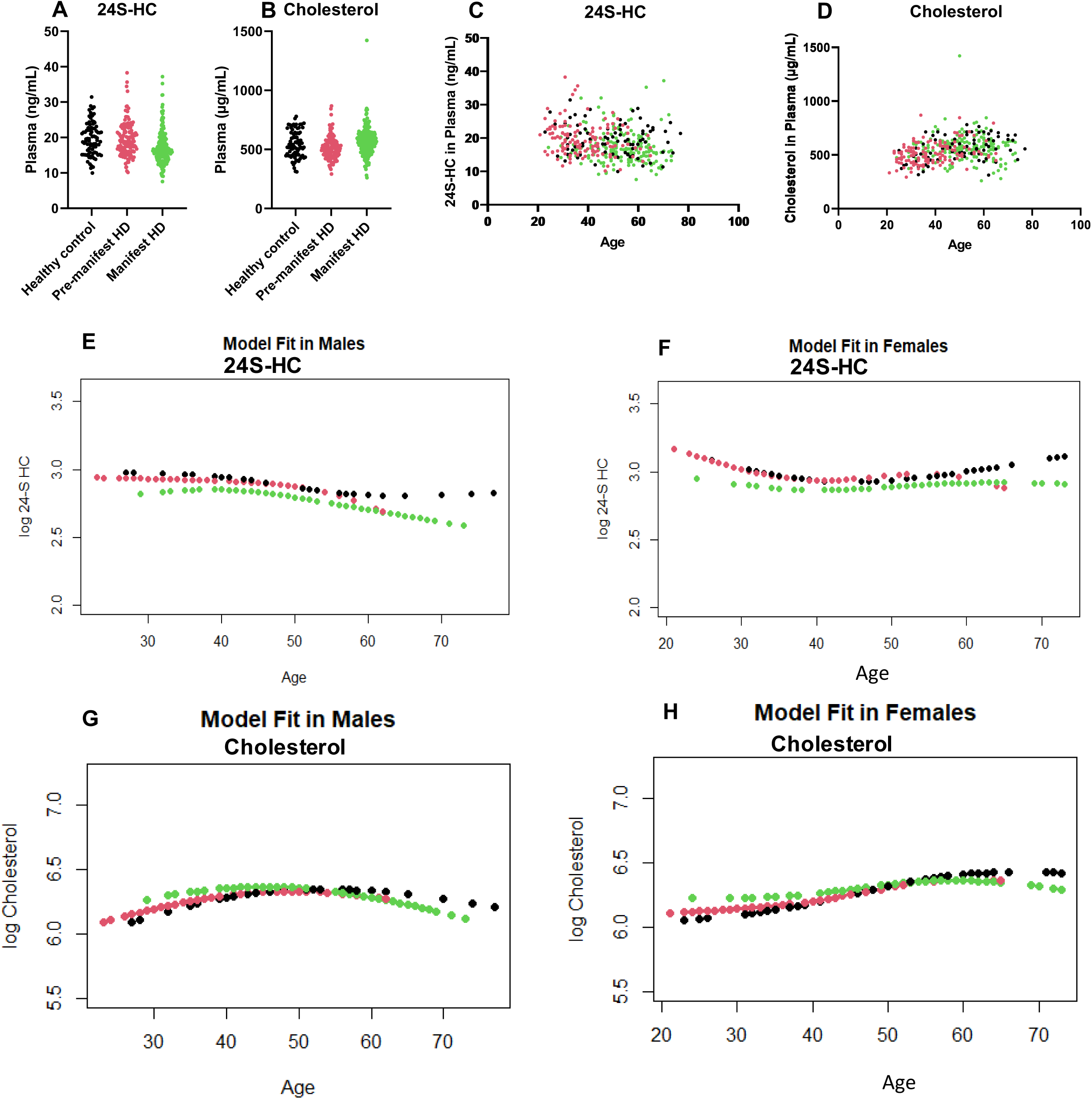
Plasma concentration of 24S-HC but not cholesterol shows group differences between manifest HD, premanifest HD and controls after considering age as a potential confounder. Concentration of (**A**) 24S-HC and (**B**) cholesterol, in each sample analysed. Plot of concentration of (**C**) 24S-HC, and (**D**) cholesterol against donor age. The dots are colour coded according to donor group. Considering age as a potential confounder and modelled by a significant 3-df natural spline interaction with sex, mean levels of 24S-HC are lower in manifest HD than in the other two groups in both (**E**) males and (**F**) females (sex interactions with the HD groups were nonsignificant. Statin medication effect, which does not significantly interact with other model predictors, is not shown). Similar model for cholesterol are shown in (**G**) males and (**H**) females. There is a significant HD group interaction with age. The concentrations are for the non-esterified molecules.

We performed a secondary analysis on the remaining 24 oxysterols, using the same linear regression analysis as for 24S-HC and cholesterol and controlling for age and sex and their interactions. No significant differences were found between the oxysterol levels in the different groups.

We conducted further statistical analysis on the cholesterol precursors 7-DHC, 8-DHC, their isomer 8(14)-dehydrocholesterol (8(14)-DHC), and desmosterol (24-DHC), besides the plant sterol brassicasterol and 22-dehydrocholesterol (22-DHC) of possible plant or fungal origin (Supplementary Figure S2).^39,40^ Controlling for age, sex and their interactions and considering natural log of concentrations, comparison of control, premanifest HD and manifest HD groups revealed substantial trends in order of severity for 7-DHC (*P* = 0.0001), 8-DHC (*P* = 0.0003), 8(14)-DHC (*P* = 0.0027) and desmosterol (*P* = 0.0198), where the sterols 7-DHC, 8-DHC and 8(14)-DHC were increased in the manifest HD group compared to premanifest and control groups but desmosterol decreased. The group differences remained significant after controlling for cholesterol lowering medications or for the antipsychotics medications haloperidol and aripiprazole, known to effect concentrations of dehydrocholesterols in blood.^31^ After adjusting for statins, haloperidol and aripiprazole simultaneously, 7-DHC (*P* = 0.008) and 8-DHC (*P* = 0.009) were still increased in manifest HD compared to controls and 7-DHC (*P* = 0.002) and 8-DHC (*P* = 0.004) were still elevated in manifest HD compared to premanifest HD. Notably, neither haloperidol nor aripiprazole had a detectable effect on plasma concentrations of 24S-HC.

### CSF

In contrast to the analysis of plasma samples, we analysed CSF after a saponification step to hydrolyse sterol esters; this was necessary due to the very low concentration of non-esterified hydroxycholesterols compared to their esters (i.e. about 2%) in CSF. The data is summarised in Table 3 and the entire data set in dot plot form shown in Supplementary Figure S4.

**Table 3.**
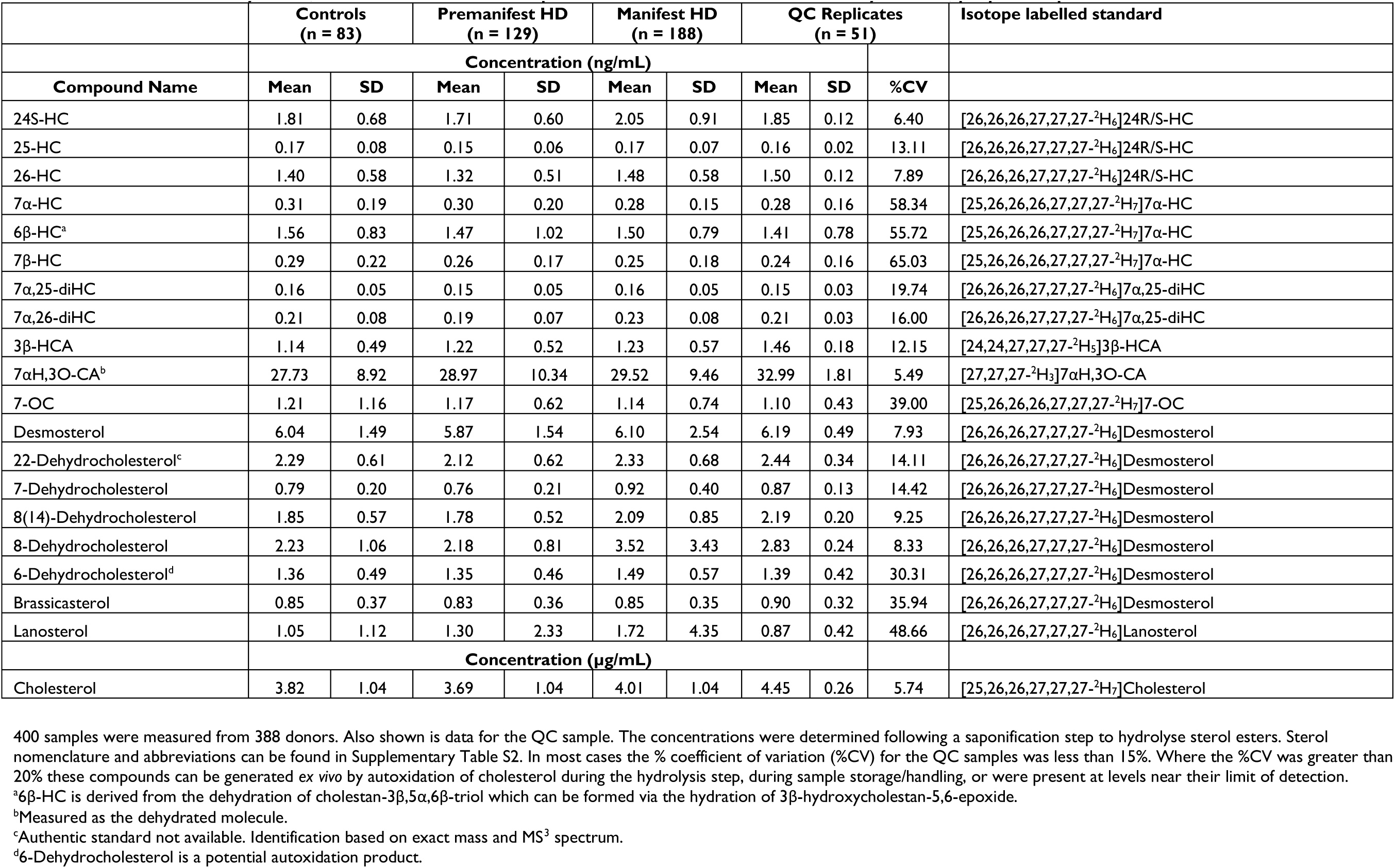
Concentrations of oxysterols, cholesterol and cholesterol precursors measured in CSF from healthy controls, people with premanifest HD and manifest HD.

Eleven oxysterols were quantified in CSF including 24S-HC (Figure 3). Following a statistical procedure similar to that outlined for plasma, treating premanifest HD and manifest HD as categorical variables and controlling for age and sex, no significant group differences were observed for 24S-HC and there was no significant statin effect on 24S-HC in CSF. Similarly, after age and sex corrections, there were no group differences in cholesterol CSF levels and no statin effect on CSF cholesterol. Interestingly, haloperidol significantly lowers 24S-HC concentrations in CSF, but 24S-HC levels in CSF are not associated with HD status, regardless of haloperidol.

**Figure 3.**
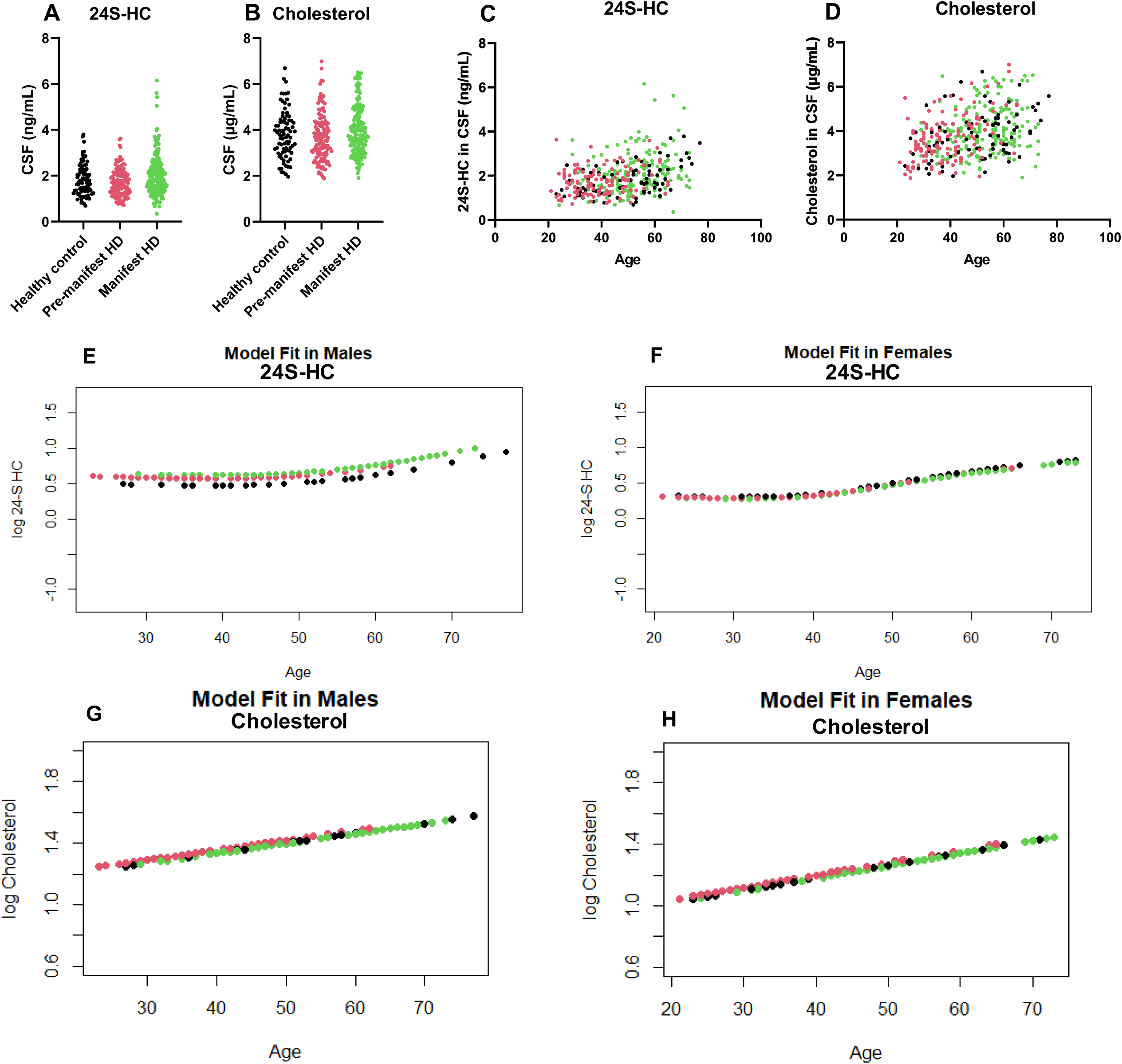
Neither CSF concentrations of 24S-HC nor of cholesterol show group differences between manifest HD, premanifest HD and controls after controlling for age. Concentration of (**A**) 24S-HC and (**B**) cholesterol in each sample. Plot of concentration of (**C**) 24S-HC, and (**D**) cholesterol against donor age. The dots are colour coded according to donor group. Model controlling for sex and nonlinear (significant 3 df natural spline) age relationships to (**E & F**) 24S-HC in CSF but finding no significant association with HD status. Similar model for (**G & H**) cholesterol showing age and sex effects but no HD group differences. The concentrations were determined after ester hydrolysis.

A secondary analysis was performed on 10 further oxysterols (Table 3), performing the same HD severity regression analysis as for the primary hypothesis controlling for age and sex and their interactions. No severity trends were observed.

Additional statistical analysis was performed on the cholesterol precursors 7-DHC, 8-DHC, 8(14)-DHC, desmosterol, lanosterol, besides the plant sterol brassicasterol, 22-DHC and 6-dehydrocholesterol (6-DHC), a probable autoxidation product of cholesterol (see Supplementary Figure S4 for dot plots). Severity trends were observed for 7-DHC (*P* = 0.0155), and 8-DHC (*P* = 0.0002) with these metabolites increased in the manifest HD group compared to the other two groups. After adjustment for statin and antipsychotic medications *P* values remined similar for 7-DHC (*P =* 0.0209 for manifest HD v control and *P* = 0.0285 for manifest HD v premanifest) and 8-DHC (*P* = 0.0013 for manifest HD v control and *P* = 0.0183 for manifest HD v premanifest).

### Comparison of Binary Classification Techniques

An extended description of the binary classification results is given in Supplementary Workbook 1 (available to view on figshare https://doi.org/10.6084/m9.figshare.30510515) with a summary in Supplementary Workbook 2 (available to view on figshare https://doi.org/10.6084/m9.figshare.30510515) and narrative in Supplementary Material 2. In Supplementary Figure S5 ROC curves for the premanifest HD and manifest HD comparison are shown for the seven classification methods. ROC curves show clinical sensitivity and specificity where the area under a ROC curve (AUC) is a measure of the discriminative ability of a test.^41^ Tests with good diagnostic performance have a ROC curve that is close to the top-left corner, with an area under the curve close to 1. The AUC values for the premanifest HD and manifest HD comparison for the full dataset are consistently high (0.802 – 0.933). Similar AUC values were obtained for the means of the five test samples from cross-validation analysis (0.802 – 0.848). For the two binary comparisons involving control, the full dataset also gives high values of AUC and significant values for the confusion table. However, for the cross-validation test samples the AUC values are lower and many of the *P*-values for the confusion tables are not significant (see Supplementary Workbooks on figshare https://doi.org/10.6084/m9.figshare.30510515)

Ranked lists of the metabolites most useful in distinguishing between the three groups are detailed in the Supplementary Workbooks. The consensus ranking for the premanifest HD v manifest HD comparison is shown in Table 4. The highest consensus ranked metabolites are plasma 24S-HC followed by plasma 7-DHC, plasma cholesterol, CSF 24S-HC, CSF 3β-hydroxycholest-5-en-(25R)26-oic acid (3β-HCA), CSF 8-DHC and plasma 3β-HCA.

**Table 4.**
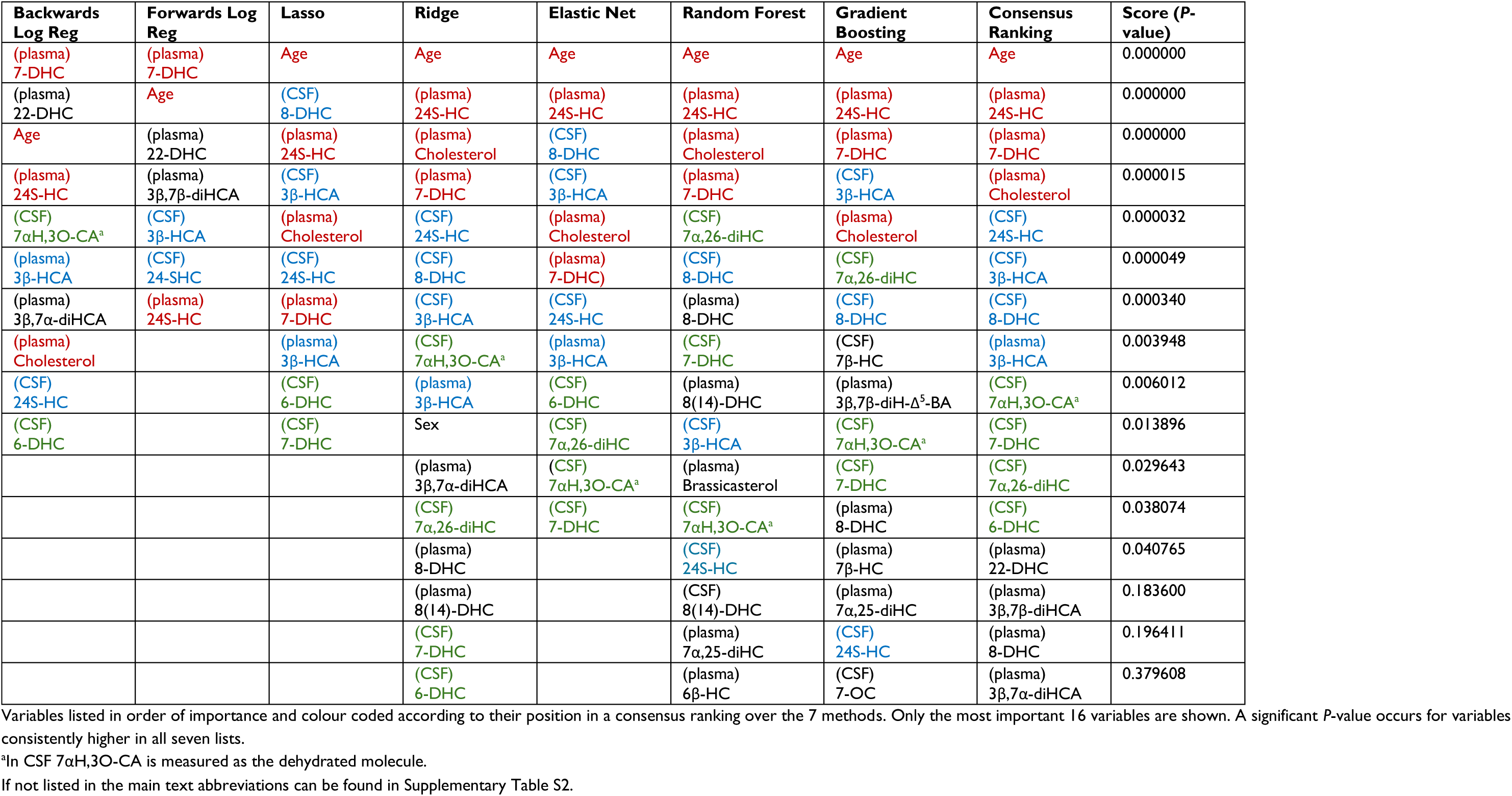
Independent variables retained as being important in seven classification models for comparison of premanifest and manifest HD.

## Discussion

Disordered cholesterol homeostasis in brain is implicated in a number of neurodegenerative diseases, including HD.^6,20,42,43^ This is not surprising considering the high cholesterol levels in the CNS.^15^ Cholesterol in brain is isolated from the circulation by the BBB, which is impermeable to cholesterol itself but not to oxysterols, its oxidised forms.^18^ Essentially all cholesterol in brain is made *in situ*, but oxysterols can be synthesised in brain (i.e., 24S-HC) and also the periphery.^10,22–24^ While 24S-HC is exported down a concentration gradient from brain to the circulation,^16^ other oxysterols e.g. (25R)26-hydroxycholesterol (26-HC; more commonly known by the non-systematic name 27-hydroxycholesterol)^44^ is imported via passive diffusion to brain and metabolised further to the cholestenoic acid 7α-hydroxy-3-oxocholest-4-en-(25R)26-oic acid (7αH,3O-CA(25R)).^22–24^ Cholesterol deficiency in brain has been implicated in HD pathology,^6^ and its supplementation proposed as a treatment.^9,11^ Conversely, increasing cholesterol metabolism via the neuronal enzyme CYP46A1 has also been suggested as a treatment.^13,14,42^ These two potential treatment regimens appear contradictory.

24S-HC is formed from cholesterol in neurons in a CYP46A1 catalysed reaction (Figure 4),^17^ and 24S-HC in the circulation is essentially all derived from brain^16^; its circulatory concentrations can be considered a measure of the number of metabolically-active neurons in brain.^10,19,20^ 24S-HC exists along with its epimer 24R-HC, and invariably 24S-HC is the dominant isomer,^18^ however, both isomers are present in human plasma and should ideally be resolved. This is achieved in the present study,^36^ but seldom achieved by other methods where the combined measurement of 24S-HC and 24R-HC (indicated here by the abbreviation 24-HC) can compromise accurate quantification of the brain-derived S-epimer. 24-HC has previously been measured in the circulation of PwHD and found to be at lower concentrations in a manifest PwHD group than premanifest PwHD or control groups.^28–30^ However, premanifest PwHD are usually younger than those with manifest HD, and age was not considered as a confounder in these studies,^28–30^ although one study did suggest that 24-HC concentrations did not vary with age for the populations investigated.^28^ Despite being meticulously performed using gas chromatography – mass spectrometry (GC-MS), a disadvantage of these studies was that 24-HC was measured following a saponification step where fatty acyl esters of 24-HC are hydrolysed and the total 24-HC then measured, rather than just the non-esterified molecule which is the species that actually crosses the BBB into the circulation.^28–30^ Additionally, 24S-HC was not specifically measured, but rather the R- and S-epimers in combination.

**Figure 4.**
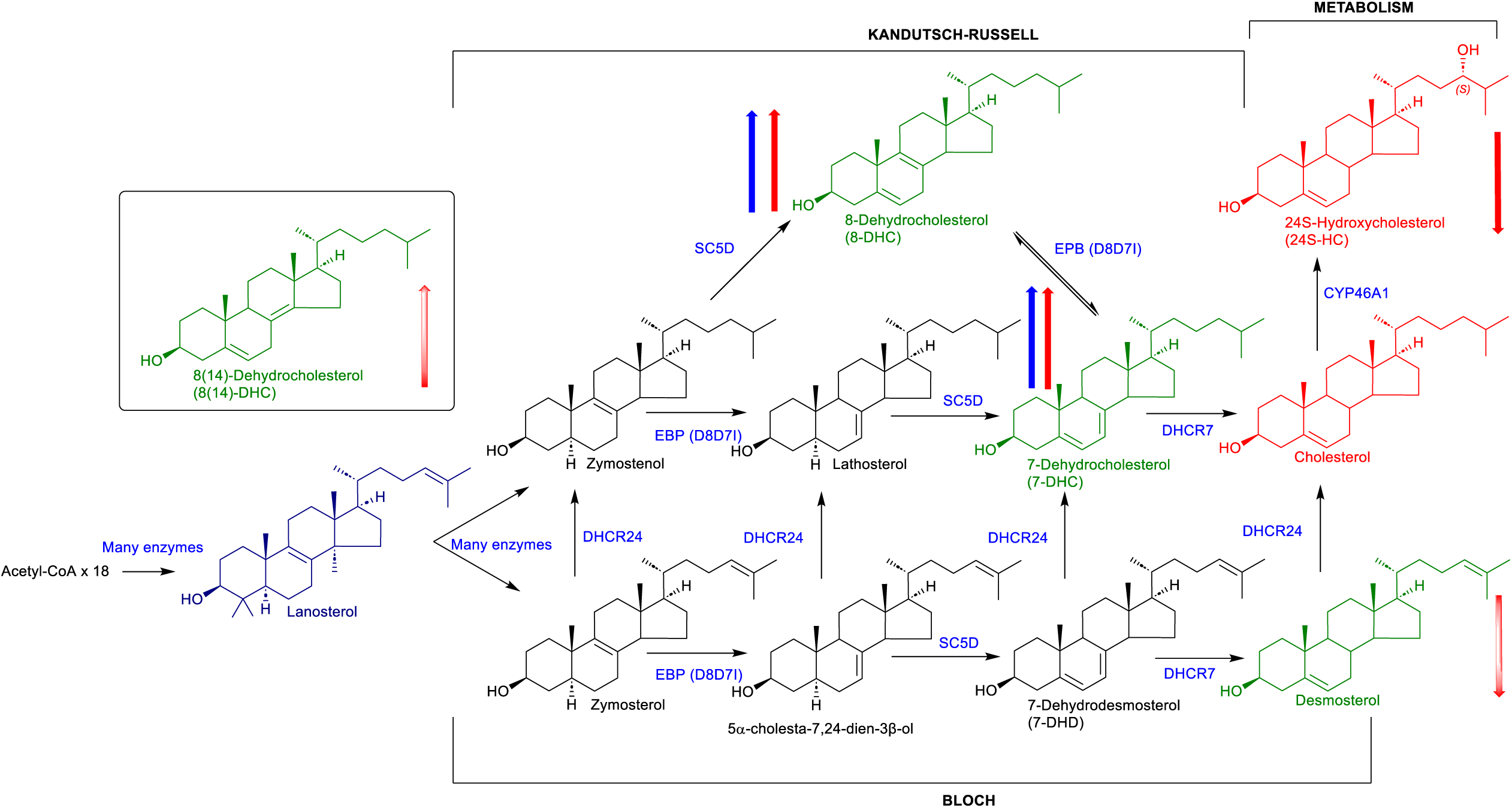
Simplified scheme of cholesterol biosynthesis and metabolism to 24S-HC. Quantified sterols are colour coded: Cholesterol and 24S-HC are shown in red, cholesterol precursors demosterol, 7-DHC, 8-DHC and their isomer 8(14)-DHC are shown in green, lanosterol measured without derivatisation is shown in dark blue. Thick red or blue arrows indicate the direction of concentration change according to HD severity in plasma and CSF, respectively. Statin or antipsychotic medications did not alter these trends except in the case of desmosterol and 8(14)-DHC in plasma where significance was lost after adjusting for medications (indicated by a shaded red arrow).

The previous studies highlighted above suggest that 24S-HC in the circulation might be a biomarker for the progression of HD towards the fully manifest disorder. In the current study we attempted to confirm this hypothesis by exclusively measuring 24S-HC, rather than the unresolved mixture of 24S- and 24R-HC, and measuring the non-esterified free molecule that actually crosses the BBB into the circulation. We also considered age as a potential confounder.

As can be seen in the model developed in Figure 2E, concentrations of 24S-HC in plasma fall with age in males with manifest HD. When age was treated as a potential confounder, controlling for age by sex interactions and treating premanifest HD and manifest HD as categorical variables, levels of 24S-HC in the manifest PwHD group were lower than in controls (*P* = 0.0072) and, after correcting for the use of statins, levels of 24S-HC in the manifest PwHD group were lower than in both control (*P* = 0.0058) and premanifest PwHD (*P* = 0.0418) groups. Unsurprisingly, statins were associated with substantially lower levels of 24S-HC (see Supplementary Table S3A & S3B). As the statin medications were a mixture of uncertain-, low- and brain-penetrant, the low level of 24S-HC was probably due to statin-induced upregulation of the LDL receptor in liver.^45^ The antipsychotic medications aripiprazole and haloperidol did not have a detectable effect on 24S-HC plasma levels. Although our data does not suggest that plasma 24S-HC is a prognostic biomarker at the individual level, it could be used to monitor a pharmacodynamic response for groups of patients or perhaps individuals (See Figure 2E & F).^46^

Recently, in agreement with GC-MS results described above,^28–30^ Gray et al^47^ using LC-MS/MS also found that the level of total 24S-HC in a cohort of manifest PwHD was lower than in a premanifest PwHD group, however, age was not considered as a confounder and the premanifest PwHD group was on average about 10 years younger than the manifest PwHD group.

24S-HC can also cross the blood CSF barrier, but the export rate via this route is estimated to be about 1 µg/day compared to 6 mg/day over the BBB.^25^ CSF has a slow turnover rate (about 3 – 4 times per day)^37^ and the presence of the LCAT enzyme in lipoprotein particles results in the majority of 24S-HC in CSF existing as fatty acyl esters (98%) with only pg/mL concentrations of the non-esterified molecule present.^35^ Thus, in our study of CSF we measured the total 24S-HC following a hydrolysis step. Age and sex effects on the concentrations of 24S-HC in CSF were found to be highly significant, but after age adjustment there were no significant associations of 24S-HC with HD status (Figure 3E & F). Similarly, after correcting for statin use there were no significant group differences and CSF 24S-HC levels were not associated with HD status regardless of haloperidol or aripiprazole use.

Cholesterol is the substrate for CYP46A1 catalysed formation of 24S-HC (Figure 4), however, unlike 24S-HC, cholesterol found in the circulation is of extracerebral origin. Despite this, its measurement in plasma may be of value in the study of HD as mutant HTT is widely expressed and any effect it may have on cholesterol synthesis may also be evident in the periphery. Cholesterol levels in plasma, measured as the non-esterified molecule, were found to show sex and non-linear age relationships but no overall HD group difference. However, as shown in Figure 2G and 2H, cholesterol levels in manifest PwHD were higher than in controls in younger donors (see ages 30 – 40 years in Figures 3G & H) but lower than controls when older (60 – 70 years) and HD severity is associated with a significantly greater cholesterol decrease with age. Unsurprisingly, statins have a strong effect on cholesterol levels (see Supplementary Table S3), but no groupwise differences were observed after controlling for statins. In earlier studies of total cholesterol concentration in plasma, no differences between control, premanifest HD and manifest HD groups were found.^28^ However, levels of cholesterol were found to fall in the latter stages of manifest HD when compared to the earlier stages.^29^

In the current study we also measured (as the non-esterified molecules) in plasma the cholesterol precursors 7-DHC, 8-DHC, their isomer 8(14)-DHC and desmosterol (Supplementary Figure S2). After controlling for age, sex and their interactions, plasma concentrations of 7-DHC, 8-DHC and 8(14)-DHC were found to be elevated in the manifest HD group compared to premanifest HD and control groups, while desmosterol was diminished in the manifest HD group compared to the other two groups. The group differences for 7-DHC and 8-DHC remain significant after controlling for statins and simultaneously for statins and the two antipsychotic medications. 7-DHC and 8-DHC are members of the Kandutsch-Russell pathway of cholesterol biosynthesis where the sterol side-chain double bond becomes saturated in an early step (Figure 4). Alternatively, desmosterol is the immediate precursor of cholesterol in the Bloch pathway where the sterol side-chain remains unsaturated up to the final step. The abundance of the pathway intermediates 7-DHC, 8-DHC and desmosterol can be regarded as a measure of the relative importance of the Kandutsch-Russell and Bloch pathways in cholesterol synthesis. Thus, our data suggests that the Kandutsch-Russell pathway becomes more important for the non-cerebral synthesis of cholesterol in people with manifest HD than for people with premanifest HD or controls. This does not agree, however, with previous reports using GC-MS and measuring total sterols after a saponification step where lathosterol, another marker of the Kandutsch-Russell pathway, was reduced in premanifest and some groups of manifest HD people compared to controls.^29^ However, lathosterol is not a requisite intermediate of the Kandutsch-Russel pathway as 7-DHC can be formed via 8-DHC rather than lathosterol (see Figure 4).^48^ It should be noted that in both our LC-MS(MS^n^) study and in the earlier GC-MS study,^29^ the concentrations of cholesterol precursors varied greatly in the individual groups.

While analysis of plasma will give information on the extracerebral effects of mutant HTT on sterol biochemistry, measurements in CSF can give a better picture of cholesterol synthesis in brain. Studies using deuterium-labelled cholesterol in a human volunteer indicate that very little of the cholesterol content of CSF is derived from plasma, and this is also presumably true for cholesterol precursors.^49^ Our results for total cholesterol in CSF revealed no group differences between control, premanifest HD and manifest HD groups although age and sex effects were present (Figure 3G & H). Interestingly, no statin effects were found on CSF cholesterol. Considering the cholesterol precursors in CSF, and controlling for age, sex, and their interactions, both 7-DHC and 8-DHC were elevated in manifest HD compared to premanifest HD and control groups, and again statins did not affect the significance of group differences, nor did controlling for statins and antipsychotic medications. This data reinforces the suggestion that the Kandutsch-Russell pathway is increased in people with manifest HD. An alternative explanation for these results could, however, be a decreased activity of the enzyme DHCR7 causing a build-up of its substrate 7-DHC and its isomer 8-DHC.

Besides analysing 24S-HC, cholesterol and its precursors in plasma and CSF, we also quantified multiple other metabolites that gave us information on 55 independent variables, of which 51 were the measured levels of metabolites in either plasma or CSF besides, age, BMI, sex (male or female) or taking statins (yes or no). These variables were used in seven binary classification techniques to provide a means of both effectively classifying HD individuals and identifying the most important independent variables to be used for classification. The classification of premanifest PwHD and manifest PwHD were the most successful of the three comparisons. This is the most important classification in the drug discovery process where the goal is to prevent progression into the manifest HD state. The AUC for the ROC curves for the means of the five-fold cross validation test samples, which provide a robust estimate of out-of-sample classification performance, gave values varying from 0.802 to 0.848 (Supplementary Workbook 2, sheet AUC 2×2, available to view on figshare https://doi.org/10.6084/m9.figshare.30510515) corresponding to a good to very good diagnostic for the premanifest HD against manifest HD comparison.^41^ The independent variables most important in the seven classification models for comparison of premanifest HD and manifest HD were: age, plasma 24S-HC, plasma 7-DHC and plasma cholesterol (Table 4). This is compatible with the trend analysis described above where plasma concentrations of 24S-HC and 7-DHC were found to be different between these donor groups and where there is evidence of an age by HD severity interaction.

The promising results for the binary classification techniques for differentiation of premanifest HD from manifest HD status is encouraging, and in future studies we plan to use models generated from the current data to predict group classification in a new set of samples.

Recently, the Genetic Modifiers of Huntington’s Disease (GeM-HD) Consortium performed a GWAS study with a sample size of more than 16,000 HD individuals and identified several new significant signals at two new loci, adding to the number of known HD modifier genes.^50^ The new loci included a regulator of gene expression, *MED15* found on chromosome 22, which codes a subunit of the Mediator complex involved in the regulated transcription of RNA polymerase II-dependent genes and is required for SREBP (sterol regulatory element-binding protein) control of cholesterol and lipid homeostasis.^51^

There are two major SREBP proteins, SREBP1c and SREBP-2 which are master regulators of fatty acid and cholesterol synthesis, respectively. They are coded by the genes *SREBF-1* and *SREBF-2*.^52^ Interestingly, a new landmark-delaying effect (22BM1) was also found on chromosome 22 mapping to a gene dense region including *SREBF-2*.^50^ In combination these results further implicate cholesterol in HD pathogenesis.

In the context of cholesterol metabolism in HD, Chiang et al have determined bile acid levels in plasma of healthy controls, premanifest and manifest PwHD.^53^ They found that the combined HD group had higher levels of glycochenodeoxycholic (GCDCA) and glycoursodeoxycholic (GUDCA) acids than the healthy control group. An elevation in GUDCA in HD is interesting as it has been suggested to be formed from 7-DHC in human,^54^ and in the current study we found 7-DHC to be elevated in the manifest HD group. The results of Chiang should, however, be treated with some caution as confounding effects of age and sex were not considered.^53^

Finally, it is worth considering if the data from the present study favours either of the two proposed sterol-related treatments for HD. One treatment favoured by Cattaneo, Valenza and colleagues is to increase the cholesterol content of striatum, the area of brain most affected by HD,^9,11,12^ the other is to increase cholesterol metabolism and is favoured by Cartier, Betuing and colleagues.^13,14^ Our data could be explained by the hypothesis of Cattaneo and Valenza in that there is a deficiency of cholesterol in striatum and this deficiency would lead to a decreased production of 24S-HC in striatum resulting in decreased flux of 24S-HC across the BBB into the circulation. Our data would also be compatible with the suggestion of Cartier and Betuing that there is a striatal-deficiency of CYP46A1, the enzyme responsible for 24S-HC formation in brain, again leading to a decreased flux of 24S-HC across the BBB. Whether it is a deficiency in cholesterol or deficiency in CYP46A1 which is responsible for the fall of 24S-HC in the circulation is not obvious from our study, however, the availability of oxysterol - mass spectrometry imaging should go some way to answering this question in the near future.^55,56^

In summary, in the current study we have shown that non-esterified 24S-HC, the stereo-specific brain-derived cholesterol metabolite that can cross the BBB, is decreased in plasma of a manifest PwHD group. Although our data does not indicate that 24S-HC is a prognostic biomarker at the individual level, it is an open question whether it could be used to monitor a pharmacodynamic response for groups of PwHD, or perhaps of disease progression in individuals. Studies to answer these questions are being performed with HTT-lowering mouse models. Further, by exploiting binary classification techniques we show that a diagnostic model can be developed giving a good to very good performance according to the AUC in ROC curves to distinguish premanifest PwHD from manifest PwHD. This will be tested in future studies and could be of value in monitoring the performance of response to drug treatment. In addition, the most important metabolites in the binary classification models were plasma levels of 24S-HC and 7-DHC, both of which showed statistical changes in the manifest PwHD group, reinforcing the involvement of cholesterol in HD.

## Data Availability

All data produced in the present study are available upon reasonable request to the authors.

https://doi.org/10.6084/m9.figshare.30510515

## Data availability

Datasets are available as Supplementary material files.

## Acknowledgements

Samples and data used in this work would not be possible without the vital contribution of the research participants and their families in the HD-Clarity and HD-CSF studies. HD-Clarity and HD-CSF are cerebrospinal fluid collection initiatives designed to facilitate therapeutic development for Huntington’s disease. HD-Clarity and HD-CSF are led by Dr Edward Wild and sponsored by University College London. HD-Clarity is funded by CHDI Foundation, Inc. The Medical Research Council UK (MR/M008592/1) funded HD-CSF.

This project was initiated following a “Cholesterol and Huntington’s Disease Advisory Meeting” supported by CHDI Foundation and convened Drs Tom Vogt and Cristina Sampaio in Cambridge, MA, November 15 -16, 2018. The authors thank Dr Simon Noble of the CHDI Foundation for critically reading the manuscript.

For the purpose of open access, the authors have applied a Creative Commons Attribution (CC BY) licence to any Author Accepted Manuscript version arising from this submission.

## Supplementary Material

Supplementary material is available online.

## Supplementary Information Provided

**Supplementary Figure S1 Enzyme-Assisted derivatisation for sterol analysis (EADSA).**

(**A**) The 3β-hydroxy group of oxysterols/sterols is converted by cholesterol oxidase enzyme to a 3-oxo group. This is then reacted with the [^2^H_5_]GP to give a GP-hydrazone. Native oxo groups similarly react with [^2^H_5_]GP. This is illustrated for 24S-HC, 3β,7α-diHCA and 7αH,3O-CA.

(**B**) Oxysterols/sterols are reacted with [^2^H_0_]GP in the absence of cholesterol oxidase. Only molecules with a native oxo group will become derivatised.

**Supplementary Figure S2 Concentrations of oxysterols, cholesterol and cholesterol precursors measured in plasma from healthy controls, people with premanifest HD and manifest HD with values for median, 25^th^ and 75^th^ percentiles.** 400 samples were measured from 388 donors. Each sample is shown as a dot. 6β-Hydroxycholesterol (6β-HC) is derived from the dehydration of cholestan-3β,5α,6β-triol which can be formed via the hydration of 3β-hydroxycholestan-5,6-epoxide. Concentrations for 3β,7α-diHCA and 7αH,3O-CA are given as the sum of 25R- and 25S-epimers. Values for the 25S-epimers alone are given separately. All concentrations are for the non-esterified molecules.

**Supplementary Figure S3 Model considering age as a potential confounder showing on a natural logarithm scale that mean levels of cholesterol in plasma do not differ between manifest HD and the other two groups.** However, there is a strong statin effect on cholesterol levels and that at a young age cholesterol levels for manifest HD are higher than for controls but at older ages the reverse is true. The concentrations are for the non-esterified molecules.

**Supplementary Figure S4 Concentration of oxysterols, cholesterol and cholesterol precursors measured in CSF from healthy controls, people with premanifest HD and manifest HD with values for median 25^th^ and 75^th^ percentiles.** 400 samples were measured from 388 donors. The concentrations were determined following a saponification step to hydrolyse sterol esters. Each sample is shown as a dot. 6β-HC is derived from the dehydration of cholestane-3β,5α,6β-triol which can be formed via the hydration of 3β-hydroxycholestan-5,6-epoxide. 6-Dehydrocholesterol is a potential autoxidation product. 7αH,3O-CA was measured as the dehydrated molecule.

**Supplementary Figure S5 ROC curves for the premanifest HD v manifest HD comparison determined by the different classification methods using the full data set.** (**A**) Logistic Regression Forward. (**B**) Logistic Regression Backward. (**C**) Lasso. (**D**) Ridge. (**E**) Elastic Net.

(**F**) Random Forest. (**G**) Gradient Boosting.

**Supplementary Table S1 Isotope-labelled standards used for sterol and oxysterol quantification.**

**Supplementary Table S2 List of oxysterol/sterol abbreviations.**

**Supplementary Table S3 Concentrations measured in plasma samples from people (A) taking statins and (B) not takin statins and measured in CSF samples from people (C) taking statins and (D) not taking statins.**

**Table S4. Consensus ranking lists of seven binary classification techniques for each of three pairwise comparisons amongst control, premanifest HD and manifest HD.**

**Supplementary Material Text 1.**

**Supplementary Material Text 2.**

**Supplementary Workbook 1 (**available to view on figshare https://doi.org/10.6084/m9.figshare.30510515).

**Supplementary Workbook 2 (**available to view on figshare https://doi.org/10.6084/m9.figshare.30510515).

