## Supplemental Figure S1 for "24S-Hydroxycholesterol: A potential brain-derived biomarker of Huntington’s Disease"

**Supplementary Figure S1 Enzyme-Assisted derivatisation for sterol analysis (EADSA).** (**A**) The 3β-hydroxy group of oxysterols/sterols is converted by cholesterol oxidase enzyme to a 3-oxo group. This is then reacted with the [$^{2}H_{5}$]GP to give a GP-hydrazone. Native oxo groups similarly react with [$^{2}H_{5}$]GP. This is illustrated for 24S-HC, 3β,7α-diHCA and 7αH,3O-CA. (**B**) Oxysterols/sterols are reacted with [$^{2}H_{0}$]GP in the absence of cholesterol oxidase. Only molecules with a native oxo group will become derivatised.

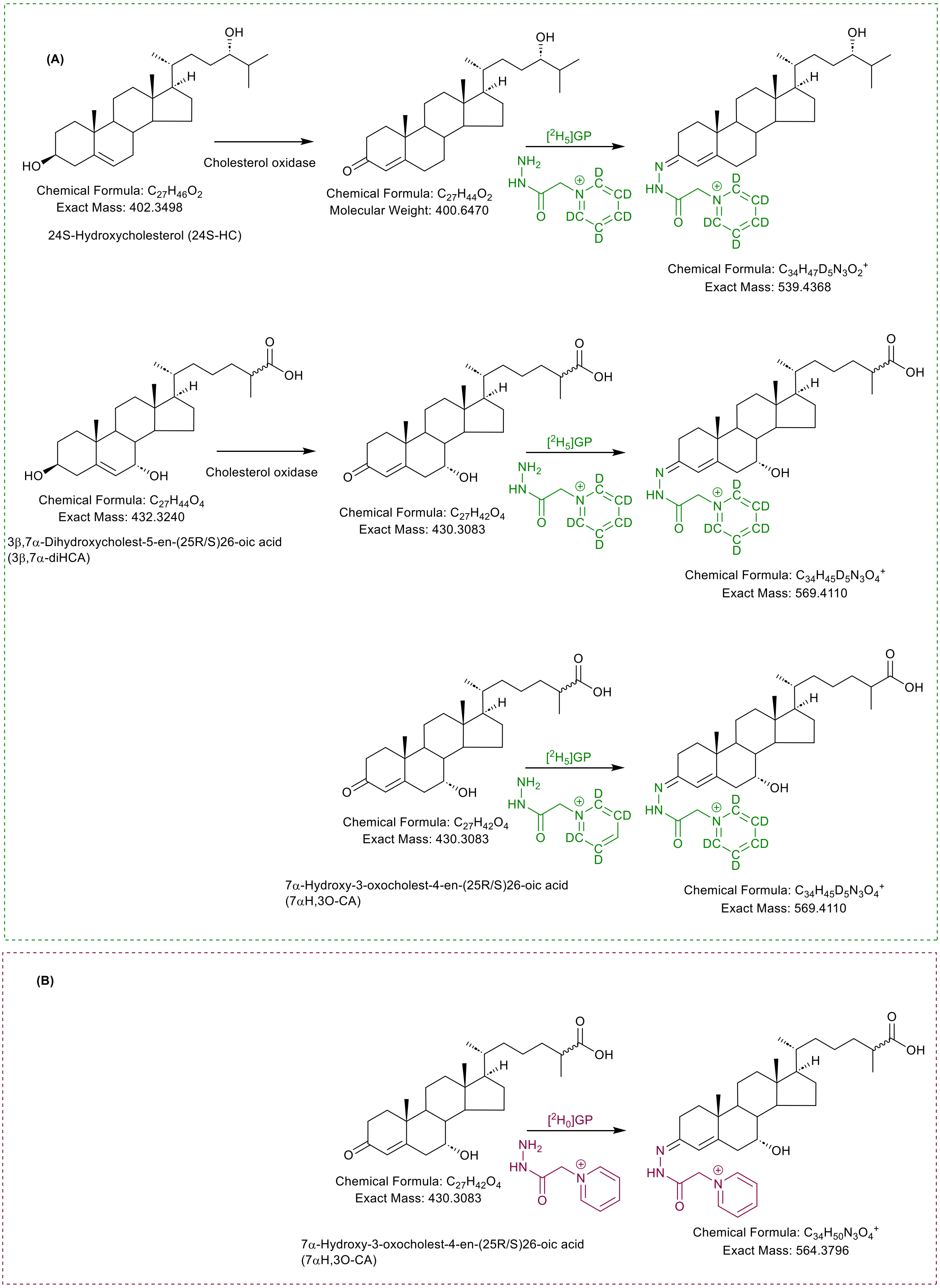
