## Supplemental Figure S2 for "24S-Hydroxycholesterol: A potential brain-derived biomarker of Huntington’s Disease"

**Supplementary Figure S2 Concentrations of oxysterols, cholesterol and cholesterol precursors measured in plasma from healthy controls, people with premanifest HD and manifest HD with values for median, 25$^{th}$ and 75$^{th}$ percentiles.** 400 samples were measured from 388 donors. Each sample is shown as a dot. 6β-Hydroxycholesterol (6β-HC) is derived from the dehydration of cholestane-3β,5α,6β-triol which can be formed via the hydration of 3β-hydroxycholestan-5,6-epoxide. Concentrations for 3β,7α-diHCA and 7αH,3O-CA are given as the sum of 25R- and 25S-epimers. Values for the 25S-epimers alone are given separately. All concentrations are for the non-esterified molecules.

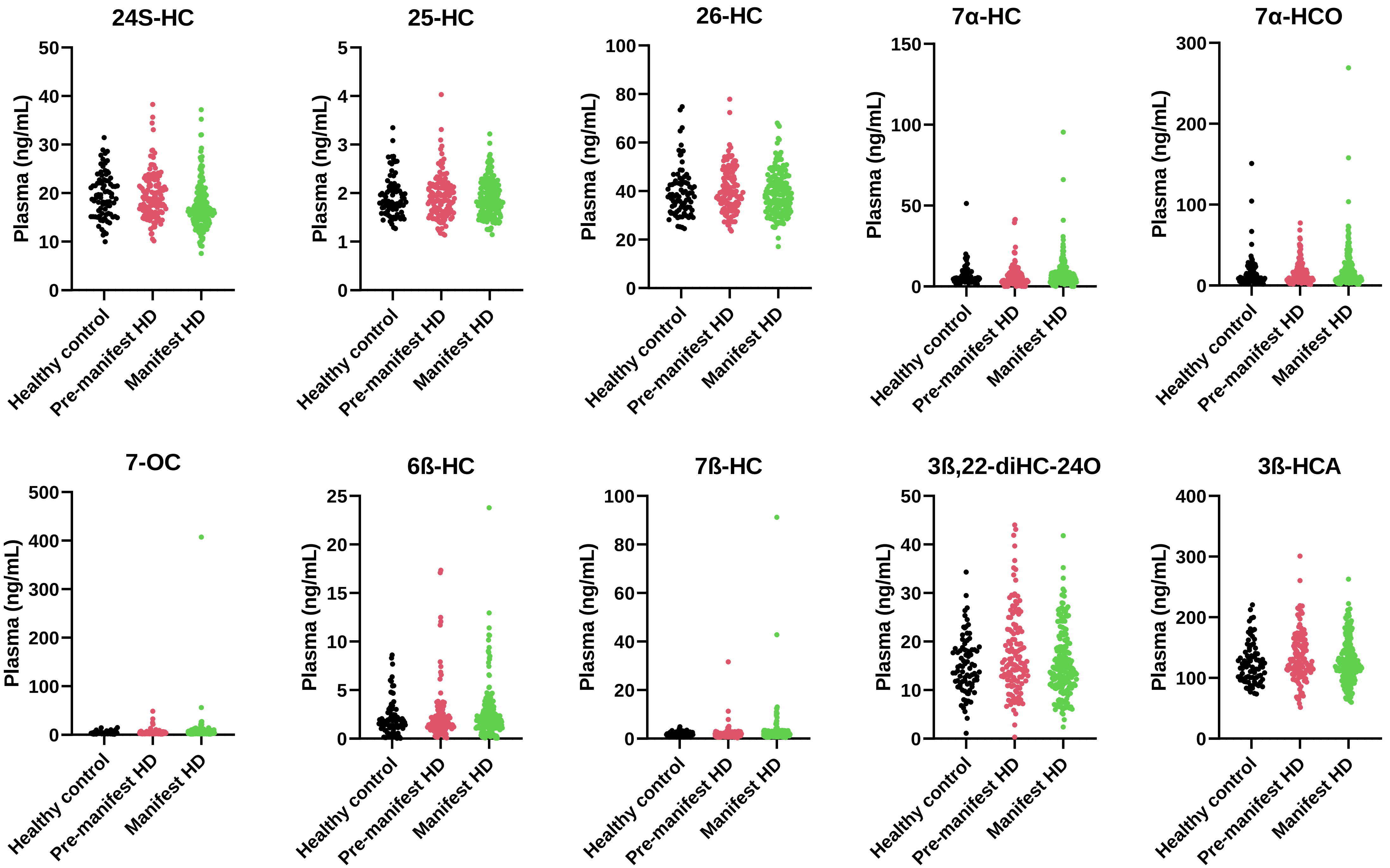
24S-HC
25-HC
26-HC
7α-HC
7α-HCO
7-OC
6ß-HC
7ß-HC
3ß,22-diHC-24O
3ß-HCA
Plasma (ng/mL)
Healthy control
Pre-manifest HD
Manifest HD

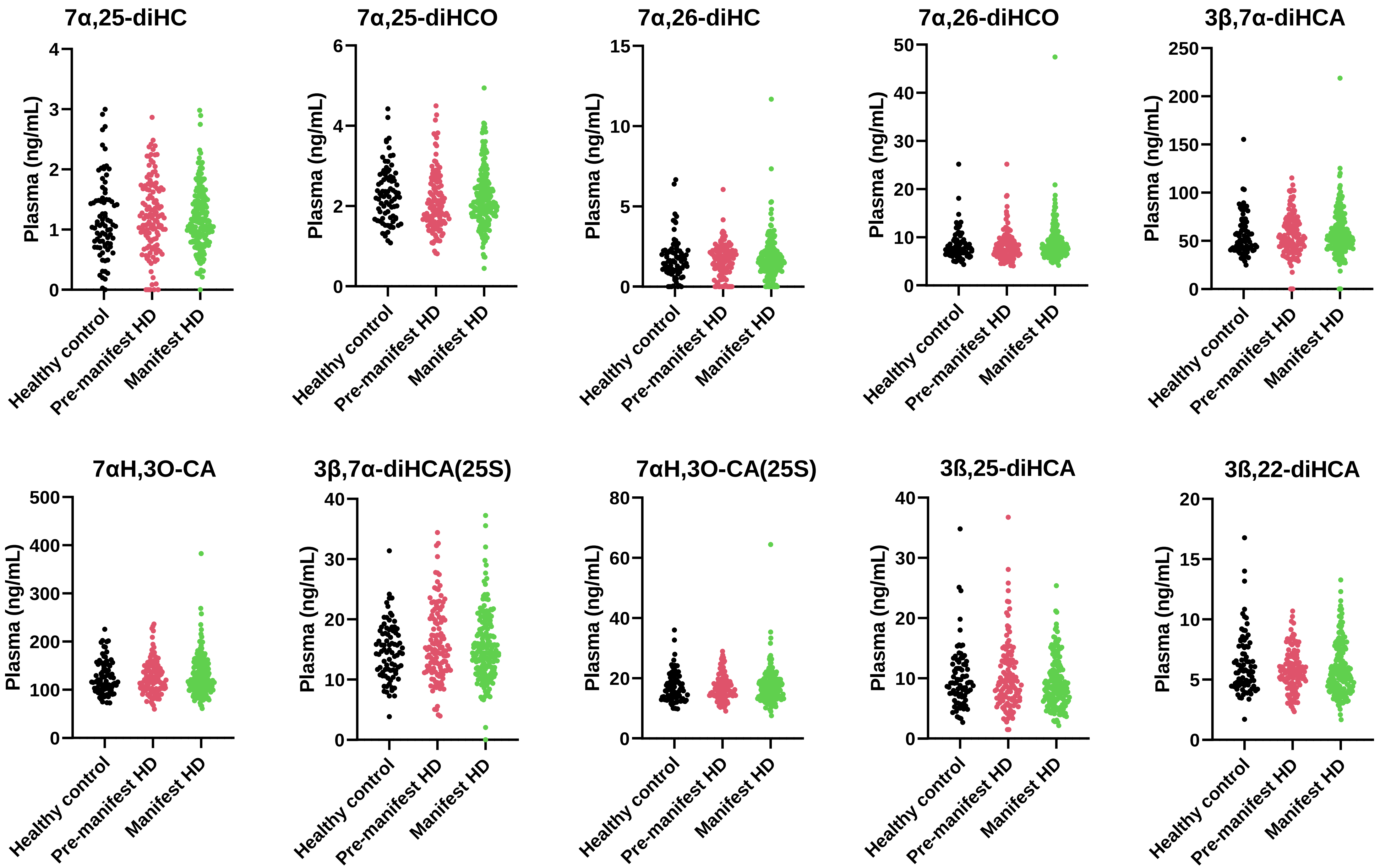

7α,25-diHC
7α,25-diHCO
7α,26-diHC
7α,26-diHCO
3β,7α-diHCA
7αH,3O-CA
3β,7α-diHCA(25S)
7αH,3O-CA(25S)
3ß,25-diHCA
3ß,22-diHCA
Plasma (ng/mL)
Healthy control
Pre-manifest HD
Manifest HD

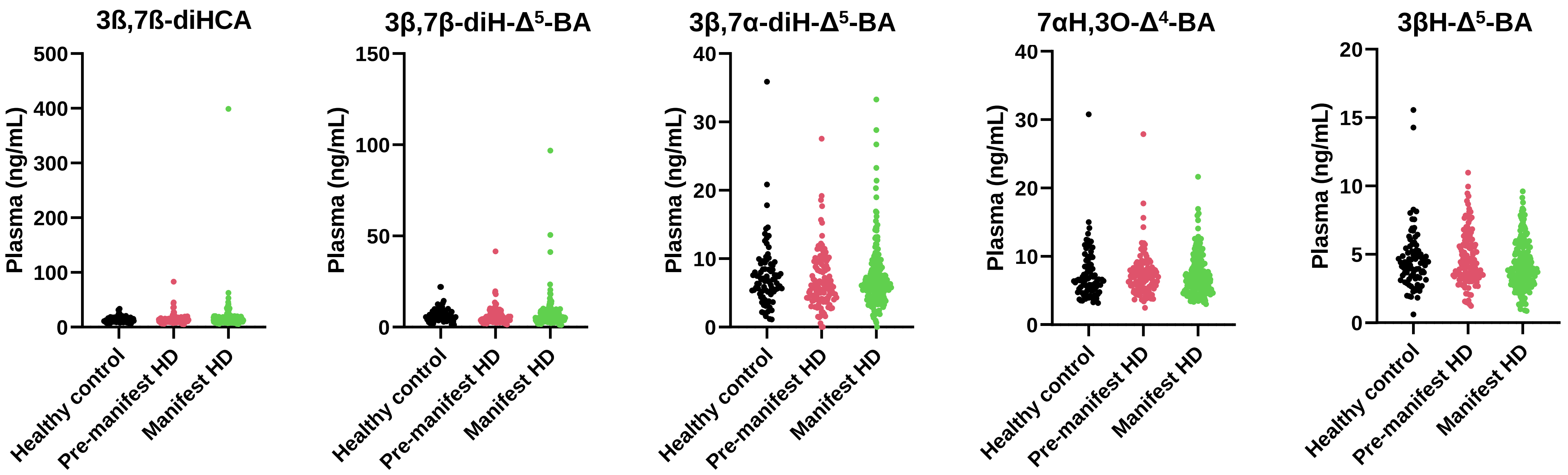
3ß,7ß-diHCA
3β,7β-diH-Δ⁵-BA
3β,7α-diH-Δ⁵-BA
7αH,3O-Δ⁴-BA
3βH-Δ⁵-BA
Plasma (ng/mL)
Healthy control
Pre-manifest HD
Manifest HD

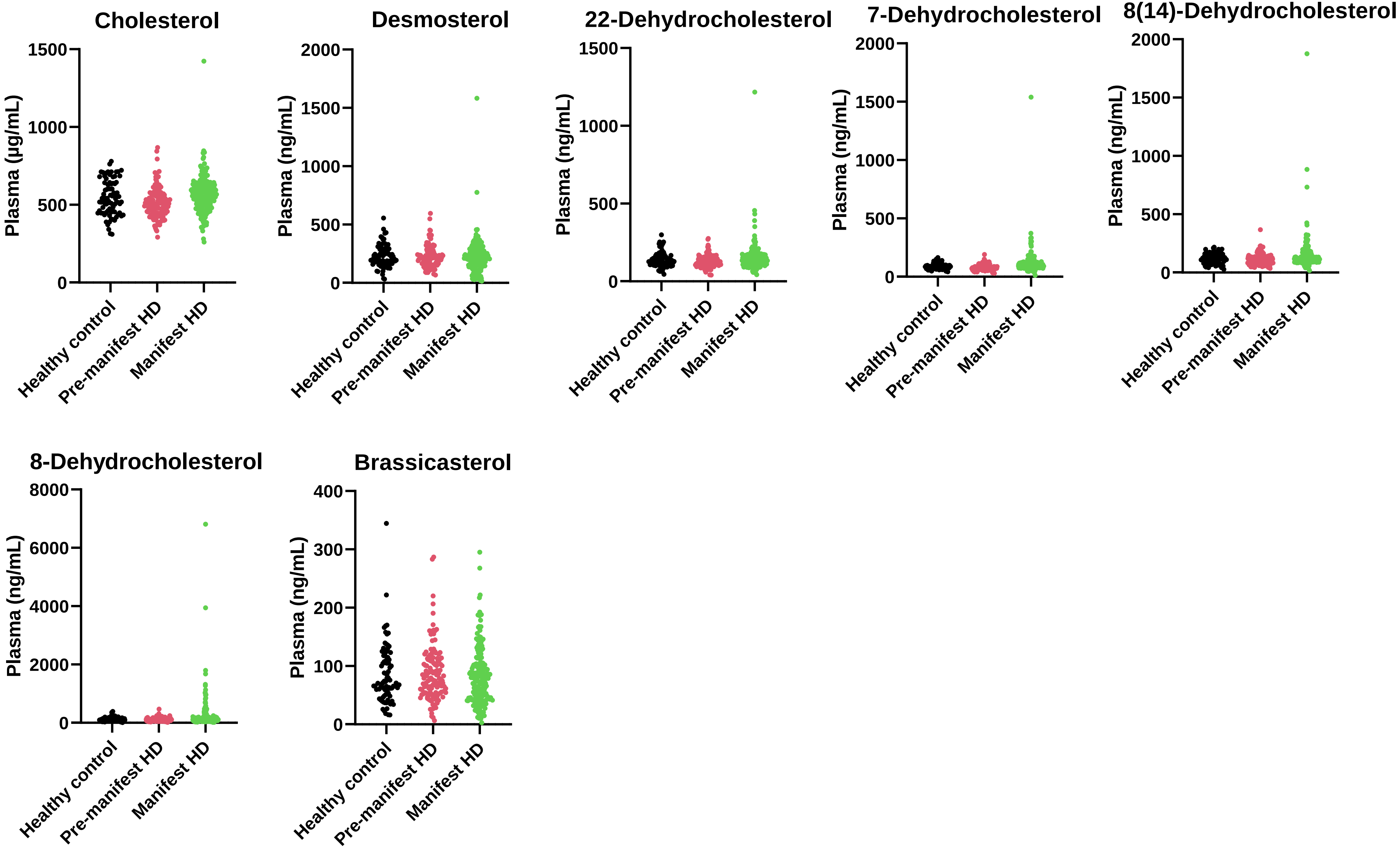

Cholesterol
Plasma (µg/mL)
0
500
1000
1500
Desmosterol
Plasma (ng/mL)
0
500
1000
1500
2000
22-Dehydrocholesterol
Plasma (ng/mL)
0
500
1000
1500
7-Dehydrocholesterol
Plasma (ng/mL)
0
500
1000
1500
2000
8(14)-Dehydrocholesterol
Plasma (ng/mL)
0
500
1000
1500
2000
8-Dehydrocholesterol
Plasma (ng/mL)
0
2000
4000
6000
8000
Brassicasterol
Plasma (ng/mL)
0
100
200
300
400
Healthy control
Pre-manifest HD
Manifest HD

**Values for median, $25^{th}$ and $75^{th}$ percentile oxysterol/sterol concentrations measured in plasma from healthy controls, people with premanifest HD and manifest HD**

| | Control (n = 83) | | | Premanifest HD (n = 129) | | | Manifest HD (n = 188) | | | Isotope labelled standard |
|---|---|---|---|---|---|---|---|---|---|---|
| | Concentration (ng/mL) | | | | | | | | | |
| **Compound Name** | **$25^{th}$** | **Median** | **$75^{th}$** | **$25^{th}$** | **Median** | **$75^{th}$** | **$25^{th}$** | **Median** | **$75^{th}$** | |
| 24S-HC | 15.57 | 19.24 | 22.55 | 16.11 | 18.81 | 22.67 | 14.37 | 16.44 | 19.40 | [26,26,26,27,27,27-$^{2}H_{6}$]24R/S-HC |
| 25-HC | 1.67 | 1.82 | 2.11 | 1.60 | 1.87 | 2.15 | 1.59 | 1.84 | 2.15 | [26,26,26,27,27,27-$^{2}H_{6}$]24R/S-HC |
| 26-HC | 32.64 | 38.48 | 43.74 | 34.41 | 39.49 | 47.44 | 32.61 | 39.15 | 46.05 | [26,26,26,27,27,27-$^{2}H_{6}$]24R/S-HC |
| 7α-HC | 3.33 | 4.98 | 8.60 | 2.65 | 4.59 | 8.11 | 3.16 | 5.49 | 8.97 | [25,26,26,26,27,27,27-$^{2}H_{7}$]7α-HC |
| 7α-HCO | 6.04 | 9.31 | 21.51 | 6.12 | 10.68 | 20.24 | 7.52 | 12.72 | 24.88 | [25,26,26,26,27,27,27-$^{2}H_{7}$]7α-HC[a] |
| 7-OC | 2.51 | 3.37 | 4.31 | 2.48 | 3.21 | 4.21 | 2.77 | 3.62 | 5.44 | [25,26,26,26,27,27,27-$^{2}H_{7}$-]7-OC |
| 6β-HC[b] | 1.12 | 1.75 | 2.48 | 1.07 | 1.61 | 2.52 | 1.19 | 1.91 | 3.14 | [25,26,26,26,27,27,27-$^{2}H_{7}$]7α-HC |
| 7β-HC | 2.46 | 1.66 | 4.46 | 1.03 | 1.51 | 2.21 | 1.27 | 1.91 | 2.68 | [25,26,26,26,27,27,27-$^{2}H_{7}$]7α-HC |
| 3β,22-diHC-24O[c] | 11.22 | 14.49 | 18.31 | 12.04 | 15.88 | 22.63 | 11.01 | 14.23 | 18.92 | [26,26,26,27,27,27-$^{2}H_{6}$]24R/S-HC |
| 3β-HCA | 100.60 | 119.80 | 140.70 | 110.10 | 127.20 | 159.70 | 100.50 | 122.00 | 146.40 | [24,24,27,27,27-$^{2}H_{5}$]3β-HCA |
| 7α,25-diHC | 0.77 | 1.06 | 1.54 | 0.77 | 1.16 | 1.64 | 0.87 | 1.16 | 1.45 | [26,26,26,27,27,27-$^{2}H_{6}$]7α,25-diHC |
| 7α,25-diHCO | 1.74 | 2.22 | 2.70 | 1.64 | 2.03 | 2.61 | 1.74 | 2.03 | 2.51 | [26,26,26,27,27,27-$^{2}H_{6}$]7α,25-diHC[d] |
| 7α,26-diHC | 0.97 | 1.54 | 2.22 | 0.97 | 1.74 | 2.22 | 1.06 | 1.54 | 2.22 | [26,26,26,27,27,27-$^{2}H_{6}$]7α,25-diHC |
| 7α,26-diHCO | 6.66 | 7.62 | 9.07 | 6.27 | 7.33 | 9.07 | 6.85 | 7.91 | 9.75 | [26,26,26,27,27,27-$^{2}H_{6}$]7α,25-diHC[d] |
| 3β,7α-diHCA | 40.88 | 49.83 | 65.23 | 41.91 | 52.62 | 69.30 | 43.14 | 52.58 | 67.59 | [27,27,27-$^{2}H_{3}$]7αH,3O-CA |
| 7αH,3O-CA | 100.60 | 117.60 | 152.20 | 102.50 | 118.50 | 141.90 | 102.40 | 119.30 | 152.40 | [27,27,27-$^{2}H_{3}$]7αH,3O-CA |
| 3β,7α-diHCA(25S) | 11.50 | 14.52 | 18.07 | 11.59 | 15.00 | 20.12 | 11.61 | 14.44 | 18.55 | [27,27,27-$^{2}H_{3}$]7αH,3O-CA |
| 7αH,3O-CA(25S) | 13.60 | 16.24 | 19.58 | 14.17 | 16.19 | 19.34 | 13.82 | 16.54 | 19.41 | [27,27,27-$^{2}H_{3}$]7αH,3O-CA |
| 3β,25-diHCA[e] | 7.03 | 9.21 | 12.31 | 6.41 | 8.87 | 12.92 | 6.05 | 8.31 | 11.39 | [26,26,26,27,27,27-$^{2}H_{6}$]24R/S-HC |
| 3β,22-diHCA[f] | 4.53 | 5.72 | 7.76 | 4.64 | 5.61 | 6.73 | 4.23 | 5.43 | 7.38 | [26,26,26,27,27,27-$^{2}H_{6}$]24R/S-HC |
| 3β,7β-diHCA | 9.86 | 11.38 | 14.73 | 10.39 | 12.10 | 14.93 | 10.34 | 13.05 | 16.75 | [27,27,27-$^{2}H_{3}$]7αH,3O-CA |
| 3β,7β-diH-$\Delta^{5}$-BA | 4.30 | 5.79 | 7.56 | 3.66 | 4.68 | 6.71 | 4.00 | 5.29 | 7.22 | [27,27,27-$^{2}H_{3}$]7αH,3O-CA |
| 3β,7α-diH-$\Delta^{5}$-BA | 5.14 | 6.96 | 9.20 | 4.08 | 5.78 | 8.81 | 4.39 | 6.28 | 8.46 | [27,27,27-$^{2}H_{3}$]7αH.3O-CA |
| 7αH,3O-$\Delta^{4}$-BA | 4.99 | 6.49 | 8.92 | 5.32 | 7.00 | 8.32 | 4.83 | 6.62 | 8.82 | [27,27,27-$^{2}H_{3}$]7αH,3O-CA |
| 3βH-$\Delta^{5}$-BA | 3.22 | 4.36 | 5.31 | 3.30 | 4.27 | 6.05 | 3.01 | 3.94 | 5.40 | [26,26,26,27,27,27-$^{2}H_{6}$]24R/S-HC |
| Desmosterol | 168.00 | 206.90 | 278.20 | 155.40 | 213.70 | 269.80 | 146.80 | 217.50 | 270.90 | [26,26,26,27,27,27-$^{2}H_{6}$]Desmosterol |
| 22-Dehydrocholesterol | 110.60 | 136.20 | 162.80 | 102.70 | 127.20 | 147.80 | 115.00 | 136.50 | 169.20 | [26,26,26,27,27,27-$^{2}H_{6}$]Desmosterol |
| 7-Dehydrocholesterol | 69.87 | 89.56 | 106.60 | 68.29 | 82.70 | 95.01 | 76.68 | 95.07 | 127.20 | [26,26,26,27,27,27-$^{2}H_{6}$]Desmosterol |
| 8(14)-Dehydrocholesterol | 81.56 | 113.80 | 151.00 | 80.41 | 98.64 | 129.90 | 94.56 | 119.90 | 157.20 | [26,26,26,27,27,27-$^{2}H_{6}$]Desmosterol |
| 8-Dehydrocholesterol | 72.74 | 101.90 | 138.20 | 72.38 | 93.79 | 122.40 | 82.02 | 118.20 | 175.10 | [26,26,26,27,27,27-$^{2}H_{6}$]Desmosterol |
| Brassicasterol | 50.86 | 70.95 | 114.90 | 54.60 | 79.44 | 112.90 | 43.33 | 73.75 | 103.60 | [26,26,26,27,27,27-$^{2}H_{6}$]Desmosterol |
| | Concentration (µg/mL) | | | | | | | | | |
| Cholesterol | 452.50 | 539.50 | 638.30 | 446.00 | 507.90 | 568.20 | 489.60 | 567.20 | 625.80 | [25,26,26,27,27,27-$^{2}H_{7}$]Cholesterol |

[a] [25,26,26,26,27,27,27-$^{2}H_{7}$]7α-HC oxidised to [25,26,26,26,27,27,27- $^{2}H_{7}$]7α-HCO internal standard and [25,26,26,26,27,27,27-$^{2}H_{7}$ ]22S-HCO used for any A/B volume correction.

[b] 6β-HC is derived from the dehydration of cholestan-3β,5α,6β-triol which can be formed via the hydration of 3β-hydroxycholestan-5,6-epoxide.

[c] Authentic standard not available. Identification based on exact mass and $MS^3$ spectrum. Possible alternative structure 3β,20-dhydroxycholest-5-en-22-one.

[d] [26,26,26,27,27,27-$^{2}H_{6}$]7α,25-diHC oxidised to [26,26,26,27,27,27-$^{2}H_{7}$]7α,25-diHCO internal standard and [27,27,27-$^{2}H_{3}$]7αH,3O-CA used for any A/B volume correction.

[e] Authentic standard not available. Identification based on exact mass and $MS^3$ spectrum.

[f] Authentic standard not available. Identification based on exact mass and $MS^3$ spectrum. Possible alternative structure 3β,27-dihydroxycholest-5-en-26-oic acid.
