## Supplemental Figure S3 for "24S-Hydroxycholesterol: A potential brain-derived biomarker of Huntington’s Disease"

**Supplementary Figure S3 Model considering age as a potential confounder showing on a natural logarithm scale that mean levels of cholesterol in plasma do not differ between manifest HD and the other two groups.** However, there is a strong statin effect on cholesterol levels and that at a young age cholesterol levels for manifest HD are higher than for controls but at older ages the reverse is true. The concentrations are for the non-esterified molecules.

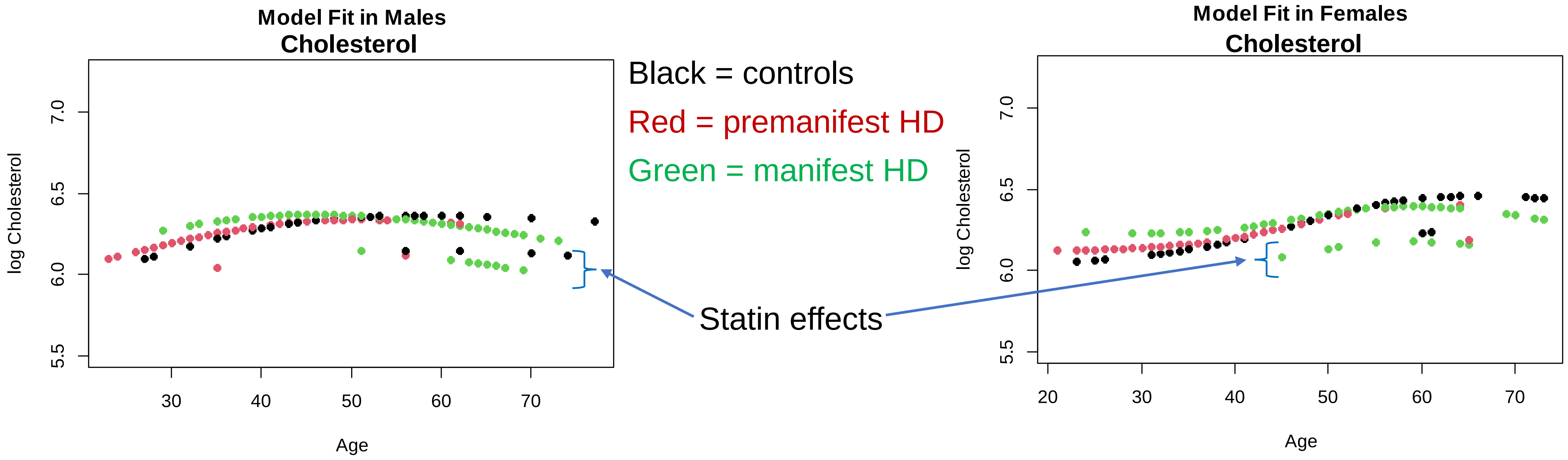

Model Fit in Males
Cholesterol
log Cholesterol
7.0
6.5
6.0
5.5
30
40
50
60
70
Age
Black = controls
Red = premanifest HD
Green = manifest HD
Statin effects
Model Fit in Females
Cholesterol
log Cholesterol
7.0
6.5
6.0
5.5
20
30
40
50
60
70
Age
