## Supplemental Figure S4 for "24S-Hydroxycholesterol: A potential brain-derived biomarker of Huntington’s Disease"

**Supplementary Figure S4 Concentrations of oxysterols, cholesterol and cholesterol precursors measured in CSF from healthy controls, people with premanifest HD and manifest HD with values for median, 25$^{th}$ and 75$^{th}$ percentiles.** 400 samples were measured from 388 donors. The concentrations were determined following a saponification step to hydrolyse sterol esters. Each sample is shown as a dot. 6β-HC is derived from the dehydration of cholestan-3β,5α,6β-triol which can be formed via the hydration of 3β-hydroxycholestan-5,6-epoxide. 6-Dehydrocholesterol is a potential autoxidation product. 7αH,3O-CA was measured as the dehydrated molecule.

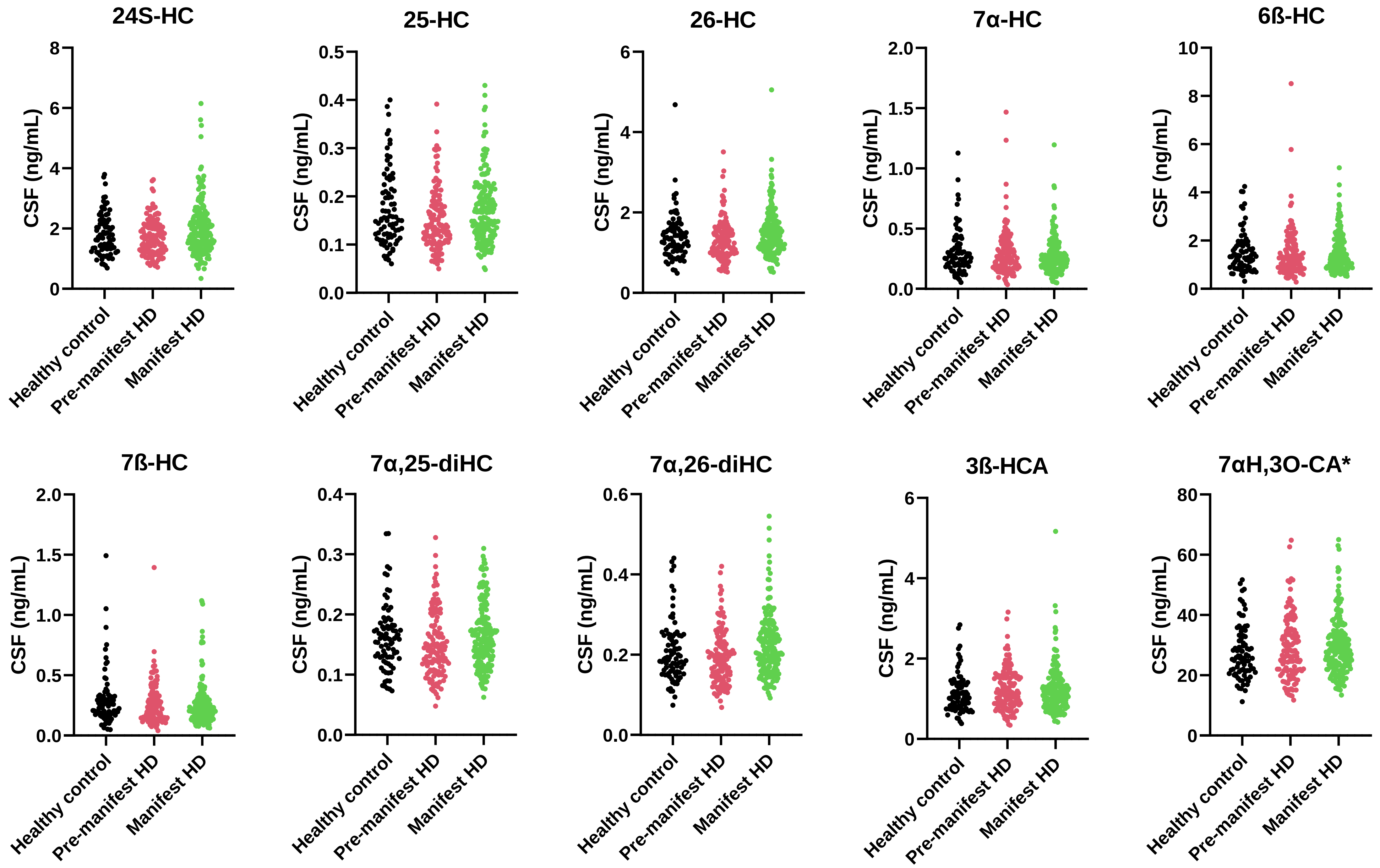
24S-HC
25-HC
26-HC
7α-HC
6ß-HC
7ß-HC
7α,25-diHC
7α,26-diHC
3ß-HCA
7αH,3O-CA*
CSF (ng/mL)
Healthy control
Pre-manifest HD
Manifest HD

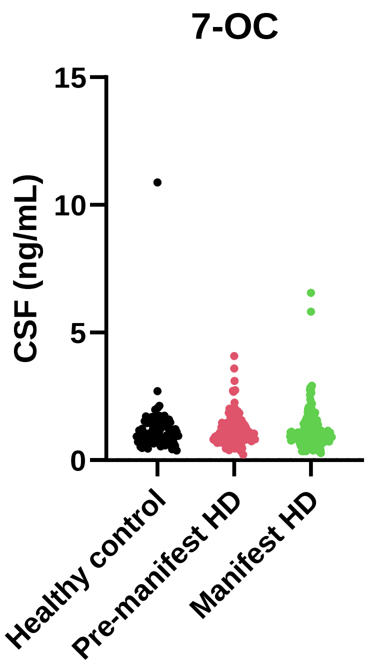
7-OC
CSF (ng/mL)
15
10
5
0
Healthy control
Pre-manifest HD
Manifest HD

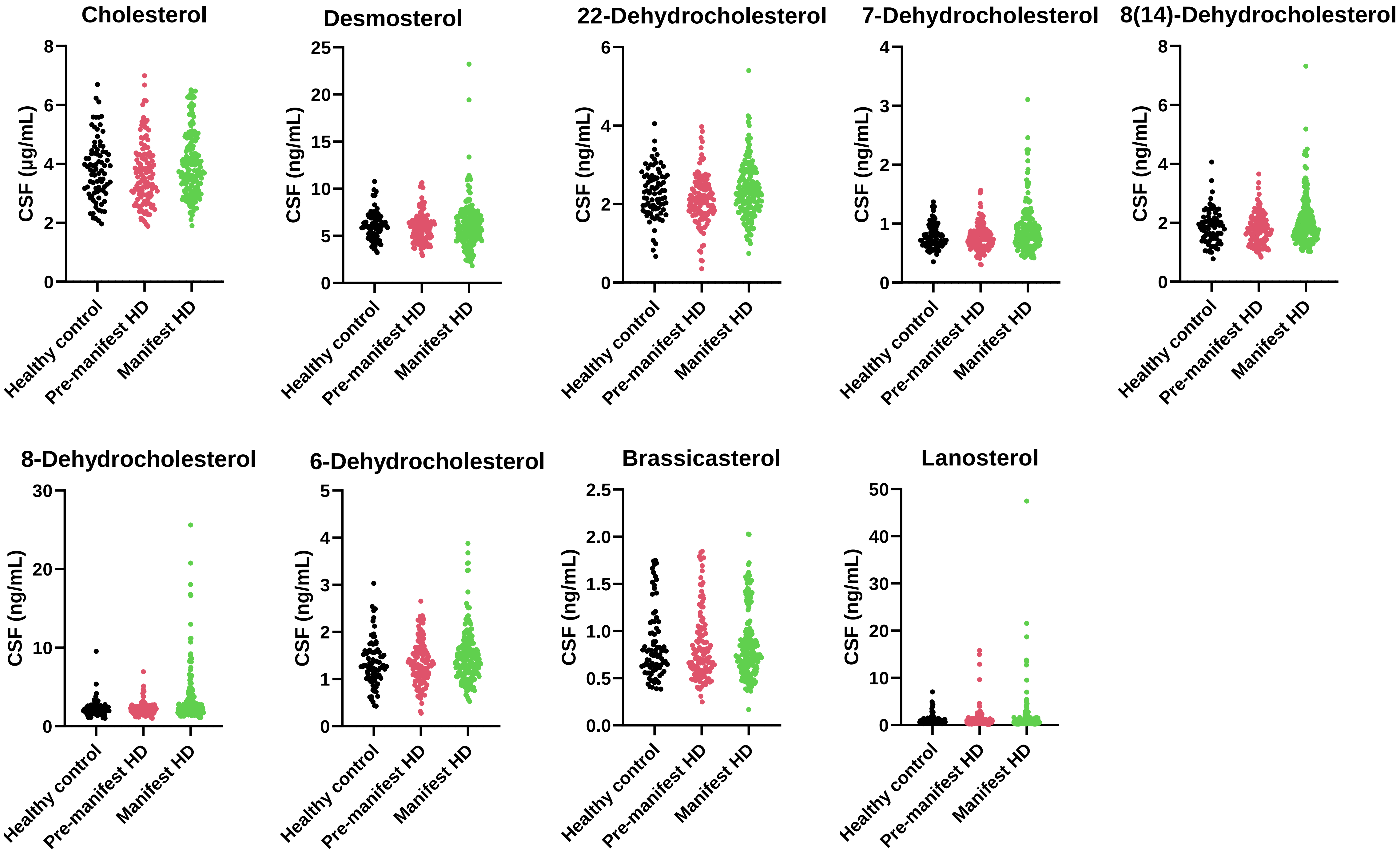

Cholesterol
CSF (µg/mL)
8
6
4
2
0
Desmosterol
CSF (ng/mL)
25
20
15
10
5
0
22-Dehydrocholesterol
CSF (ng/mL)
6
4
2
0
7-Dehydrocholesterol
CSF (ng/mL)
4
3
2
1
0
8(14)-Dehydrocholesterol
CSF (ng/mL)
8
6
4
2
0
8-Dehydrocholesterol
CSF (ng/mL)
30
20
10
0
6-Dehydrocholesterol
CSF (ng/mL)
5
4
3
2
1
0
Brassicasterol
CSF (ng/mL)
2.5
2.0
1.5
1.0
0.5
0.0
Lanosterol
CSF (ng/mL)
50
40
30
20
10
0
Healthy control
Pre-manifest HD
Manifest HD

Values for median, 25th and 75th percentile oxysterol/sterol concentrations measured in CSF from healthy controls, people with premanifest HD and manifest HD.

| | Control (n = 83) | | | Premanifest HD (n = 129) | | | Manifest HD (n = 188) | | | Isotope labelled standard |
|---|---|---|---|---|---|---|---|---|---|---|
| | Concentration (ng/mL) | | | | | | | | | |
| **Compound Name** | **25th** | **Median** | **75th** | **25th** | **Median** | **75th** | **25th** | **Median** | **75th** | |
| 24S-HC | 1.33 | 1.70 | 2.26 | 1.23 | 1.65 | 2.09 | 1.44 | 1.84 | 2.45 | [26,26,26,27,27,27-$^{2}H_{6}$]24R/S-HC |
| 25-HC | 0.12 | 0.15 | 0.22 | 0.11 | 0.14 | 0.18 | 0.12 | 0.17 | 0.21 | [26,26,26,27,27,27-$^{2}H_{6}$]24R/S-HC |
| 26-HC | 1.04 | 1.35 | 1.62 | 0.98 | 1.23 | 1.57 | 1.10 | 1.41 | 1.74 | [26,26,26,27,27,27-$^{2}H_{6}$]24R/S-HC |
| 7α-HC | 0.19 | 0.27 | 0.38 | 0.17 | 0.25 | 0.37 | 0.19 | 0.25 | 0.34 | [25,26,26,26,27,27,27-$^{2}H_{7}$]7α-HC |
| 6β-HC[a] | 0.93 | 1.38 | 1.88 | 0.85 | 1.24 | 1.83 | 0.92 | 1.24 | 1.88 | [25,26,26,26,27,27,27-$^{2}H_{7}$]7α-HC |
| 7β-HC | 0.17 | 0.24 | 0.32 | 0.14 | 0.21 | 0.34 | 0.15 | 0.21 | 0.29 | [25,26,26,26,27,27,27-$^{2}H_{7}$]7α-HC |
| 7α,25-diHC | 0.13 | 0.16 | 0.18 | 0.11 | 0.14 | 0.18 | 0.13 | 0.02 | 0.02 | [26,26,26,27,27,27-$^{2}H_{6}$]7α,25-diHC |
| 7α,26-diHC | 0.15 | 0.19 | 0.25 | 0.14 | 0.19 | 0.23 | 0.17 | 0.21 | 0.26 | [26,26,26,27,27,27-$^{2}H_{6}$]7α,25-diHC |
| 3β-HCA | 0.76 | 1.04 | 1.35 | 0.86 | 1.14 | 1.55 | 0.84 | 1.13 | 1.43 | [24,24,27,27,27-$^{2}H_{5}$]3β-HCA |
| 7αH,3O-CA[b] | 21.22 | 25.73 | 33.17 | 21.53 | 27.13 | 34.78 | 23.22 | 28.12 | 33.62 | [27,27,27-$^{2}H_{3}$]7αH,3O-CA |
| 7-OC | 0.76 | 0.98 | 1.40 | 0.76 | 1.04 | 1.44 | 0.73 | 0.94 | 1.35 | [25,26,26,26,27,27,27-$^{2}H_{7}$-]7-OC |
| 24-Dehydrocholesterol(Desmosterol) | 4.92 | 5.95 | 6.95 | 4.78 | 5.84 | 6.66 | 4.71 | 5.81 | 7.09 | [26,26,26,27,27,27-$^{2}H_{6}$]Desmosterol |
| 22-Dehydrocholesterol | 1.88 | 2.29 | 2.70 | 1.77 | 2.11 | 2.48 | 1.91 | 2.29 | 2.77 | [26,26,26,27,27,27-$^{2}H_{6}$]Desmosterol |
| 7-Dehydrocholesterol | 0.65 | 0.74 | 0.93 | 0.62 | 0.73 | 0.87 | 0.65 | 0.84 | 1.02 | [26,26,26,27,27,27-$^{2}H_{6}$]Desmosterol |
| 8(14)-Dehydrocholesterol | 1.46 | 1.84 | 2.17 | 1.38 | 1.75 | 2.11 | 1.52 | 1.85 | 2.37 | [26,26,26,27,27,27-$^{2}H_{6}$]Desmosterol |
| 8-Dehydrocholesterol | 1.69 | 2.00 | 2.42 | 1.65 | 2.03 | 2.44 | 1.82 | 2.34 | 3.54 | [26,26,26,27,27,27-$^{2}H_{6}$]Desmosterol |
| 6-Dehydrocholesterol[c] | 1.04 | 1.29 | 1.59 | 1.06 | 1.31 | 1.60 | 1.12 | 1.42 | 1.74 | [26,26,26,27,27,27-$^{2}H_{6}$]Desmosterol |
| Brassicasterol | 0.59 | 0.75 | 1.03 | 0.58 | 0.71 | 1.00 | 0.61 | 0.75 | 0.99 | [26,26,26,27,27,27-$^{2}H_{6}$]Desmosterol |
| Lanosterol | 0.42 | 0.65 | 1.14 | 0.44 | 0.62 | 1.18 | 0.43 | 0.65 | 1.32 | [26,26,26,27,27,27-$^{2}H_{6}$]Lanosterol |
| | Concentration (μg/mL) | | | | | | | | | |
| Cholesterol | 3.06 | 3.77 | 4.41 | 2.96 | 3.58 | 4.30 | 3.23 | 3.88 | 4.69 | [25,26,26,27,27,27-$^{2}H_{7}$]Cholesterol |

[a]6β-HC is derived from the dehydration of cholestan-3β,5α,6β-triol which can be formed via the hydration of 3β-hydroxycholestan-5,6-epoxide.

[b]Measured as the dehydrated molecule.

[c]6-Dehydrocholesterol is a potential autoxidation product.
