## Supplemental Figure S5 for "24S-Hydroxycholesterol: A potential brain-derived biomarker of Huntington’s Disease"

**Supplementary Figure S5 ROC curves for the premanifest HD v manifest HD comparison determined by the different classification methods using the full data set.** (**A**) Logistic Regression Forward. (**B**) Logistic Regression Backward. (**C**) Lasso. (**D**) Ridge. (**E**) Elastic Net. (**F**) Random Forest. (**G**) Gradient Boosting.

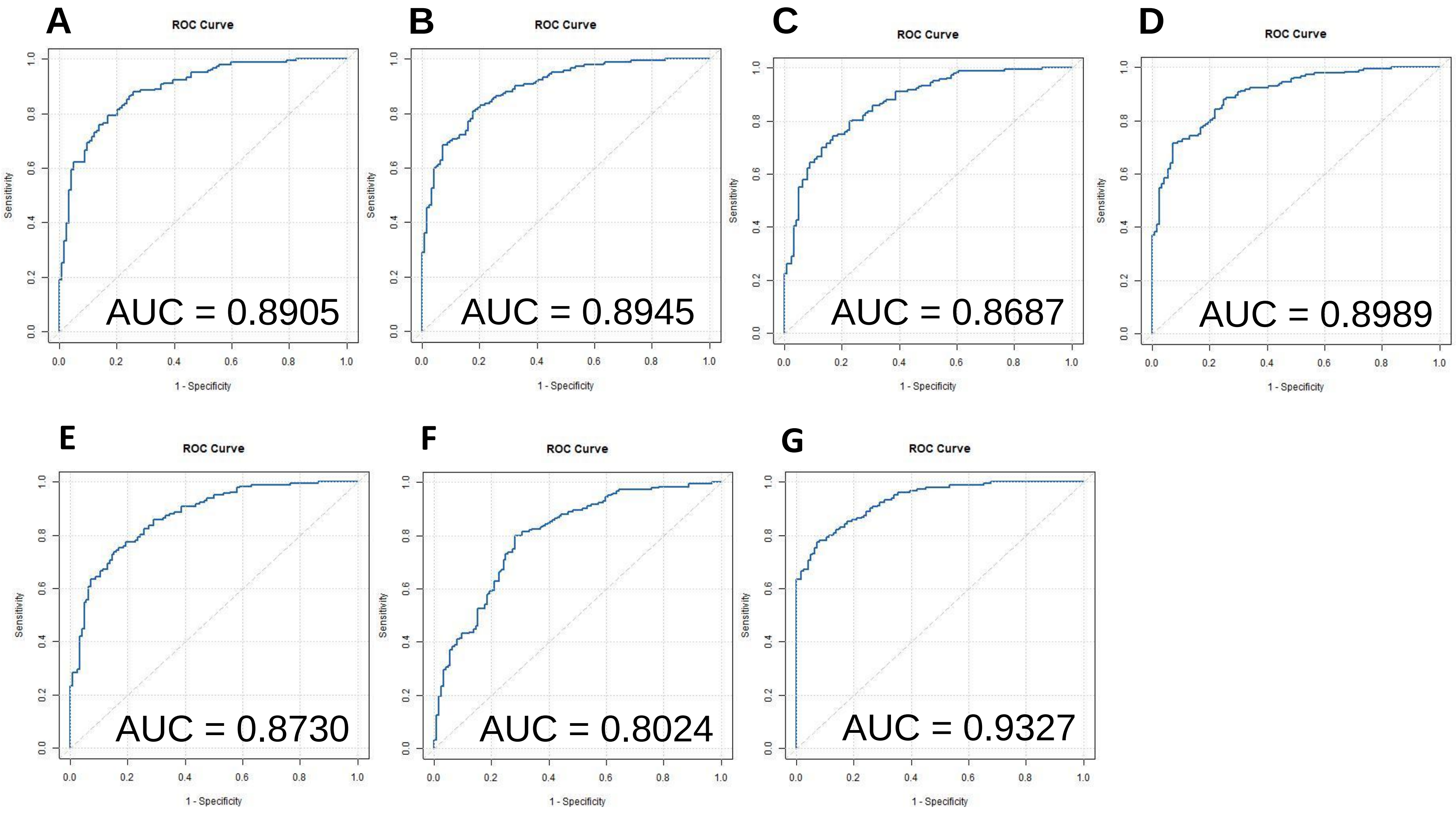
