## Supplemental Table S1 for "24S-Hydroxycholesterol: A potential brain-derived biomarker of Huntington’s Disease"

**Supplementary Table S1 Isotope-labelled standards used for sterol and oxysterol quantification**

| Standard | Abbreviation | Quantity added ng/100 µL plasma | Quantity added ng/100 µL CSF |
|---|---|---|---|
| [26,26,26,27,27,27-$^{2}H_{6}$]24R/S-Hydroxycholesterol | [$^{2}H_{6}$]24R/S-HC | 10.00[a] | 2.00[a] |
| [25,26,26,26,27,27,27-$^{2}H_{7}$]7α-Hydroxycholesterol | [$^{2}H_{7}$]7α-HC | 10.00[a] | 1.00[a] |
| [25,26,26,26,27,27,27-$^{2}H_{7}$]7-Oxocholesterol | [$^{2}H_{7}$]7-OC | 10.00[a] | 1.00[a] |
| [25,26,26,26,27,27,27-$^{2}H_{7}$]22S-Hydroxycholest-4-en-3-one | [$^{2}H_{7}$]22S-HCO | 5.00[b] | 1.00[b] |
| [26,26,26,27,27,27-$^{2}H_{6}$]7α,25-Dihydroxycholesterol | [$^{2}H_{6}$]7α,25-diHC | 4.82[c] | 0.96[c] |
| [24,24,27,27,27-$^{2}H_{5}$]3β-Hydroxycholest-5-en-(25R)26-oic acid | [$^{2}H_{5}$]3β-HCA | 10.00[d] | 1.00[d] |
| [27,27,27-$^{2}H_{3}$]7α-Hydroxy-3-oxocholest-4-en-(25R/S)26-oic acid | [$^{2}H_{3}$]7αH,3O-CA | 10.00[e] | 2.00[e] |
| [26,26,26,27,27,27-$^{2}H_{6}$]Desmosterol | -- | 60.00[f] | 3.00[f] |
| [26,26,26,27,27,27-$^{2}H_{6}$]Lanosterol | -- | -- | 6.00 |
| [25,26,26,26,27,27,27-$^{2}H_{7}$]Cholesterol | -- | 32,280[h] | 322.80[h] |

[a] Avanti Polar Lipids, Lipid Maps MS quantitative standard.

[b] Prepared from [25,26,26,26,27,27,27-$^{2}H_{7}$]22S-hydroxycholesterol, Avanti Polar Lipids standard, nominal weight.

[c] Avanti Polar Lipids standard, quantified against Avanti Polar Lipids OxysterolSPLASH (Lot 6670WAA011).

[d] Avanti Polar Lipids standard quantified against (25R)26-HC in Avanti Polar Lipids OxysterolSPLASH (Lot 6670WAA010).

[e] Avanti Polar Lipids standard quantified against Avanti Polar Lipids OxysterolSPLASH (Lot 6670WAA010).

[f] Avanti Polar Lipids standard, quantified against Lipid Maps MS quantitative standard.

[h] Avanti Polar Lipids standard.

[g] Avanti Polar Lipids standard quantified against Lipid Maps MS quantitative standard.
