## Supplemental Table S2 for "24S-Hydroxycholesterol: A potential brain-derived biomarker of Huntington’s Disease"

**Table S2. List of oxysterol/sterol abbreviations**

| | | | | | | | |
|---|---|---|---|---|---|---|---|
| $[^2H_5]$GP-[M]$^+$ *m/z* | **Systematic (common) name** | **Abbreviation** | **Nomenclature used in Binary Classification Techniques** | | **Lipid Maps ID** | **Exact Mass** | **Shorthand Nomenclature**[a] |
| | **Oxysterols** | | **(p)Plasma** | **(c)CSF** | | | |
| 511.3691 | 3β-Hydroxychol-5-en-24-oic acid | 3βH-$\Delta^5$-BA | p3βHΔ5BA | c3βHΔ5BA | LMST04010201 | 374.2821 | ST 24:2;O3 |
| 527.3640 | 3β,7α-Dihydroxychol-5-en-24-oic acid | 3β,7α-diH-$\Delta^5$-BA | p3β7αdiHΔ5BA | c3β7αdiHΔ5BA | LMST04010217 | 390.2770 | ST 24:2;O4 |
| 527.3640 | 3β,7β-Dihydroxychol-5-en-24-oic acid | 3β,7β-diH-$\Delta^5$-BA | p3β7βdiHΔ5BA | c3β7βdiHΔ5BA | LMST04010218 | 390.2770 | ST 24:2;O4 |
| 539.4368 | Cholest-5-ene-3β,24S-diol (24S-Hydroxycholesterol) | 24S-HC | p24SHC | c24SHC | LMST01010019 | 402.3498 | ST 27:1;O2 |
| 539.4368 | Cholest-5-ene-3β,25-diol (25-Hydroxycholesterol) | 25-HC | p25HC | c25HC | LMST01010018 | 402.3498 | ST 27:1;O2 |
| 539.4368 | Cholest-5-ene-3β,(25R)26-diol ((25R)26-Hydroxycholesterol, 27-Hydroxycholesterol) | 26-HC | p27HC | c27HC | LMST01010088 | 402.3498 | ST 27:1;O2 |
| 539.4368 | Cholest-5-ene-3β,7α-diol (7α-Hydroxycholesterol) | 7α-HC | p7αHC | c7αHC | LMST01010013 | 402.3498 | ST 27:1;O2 |
| 539.4368 | Cholest-5-ene-3β,7β-diol (7β-Hydroxycholesterol) | 7β-HC | p7βHC | c7βHC | LMST01010047 | 402.3498 | ST 27:1;O2 |
| 539.4368 | Cholest-4-ene-3β,6β-diol[b] (6β-Hydroxycholesterol) | 6β-HC | p6βHC | c6βHC | NA | 402.3498 | ST 27:1;O2 |
| 539.4368 | Cholest-5-ene-3β,19-diol (19-Hydroxycholesterol) | 19-HC | | | LMST01010274 | 402.3498 | ST 27:1;O2 |
| 553.4161 | 3β,22-Dihydroxycholest-5-en-24-one[c] | 3β,22-diHC-24O | p3β22diHC24O | c3β22diHC24O | NA | 416.3290 | ST 27:2;O3 |
| 553.4161 | 3β-Hydroxycholest-5-en-(25R)26-oic acid (Cholestenoic acid) | 3β-HCA | p3βHCA | c3βHCA | LMST04030072 | 416.3290 | ST 27:2;O3 |
| 555.4317 | Cholest-5-ene-3β,7α,25-triol (7α,25-Dihydroxycholesterol) | 7α,25-diHC | p7α25diHC | c7α25diHC | LMST04030166 | 418.3447 | ST 27:1;O3 |
| 555.4317 | Cholest-5-ene-3β,7α,(25R/S)26-triol (7α,(25R/S)26-Dihydroxycholesterol, 7α,27-Dihydroxycholesterol) | 7α,26-diHC | p7α26diHC | c7α26diHC | LMST04030081 | 418.3447 | ST 27:1;O3 |
| 569.4110 | 3β,25-Dihydroxycholest-5-en-26-oic acid[d] | 3β,25-diHCA | p3β25diHCA | c3β25diHCA | NA | 432.3240 | ST 27:2;O4 |
| 569.4110 | 3β,22-Dihydroxycholest-5-en-26-oic acid[e] | 3β,22-diHCA | p3β22-diHCA | c3β,22-diHCA | NA | 432.3240 | ST 27:2;O4 |
| 569.4110 | 3β,7β-Dihydroxycholest-5-en-(25R/S)26-oic acid | 3β,7β-diHCA | p3β7βdiHCA | c3β7βdiHCA | NA | 432.3240 | ST 27:2;O4 |
| 569.4110 | 3β,7α-Dihydroxycholest-5-en-(25S)26-oic acid | 3β,7α-diHCA(25S) | p3β7αdiHCA25S | c3β7αdiHCA25S | NA | 432.3240 | ST 27:2;O4 |
| 569.4110 | 3β,7α-Dihydroxycholest-5-en-(25R/S)26-oic acid | 3β,7α-diHCA(25R/S) | p3β7αdiHCA | c3β7αdiHCA | LMST04030148 | 432.3240 | ST 27:2;O4 |
| | **Sterols** | | | | | | |
| 521.4262 | Cholesta-4,6-dien-3β-ol (6-Dehydrocholesterol)[f] | 6-DHC | p6-Dehydrocholesterol | c6-Dehydrocholesterol | NA | 384.3392 | ST 27:2;O |
| 521.4262 | Cholesta-5,7-dien-3β-ol (7-Dehydrocholesterol) | 7-DHC | p7Dehydrocholesterol | c7Dehydrocholesterol | LMST01010069 | 384.3392 | ST 27:2;O |
| 521.4262 | Cholesta-5,8(9)-dien-3β-ol (8-Dehydrocholesterol) | 8-DHC | p89Dehydrocholesterol | c89Dehydrocholesterol | LMST01010242 | 384.3392 | ST 27:2;O |
| 521.4262 | Cholesta-5,8(14)-dien-3β-ol (8(14)-Dehydrocholesterol) | 8(14)-DHC | p814Dehydrocholesterol | c814Dehydrocholesterol | NA | 384.3392 | ST 27:2;O |
| 521.4262 | Cholesta-5,22-dien-3β-ol (22-Dehydrocholesterol)[d] | 22-DHC | p22Dehydrocholesterol | c22Dehydrocholesterol | LMST01010095 | 384.3392 | ST 27:2;O |

| | | | | | | | |
|---|---|---|---|---|---|---|---|
| 521.4262 | Cholesta-5,24-dien-3β-ol (Desmosterol) | 24-DHC | pDesmosterol | cDesmosterol | LMST01010016 | 384.3392 | ST 27:2;O |
| 535.4419 | 24R-Methylcholesta-5,22-diene-3β-ol (Brassicasterol) | | pBrassicasterol | cBrassicasterol | LMST01030098 | 398.3549 | ST 28:2;O |
| 523.4419 | Cholest-5-en-3β-ol (Cholesterol) | | pCholesterol | cCholesterol | LMST01010001 | 386.3549 | ST 27:1;O |
| **[MH-$H_2O$]$^+$ *m/z*** | | | | | | | |
| 409.3829 | 4,4,14α-Trimethyl-5α-cholesta-8,24-dien-3β-ol (Lanosterol) | | | | LMST01010017 | 426.3862 | ST 30:2;O |
| **[$^2H_0$]GP-[M]$^+$ *m/z*** | **Systematic (common) name** | | | | | | |
| | **Oxysterols** | | | | | | |
| 522.3326 | 7α-Hydroxy-3-oxochol-4-en-24-oic acid | 7αH,3O-$\Delta^4$-BA | p7αH3OΔ4BA | c7αH3OΔ4BA | LMST04010239 | 388.2614 | ST 24:3;O4 |
| 534.4054 | 3β-Hydroxycholest-5-en-7-one (7-Oxocholesterol, 7-Ketocholesterol) | 7-OC (7-KC) | p7OC (pKC) | c7OC (cKC) | LMST01010049 | 400.3341 | ST 27:2;O2 |
| 534.4054 | 7α-Hydroxycholest-4-en-3-one | 7α-HCO | p7αHCO | c7αHCO | LMST04030123 | 400.3341 | ST 27:2;O2 |
| 550.4003 | 7α,25-Dihydroxycholest-4-en-3-one | 7α,25-diHCO | p7α25diHCO | c7α25diHCO | LMST04030107 | 416.3290 | ST 27:2;O3 |
| 550.4003 | 7α,(25R/S)26-Dihydroxycholest-4-en-3-one (7α,27-Dihydroxycholest-4-en-3-one) | 7α,26-diHCO | p7α26diHCO | c7α26diHCO | LMST04030157 | 416.3290 | ST 27:2;O3 |
| 564.3796 | 7α-Hydroxy-3-oxocholest-4-en-(25S)26-oic acid | 7αH,3O-CA(25S) | p7αH3OCA25S | c7αH3OCA25S | NA | 430.3083 | ST 27:3;O4 |
| 564.3796 | 7α-Hydroxy-3-oxocholest-4-en-(25R/S)26-oic acid | 7αH,3O-CA(25R/S) | p7αH3OCA(25R/S) (p7αH,3O-CA) | c7αH3OCA18 | LMST04030149 | 430.3083 | ST 27:3;O4 |
| **[$^2H_5$]GP-[M]$^+$ *m/z*** | **Isotope-labelled standards** | | | | | | |
| 527.4639 | [26,26,26,27,27,27-$^2H_6$]Cholesta-5,24-dien-3β-ol ($^2H_6$]Desmosterol) | | | | LMST01010090 | 390.3769 | |
| 530.4858 | [25,26,26,26,27,27,27-$^2H_7$]Cholest-5-en-3β-ol ([$^2H_7$]Cholesterol) | | | | NA | 393.3988 | |
| 545.4745 | [26,26,26,27,27,27-$^2H_6$]Cholest-5-ene-3β,24R/S-diol ([$^2H_6$]24R/S-Hydroxycholesterol) | [$^2H_6$]24R/S-HC | | | LMST01010092 | 408.3874 | |
| 546.4807 | [25,26,26,26,27,27,27-$^2H_7$]Cholest-5-ene-3β,7α-diol ([$^2H_7$]7α-Hydroxycholesterol) | [$^2H_7$]7α-HC | | | LMST01010006 | 409.3937 | |
| 558.4475 | [24,24,27,27,27-$^2H_5$]3β-Hydroxycholest-5-en-(25R)26-oic acid | [$^2H_5$]CA$^5$-3β-ol | | | NA | 421.3604 | |
| 561.4694 | [26,26,26,27,27,27-$^2H_6$]Cholest-5-ene-3β,7α,25-triol ([$^2H_6$]7α,25-Dihydroxycholesterol) | [$^2H_6$]7α,25-diHC | | | NA | 424.3824 | |
| **[M+H-$H_2O$]$^+$ *m/z*** | **Isotope-labelled standards** | | | | | | |
| 415.4205 | [26,26,26,27,27,27-$^2H_6$]4,4,14α-Trimethyl-5α-cholesta-8,24-dien-3β-ol (Lanosterol) | | | | NA | 432.4238 | |
| **[$^2H_0$]GP-[M]$^+$ *m/z*** | **Isotope-labelled standards** | | | | | | |

| | | | | | | |
|---|---|---|---|---|---|---|
| 541.4493 | [25,26,26,26,27,27,27-$^{2}H_{7}$]22R-Hydroxycholest-4-en-3-one | [$^{2}H_{7}$]22R-HCO | | | NA | 407.3781 |
| 541.4493 | [25,26,26,26,27,27,27-$^{2}H_{7}$]3β-Hydroxycholest-5-en-7-one ([$^{2}H_{7}$]7-Oxocholesterol, [$^{2}H_{7}$]7-ketocholesterol) | [$^{2}H_{7}$]7-OC | | | NA | 407.3781 |
| 567.3984 | [27,27,27-$^{2}H_{3}$]7α-Hydroxy-3-oxocholest-4-en-(25R/S)26-oic acid | [$^{2}H_{3}$]7αH,3O-CA(25R/S) | | | NA | 433.3271 |

[a]Shorthand nomenclature at the sum composition level.
[b]Cholest-4-ene-3β,6β-diol is derived from the dehydration of cholestane-3β,5α,6β-triol which can be formed via the hydration of 3β-hydroxycholestan-5,6-epoxide.
[c]Authentic standard not available. Identification based on exact mass and $MS^3$ spectrum. Possible alternative structure 3β,20-dihydroxycholest-5-en-22-one.
[d]Authentic standard not available. Identification based on exact mass and $MS^3$ spectrum.
[e]Authentic standard not available. Identification based on exact mass and $MS^3$ spectrum. Possible alternative structure 3β,27-dihydroxycholest-5-en-26-oic acid.
[f]Likely autoxidation product.
