## Supplemental Table S3 for "24S-Hydroxycholesterol: A potential brain-derived biomarker of Huntington’s Disease"

**Table S3A Concentrations measured in plasma samples from people taking statins, plasma concentrations are for the "free" sterols/oxysterols**

| | Healthy Controls (n = 8) | | Premanifest HD (n = 5) | | Manifest HD (n = 17) | | QC Replicates (n = 52) | | | NIST SRM 1950 (n = 6) | | | Isotope labelled standard |
|---|---|---|---|---|---|---|---|---|---|---|---|---|---|
| | Concentration (ng/mL) | | | | | | | | | Concentration (ng/mL) | | | |
| **Compound Name** | **MEAN** | **SD** | **MEAN** | **SD** | **MEAN** | **SD** | **MEAN** | **SD** | **%CV** | **MEAN** | **SD** | **%CV** | |
| 24S-HC | 16.37 | 4.79 | 18.27 | 2.52 | 14.86 | 4.57 | 21.38 | 0.90 | 4.21 | 8.94 | 0.47 | 5.26 | [26,26,26,27,27,27-$^2H_6$]24R/S-HC |
| 25-HC | 1.71 | 0.26 | 1.97 | 0.22 | 1.79 | 0.48 | 1.57 | 0.16 | 9.90 | 1.32 | 0.06 | 4.69 | [26,26,26,27,27,27-$^2H_6$]24R/S-HC |
| 26-HC | 35.35 | 7.29 | 36.61 | 2.05 | 35.88 | 10.10 | 39.10 | 2.30 | 5.88 | 22.30 | 0.68 | 3.07 | [26,26,26,27,27,27-$^2H_6$]24R/S-HC |
| 7α-HC | 5.57 | 3.65 | 3.51 | 1.83 | 10.64 | 15.83 | 6.22 | 1.46 | 23.53 | 13.23 | 1.50 | 11.34 | [25,26,26,26,27,27,27-$^2H_7$]7α-HC |
| 7α-HCO | 13.07 | 9.03 | 7.30 | 2.14 | 27.10 | 38.24 | 12.80 | 1.84 | 14.38 | 19.70 | 0.84 | 4.27 | [25,26,26,26,27,27,27-$^2H_7$]7α-HC[a] |
| 7-OC | 4.38 | 1.78 | 3.33 | 0.90 | 6.45 | 6.36 | 3.13 | 0.80 | 25.68 | 6.21 | 1.99 | 31.98 | [25,26,26,26,27,27,27-$^2H_7$-]7-OC |
| 6β-HC[b] | 2.27 | 2.61 | 1.96 | 1.15 | 3.70 | 3.32 | 1.77 | 1.21 | 68.39 | 4.62 | 2.06 | 44.53 | [25,26,26,26,27,27,27-$^2H_7$]7α-HC |
| 7β-HC | 2.07 | 0.73 | 1.52 | 0.57 | 3.22 | 3.54 | 1.35 | 0.43 | 31.64 | 4.24 | 0.62 | 14.50 | [25,26,26,26,27,27,27-$^2H_7$]7α-HC |
| 3β,22-diHC-24O[c] | 12.35 | 5.08 | 10.90 | 3.86 | 13.75 | 5.28 | 12.49 | 1.62 | 12.99 | 13.59 | 0.47 | 3.44 | [26,26,26,27,27,27-$^2H_6$]24R/S-HC |
| 3β-HCA | 123.82 | 43.53 | 142.51 | 28.90 | 131.33 | 53.04 | 114.24 | 4.98 | 4.36 | 101.91 | 4.34 | 4.25 | [24,24,27,27,27-$^2H_5$]3β-HCA |
| 7α,25-diHC | 1.16 | 0.73 | 1.33 | 0.43 | 1.15 | 0.62 | 0.87 | 0.37 | 42.93 | 0.94 | 0.21 | 22.47 | [26,26,26,27,27,27-$^2H_6$]7α,25-diHC |
| 7α,25-diHCO | 2.27 | 0.78 | 2.24 | 0.82 | 2.05 | 0.47 | 1.31 | 0.36 | 27.13 | 1.92 | 0.12 | 6.08 | [26,26,26,27,27,27-$^2H_6$]7α,25-diHC[d] |
| 7α,26-diHC | 1.52 | 0.85 | 2.12 | 0.64 | 1.83 | 1.68 | 1.26 | 0.68 | 53.62 | 0.64 | 0.33 | 52.22 | [26,26,26,27,27,27-$^2H_6$]7α,25-diHC |
| 7α,26-diHCO | 8.19 | 2.26 | 6.81 | 1.65 | 8.46 | 3.25 | 6.70 | 1.08 | 16.18 | 7.58 | 0.72 | 9.54 | [26,26,26,27,27,27-$^2H_6$]7α,25-diHC[d] |
| 3β,7α-diHCA | 53.72 | 18.53 | 58.84 | 20.95 | 57.32 | 28.99 | 48.93 | 4.27 | 8.74 | 46.89 | 6.02 | 12.83 | [27,27,27-$^2H_3$]7αH,3O-CA |
| 7αH,3O-CA | 127.97 | 28.81 | 104.42 | 22.92 | 128.87 | 39.71 | 130.35 | 6.48 | 4.98 | 104.24 | 3.01 | 2.89 | [27,27,27-$^2H_3$]7αH,3O-CA |
| 3β,7α-diHCA(25S) | 14.50 | 5.01 | 14.70 | 5.45 | 16.30 | 6.26 | 13.26 | 1.73 | 13.08 | 13.67 | 1.37 | 10.02 | [27,27,27-$^2H_3$]7αH,3O-CA |
| 7αH,3O-CA(25S) | 17.42 | 3.36 | 14.40 | 3.21 | 17.53 | 5.81 | 16.46 | 1.65 | 10.02 | 14.43 | 0.59 | 4.06 | [27,27,27-$^2H_3$]7αH,3O-CA |
| 3β,25-diHCA[e] | 8.47 | 4.09 | 6.14 | 1.88 | 7.52 | 3.37 | 6.49 | 1.66 | 25.58 | 8.29 | 1.06 | 12.76 | [26,26,26,27,27,27-$^2H_6$]24R/S-HC |
| 3β,22-diHCA[f] | 7.44 | 3.31 | 5.85 | 1.79 | 7.02 | 2.45 | 4.20 | 0.85 | 20.21 | 5.48 | 0.67 | 12.30 | [26,26,26,27,27,27-$^2H_6$]24R/S-HC |
| 3β,7β-diHCA | 14.24 | 3.81 | 11.57 | 1.62 | 21.28 | 14.72 | 10.64 | 1.58 | 14.86 | 10.63 | 0.62 | 5.85 | [27,27,27-$^2H_3$]7αH,3O-CA |
| 3β,7β-diH-$\Delta^5$-BA | 7.13 | 2.23 | 4.15 | 0.71 | 9.76 | 11.35 | 4.38 | 0.82 | 18.62 | 5.83 | 1.14 | 19.51 | [27,27,27-$^2H_3$]7αH,3O-CA |
| 3β,7α-diH-$\Delta^5$-BA | 7.62 | 2.80 | 4.68 | 1.11 | 7.64 | 6.21 | 5.82 | 1.59 | 27.28 | 6.53 | 2.01 | 30.86 | [27,27,27-$^2H_3$]7αH,3O-CA |
| 7αH,3O-$\Delta^4$-BA | 7.26 | 2.39 | 5.38 | 0.99 | 7.55 | 2.84 | 8.17 | 1.05 | 12.87 | 8.86 | 1.20 | 13.51 | [27,27,27-$^2H_3$]7αH,3O-CA |
| 3βH-$\Delta^5$-BA | 4.46 | 1.79 | 3.78 | 1.32 | 4.17 | 2.07 | 3.49 | 0.76 | 21.70 | 4.19 | 0.33 | 7.96 | [26,26,26,27,27,27-$^2H_6$]24R/S-HC |
| Desmosterol | 176.22 | 124.95 | 189.58 | 81.09 | 207.16 | 368.31 | 195.97 | 14.51 | 7.41 | 130.44 | 8.32 | 6.38 | [26,26,26,27,27,27-$^2H_6$]Desmosterol |

| | | | | | | | | | | | | | |
|---|---|---|---|---|---|---|---|---|---|---|---|---|---|
| 22-Dehydrocholesterol[e] | 172.21 | 58.51 | 202.06 | 78.12 | 194.45 | 75.76 | 116.54 | 13.02 | 11.17 | 96.12 | 6.13 | 6.38 | [26,26,26,27,27,27-$^{2}H_{6}$]Desmosterol |
| 7-Dehydrocholesterol | 108.30 | 28.44 | 114.58 | 43.77 | 137.06 | 58.96 | 68.51 | 9.15 | 13.36 | 73.10 | 8.85 | 12.11 | [26,26,26,27,27,27-$^{2}H_{6}$]Desmosterol |
| 8(14)-Dehydrocholesterol | 132.53 | 49.68 | 155.46 | 119.13 | 169.15 | 159.90 | 106.54 | 11.90 | 11.17 | 91.58 | 7.16 | 7.82 | [26,26,26,27,27,27-$^{2}H_{6}$]Desmosterol |
| 8-Dehydrocholesterol | 131.15 | 116.16 | 78.74 | 51.87 | 219.85 | 305.00 | 103.99 | 10.96 | 10.54 | 148.25 | 10.21 | 6.89 | [26,26,26,27,27,27-$^{2}H_{6}$]Desmosterol |
| Brassicasterol | 82.94 | 43.69 | 115.28 | 52.47 | 88.72 | 47.32 | 65.04 | 8.55 | 13.15 | 19.61 | 1.59 | 8.13 | [26,26,26,27,27,27-$^{2}H_{6}$]Desmosterol |
| | **Concentration (µg/mL)** | | | | | | | | | **Concentration (µg/mL)** | | | |
| Cholesterol | 520.88 | 134.76 | 530.75 | 155.76 | 529.02 | 264.52 | 485.62 | 21.28 | 4.38 | 405.86 | 16.13 | 3.97 | [25,26,26,27,27,27-$^{2}H_{7}$]Cholesterol |

[a][25,26,26,26,27,27,27-$^{2}H_{7}$]7α-HC oxidised to [25,26,26,26,27,27,27]7α-HCO internal standard and [25,26,26,26,27,27,27-$^{2}H_{7}$]22S-HCO used for any A/B volume correction.

[b]6β-HC is derived from the dehydration of cholestane-3β,5α,6β-triol which can be formed via the hydration of 3β-hydroxycholestan-5,6-epoxide.

[c]Authentic standard not available. Identification based on exact mass and $MS^3$ spectrum. Possible alternative structure 3β,20-dihydroxycholest-5-en-22-one.

[d][26,26,26,27,27,27-$^{2}H_{6}$]7α,25-diHC oxidised to [26,26,26,27,27,27-$^{2}H_{6}$]7α,25-diHCO internal standard and [27,27,27-$^{2}H_{3}$]7αH,3O-CA used for any A/B volume correction.

[e]Authentic standard not available. Identification based on exact mass and $MS^3$ spectrum.

[f]Authentic standard not available. Identification based on exact mass and $MS^3$ spectrum. Possible alternative structure 3β,27-dihydroxycholest-5-en-26-oic acid.

**Table S3B. Concentrations measured in plasma from people not taking statins, plasma concentrations are for the "free" sterols/oxysterols**

| | Healthy Controls (n = 75) | | Premanifest HD (n = 124) | | Manifest HD (n = 171) | | QC Replicates (n = 52) | | | NIST SRM 1950 (n = 6) | | | Isotope labelled standard |
|---|---|---|---|---|---|---|---|---|---|---|---|---|---|
| | Concentration (ng/mL) | | | | | | | | | Concentration (ng/mL) | | | |
| **Compound Name** | **MEAN** | **SD** | **MEAN** | **SD** | **MEAN** | **SD** | **MEAN** | **SD** | **%CV** | **MEAN** | **SD** | **%CV** | |
| 24S-HC | 19.88 | 4.49 | 19.62 | 4.94 | 17.51 | 4.66 | 21.38 | 0.90 | 4.21 | 8.94 | 0.47 | 5.26 | [26,26,26,27,27,27-$^2H_6$]24R/S-HC |
| 25-HC | 1.96 | 0.42 | 1.93 | 0.46 | 1.91 | 0.37 | 1.57 | 0.16 | 9.90 | 1.32 | 0.06 | 4.69 | [26,26,26,27,27,27-$^2H_6$]24R/S-HC |
| 26-HC | 40.38 | 10.42 | 40.93 | 9.41 | 40.11 | 8.96 | 39.10 | 2.30 | 5.88 | 22.30 | 0.68 | 3.07 | [26,26,26,27,27,27-$^2H_6$]24R/S-HC |
| 7α-HC | 6.92 | 6.72 | 6.55 | 7.08 | 7.72 | 9.04 | 6.22 | 1.46 | 23.53 | 13.23 | 1.50 | 11.34 | [25,26,26,26,27,27,27-$^2H_7$]7α-HC |
| 7α-HCO | 16.52 | 22.09 | 16.22 | 14.66 | 20.06 | 25.30 | 12.80 | 1.84 | 14.38 | 19.70 | 0.84 | 4.27 | [25,26,26,26,27,27,27-$^2H_7$]7α-HC[a] |
| 7-OC | 4.00 | 2.61 | 4.52 | 5.63 | 7.67 | 31.29 | 3.13 | 0.80 | 25.68 | 6.21 | 1.99 | 31.98 | [25,26,26,26,27,27,27-$^2H_7$-]7-OC |
| 6β-HC[b] | 2.16 | 1.77 | 2.41 | 2.83 | 2.55 | 2.72 | 1.77 | 1.21 | 68.39 | 4.62 | 2.06 | 44.53 | [25,26,26,26,27,27,27-$^2H_7$]7α-HC |
| 7β-HC | 1.85 | 0.85 | 2.07 | 3.00 | 3.09 | 7.70 | 1.35 | 0.43 | 31.64 | 4.24 | 0.62 | 14.50 | [25,26,26,26,27,27,27-$^2H_7$]7α-HC |
| 3β,22-diHC-24O[c] | 15.43 | 5.97 | 18.10 | 8.52 | 15.67 | 6.89 | 12.49 | 1.62 | 12.99 | 13.59 | 0.47 | 3.44 | [26,26,26,27,27,27-$^2H_6$]24R/S-HC |
| 3β-HCA | 125.05 | 32.55 | 135.18 | 41.33 | 126.37 | 35.25 | 114.24 | 4.98 | 4.36 | 101.91 | 4.34 | 4.25 | [24,24,27,27,27-$^2H_5$]3β-HCA |
| 7α,25-diHC | 1.20 | 0.65 | 1.22 | 0.62 | 1.20 | 0.49 | 0.87 | 0.37 | 42.93 | 0.94 | 0.21 | 22.47 | [26,26,26,27,27,27-$^2H_6$]7α,25-diHC |
| 7α,25-diHCO | 2.30 | 0.69 | 2.11 | 0.74 | 2.21 | 0.74 | 1.31 | 0.36 | 27.13 | 1.92 | 0.12 | 6.08 | [26,26,26,27,27,27-$^2H_6$]7α,25-diHC[d] |
| 7α,26-diHC | 1.78 | 1.30 | 1.62 | 1.02 | 1.78 | 1.29 | 1.26 | 0.68 | 53.62 | 0.64 | 0.33 | 52.22 | [26,26,26,27,27,27-$^2H_6$]7α,25-diHC |
| 7α,26-diHCO | 8.37 | 3.13 | 8.27 | 3.15 | 8.91 | 4.09 | 6.70 | 1.08 | 16.18 | 7.58 | 0.72 | 9.54 | [26,26,26,27,27,27-$^2H_6$]7α,25-diHC[d] |
| 3β,7α-diHCA | 55.51 | 21.32 | 55.50 | 21.45 | 57.32 | 23.69 | 48.93 | 4.27 | 8.74 | 46.89 | 6.02 | 12.83 | [27,27,27-$^2H_3$]7αH,3O-CA |
| 7αH,3O-CA | 126.14 | 35.20 | 125.84 | 34.41 | 129.52 | 40.92 | 130.35 | 6.48 | 4.98 | 104.24 | 3.01 | 2.89 | [27,27,27-$^2H_3$]7αH,3O-CA |
| 3β,7α-diHCA(25S) | 14.81 | 4.71 | 16.13 | 6.08 | 15.31 | 5.51 | 13.26 | 1.73 | 13.08 | 13.67 | 1.37 | 10.02 | [27,27,27-$^2H_3$]7αH,3O-CA |
| 7αH,3O-CA(25S) | 17.11 | 5.06 | 17.01 | 4.28 | 17.23 | 5.60 | 16.46 | 1.65 | 10.02 | 14.43 | 0.59 | 4.06 | [27,27,27-$^2H_3$]7αH,3O-CA |
| 3β,25-diHCA[e] | 10.17 | 5.15 | 10.38 | 5.71 | 9.23 | 4.21 | 6.49 | 1.66 | 25.58 | 8.29 | 1.06 | 12.76 | [26,26,26,27,27,27-$^2H_6$]24R/S-HC |
| 3β,22-diHCA[f] | 6.21 | 2.42 | 5.72 | 1.73 | 5.75 | 2.09 | 4.20 | 0.85 | 20.21 | 5.48 | 0.67 | 12.30 | [26,26,26,27,27,27-$^2H_6$]24R/S-HC |
| 3β,7β-diHCA | 12.86 | 5.12 | 14.31 | 8.91 | 16.65 | 30.20 | 10.64 | 1.58 | 14.86 | 10.63 | 0.62 | 5.85 | [27,27,27-$^2H_3$]7αH,3O-CA |
| 3β,7β-diH-$\Delta^5$-BA | 6.55 | 3.76 | 5.96 | 4.54 | 6.73 | 8.10 | 4.38 | 0.82 | 18.62 | 5.83 | 1.14 | 19.51 | [27,27,27-$^2H_3$]7αH,3O-CA |
| 3β,7α-diH-$\Delta^5$-BA | 7.51 | 4.98 | 6.77 | 4.10 | 7.25 | 4.63 | 5.82 | 1.59 | 27.28 | 6.53 | 2.01 | 30.86 | [27,27,27-$^2H_3$]7αH,3O-CA |
| 7αH,3O-$\Delta^4$-BA | 7.43 | 3.97 | 7.37 | 3.08 | 7.15 | 3.01 | 8.17 | 1.05 | 12.87 | 8.86 | 1.20 | 13.51 | [27,27,27-$^2H_3$]7αH,3O-CA |
| 3βH-$\Delta^5$-BA | 4.64 | 2.33 | 4.76 | 2.00 | 4.23 | 1.74 | 3.49 | 0.76 | 21.70 | 4.19 | 0.33 | 7.96 | [26,26,26,27,27,27-$^2H_6$]24R/S-HC |
| Desmosterol | 232.59 | 89.63 | 223.76 | 93.22 | 222.68 | 101.97 | 195.97 | 14.51 | 7.41 | 130.44 | 8.32 | 6.38 | [26,26,26,27,27,27-$^2H_6$]Desmosterol |

| | | | | | | | | | | | | |
|---|---|---|---|---|---|---|---|---|---|---|---|---|
| 22-Dehydrocholesterol[e] | 138.59 | 47.00 | 126.51 | 34.90 | 149.07 | 98.25 | 116.54 | 13.02 | 11.17 | 96.12 | 6.13 | 6.38 | [26,26,26,27,27,27-$^{2}H_{6}$]Desmosterol |
| 7-Dehydrocholesterol | 87.80 | 23.55 | 81.54 | 20.66 | 116.83 | 123.46 | 68.51 | 9.15 | 13.36 | 73.10 | 8.85 | 12.11 | [26,26,26,27,27,27-$^{2}H_{6}$]Desmosterol |
| 8(14)-Dehydrocholesterol | 115.36 | 44.08 | 107.87 | 41.12 | 148.90 | 157.79 | 106.54 | 11.90 | 11.17 | 91.58 | 7.16 | 7.82 | [26,26,26,27,27,27-$^{2}H_{6}$]Desmosterol |
| 8-Dehydrocholesterol | 112.21 | 59.66 | 105.82 | 58.18 | 268.82 | 641.41 | 103.99 | 10.96 | 10.54 | 148.25 | 10.21 | 6.89 | [26,26,26,27,27,27-$^{2}H_{6}$]Desmosterol |
| Brassicasterol | 86.18 | 52.13 | 87.87 | 47.44 | 80.44 | 50.24 | 65.04 | 8.55 | 13.15 | 19.61 | 1.59 | 8.13 | [26,26,26,27,27,27-$^{2}H_{6}$]Desmosterol |
| | **Concentration (µg/mL)** | | | | | | | | | **Concentration (µg/mL)** | | | |
| Cholesterol | 548.43 | 109.09 | 513.62 | 96.21 | 568.78 | 98.97 | 485.62 | 21.28 | 4.38 | 405.86 | 16.13 | 3.97 | [25,26,26,27,27,27-$^{2}H_{7}$]Cholesterol |

[a][25,26,26,26,27,27,27-$^{2}H_{7}$]7α-HC oxidised to [25,26,26,26,27,27,27-$^{2}H_{7}$] 7α-HCO internal standard and [25,26,26,26,27,27,27-$^{2}H_{7}$]22S-HCO used for any A/B volume correction.

[b]6β-HC is derived from the dehydration of cholestane-3β,5α,6β-triol which can be formed via the hydration of 3β-hydroxycholestan-5,6-epoxide.

[c]Authentic standard not available. Identification based on exact mass and $MS^3$ spectrum. Possible alternative structure 3β,20-dihydroxycholest-5-en-22-one.

[d][26,26,26,27,27,27-$^{2}H_{6}$]7α,25-diHC oxidised to [26,26,26,27,27,27-$^{2}H_{6}$]7α,25-diHCO internal standard and [27,27,27-$^{2}H_{3}$]7αH-3O-CA used for any A/B volume correction.

[e]Authentic standard not available. Identification based on exact mass and $MS^3$ spectrum.

[f]Authentic standard not available. Identification based on exact mass and $MS^3$ spectrum. Possible alternative structure 3β,27-dihydroxycholest-5-en-26-oic acid.

**Table S3C. Concentrations measured in CSF samples from people taking statins, CSF concentrations are for the "total" sterols/oxysterols following base hydrolysis**

| | Healthy Controls (n = 8) | | Premanifest HD (n = 5) | | Manifest HD (n = 17) | | QC Replicates (n = 51) | | | Isotope labelled standard |
|---|---|---|---|---|---|---|---|---|---|---|
| | Concentration (ng/mL) | | | | | | | | | |
| **Compound Name** | **MEAN** | **SD** | **MEAN** | **SD** | **MEAN** | **SD** | **MEAN** | **SD** | **%CV** | |
| 24S-HC | 2.01 | 0.62 | 1.68 | 0.48 | 2.42 | 1.16 | 1.85 | 0.12 | 6.40 | [26,26,26,27,27,27-$^2H_6$]24R/S-HC |
| 25-HC | 0.18 | 0.07 | 0.14 | 0.03 | 0.22 | 0.09 | 0.16 | 0.02 | 13.11 | [26,26,26,27,27,27-$^2H_6$]24R/S-HC |
| 26-HC | 1.43 | 0.52 | 1.35 | 0.28 | 1.65 | 0.49 | 1.50 | 0.12 | 7.89 | [26,26,26,27,27,27-$^2H_6$]24R/S-HC |
| 7α-HC | 0.27 | 0.17 | 0.20 | 0.05 | 0.30 | 0.19 | 0.28 | 0.16 | 58.34 | [25,26,26,26,27,27,27-$^2H_7$]7α-HC |
| 6β-HC[a] | 1.43 | 1.18 | 0.98 | 0.49 | 1.48 | 0.69 | 1.41 | 0.78 | 55.72 | [25,26,26,26,27,27,27-$^2H_7$]7α-HC |
| 7β-HC | 0.21 | 0.16 | 0.14 | 0.05 | 0.28 | 0.25 | 0.24 | 0.16 | 65.03 | [25,26,26,26,27,27,27-$^2H_7$]7α-HC |
| 7α,25-diHC | 0.15 | 0.05 | 0.15 | 0.05 | 0.19 | 0.05 | 0.15 | 0.03 | 19.74 | [26,26,26,27,27,27-$^2H_6$]7α,25-diHC |
| 7α,26-diHC | 0.21 | 0.10 | 0.22 | 0.06 | 0.24 | 0.08 | 0.21 | 0.03 | 15.85 | [26,26,26,27,27,27-$^2H_6$]7α,25-diHC |
| 3β-HCA | 1.15 | 0.39 | 1.48 | 0.19 | 1.27 | 0.33 | 1.46 | 0.18 | 12.15 | [24,24,27,27,27-$^2H_5$]3β-HCA |
| 7αH,3O-CA[b] | 29.34 | 7.98 | 31.92 | 3.93 | 31.71 | 9.07 | 32.99 | 1.81 | 5.49 | [27,27,27-$^2H_3$]7αH,3O-CA |
| 7-OC | 0.79 | 0.37 | 1.06 | 0.52 | 1.02 | 0.54 | 1.10 | 0.43 | 39.00 | [25,26,26,26,27,27,27-$^2H_7$]7-OC |
| Desmosterol | 5.36 | 1.10 | 6.25 | 0.85 | 6.43 | 4.90 | 6.19 | 0.49 | 7.93 | [26,26,26,27,27,27-$^2H_6$]Desmosterol |
| 22-Dehydrocholesterol[c] | 2.49 | 0.82 | 2.76 | 0.84 | 2.88 | 0.98 | 2.44 | 0.34 | 14.11 | [26,26,26,27,27,27-$^2H_6$]Desmosterol |
| 7-Dehydrocholesterol | 0.82 | 0.25 | 0.93 | 0.24 | 1.20 | 0.39 | 0.87 | 0.13 | 14.42 | [26,26,26,27,27,27-$^2H_6$]Desmosterol |
| 8(14)-Dehydrocholesterol | 1.95 | 0.45 | 2.30 | 0.16 | 2.79 | 0.94 | 2.19 | 0.20 | 9.25 | [26,26,26,27,27,27-$^2H_6$]Desmosterol |
| 8-Dehydrocholesterol | 2.42 | 0.94 | 2.14 | 0.59 | 4.52 | 3.84 | 2.83 | 0.24 | 8.33 | [26,26,26,27,27,27-$^2H_6$]Desmosterol |
| 6-Dehydrocholesterol[d] | 1.26 | 0.39 | 1.36 | 0.60 | 1.68 | 0.60 | 1.39 | 0.42 | 30.31 | [26,26,26,27,27,27-$^2H_6$]Desmosterol |
| Brassicasterol | 0.92 | 0.42 | 0.94 | 0.61 | 0.96 | 0.39 | 0.90 | 0.32 | 35.94 | [26,26,26,27,27,27-$^2H_6$]Desmosterol |
| Lanosterol | 0.71 | 0.73 | 0.94 | 0.63 | 3.99 | 11.25 | 0.87 | 0.42 | 48.66 | [26,26,26,27,27,27-$^2H_6$]Lanosterol |
| | Concentration (μg/mL) | | | | | | | | | |
| Cholesterol (μg/mL) | 3.76 | 0.79 | 4.31 | 0.93 | 4.50 | 1.05 | 4.45 | 0.26 | 5.74 | [25,26,26,27,27,27-$^2H_7$]Cholesterol |

[a]6β-HC is derived from the dehydration of cholestane-3β,5α,6β-triol which can be formed via the hydration of 3β-hydroxycholestan-5,6-epoxide.
[b]Measured as the dehydrated molecule.
[c]Authentic standard not available. Identification based on exact mass and $MS^3$ spectrum.
[d]Likely autoxidation product.

**Table S3D. Concentrations measured in CSF samples from people not taking statins, CSF concentrations are for the "total" sterols/oxysterols following base hydrolysis**

| | Healthy Controls (n = 75) | | Premanifest HD (n = 124) | | Manifest HD (n = 171) | | QC Replicates (n = 51) | | | Isotope labelled standard |
|---|---|---|---|---|---|---|---|---|---|---|
| | Concentration (ng/mL) | | | | | | | | | |
| Compound Name | MEAN | SD | MEAN | SD | MEAN | SD | MEAN | SD | %CV | |
| 24S-HC | 1.79 | 0.69 | 1.71 | 0.61 | 2.01 | 0.88 | 1.85 | 0.12 | 6.40 | [26,26,26,27,27,27-$^2H_6$]24R/S-HC |
| 25-HC | 0.17 | 0.08 | 0.15 | 0.06 | 0.17 | 0.06 | 0.16 | 0.02 | 13.11 | [26,26,26,27,27,27-$^2H_6$]24R/S-HC |
| 26-HC | 1.40 | 0.59 | 1.32 | 0.52 | 1.46 | 0.58 | 1.50 | 0.12 | 7.89 | [26,26,26,27,27,27-$^2H_6$]24R/S-HC |
| 7α-HC | 0.32 | 0.19 | 0.30 | 0.20 | 0.28 | 0.15 | 0.28 | 0.16 | 58.34 | [25,26,26,26,27,27,27-$^2H_7$]7α-HC |
| 6β-HC[a] | 1.57 | 0.80 | 1.49 | 1.03 | 1.50 | 0.80 | 1.41 | 0.78 | 55.72 | [25,26,26,26,27,27,27-$^2H_7$]7α-HC |
| 7β-HC | 0.30 | 0.23 | 0.27 | 0.18 | 0.25 | 0.17 | 0.24 | 0.16 | 65.03 | [25,26,26,26,27,27,27-$^2H_7$]7α-HC |
| 7α,25-diHC | 0.16 | 0.05 | 0.15 | 0.05 | 0.16 | 0.05 | 0.15 | 0.03 | 19.74 | [26,26,26,27,27,27-$^2H_6$]7α,25-diHC |
| 7α,26-diHC | 0.21 | 0.07 | 0.19 | 0.07 | 0.22 | 0.08 | 0.21 | 0.03 | 15.85 | [26,26,26,27,27,27-$^2H_6$]7α,25-diHC |
| 3β-HCA | 1.13 | 0.50 | 1.21 | 0.52 | 1.22 | 0.59 | 1.46 | 0.18 | 12.15 | [24,24,27,27,27-$^2H_5$]3β-HCA |
| 7αH,3O-CA[b] | 27.56 | 9.05 | 28.85 | 10.50 | 29.31 | 9.49 | 32.99 | 1.81 | 5.49 | [27,27,27-$^2H_3$]7αH,3O-CA |
| 7-OC | 1.25 | 1.21 | 1.17 | 0.63 | 1.16 | 0.76 | 1.10 | 0.43 | 39.00 | [25,26,26,26,27,27,27-$^2H_7$]7-OC |
| Desmosterol | 6.11 | 1.52 | 5.86 | 1.56 | 6.07 | 2.20 | 6.19 | 0.49 | 7.93 | [26,26,26,27,27,27-$^2H_6$]Desmosterol |
| 22-Dehydrocholesterol[c] | 2.27 | 0.59 | 2.10 | 0.60 | 2.28 | 0.62 | 2.44 | 0.34 | 14.11 | [26,26,26,27,27,27-$^2H_6$]Desmosterol |
| 7-Dehydrocholesterol | 0.78 | 0.20 | 0.76 | 0.21 | 0.90 | 0.39 | 0.87 | 0.13 | 14.42 | [26,26,26,27,27,27-$^2H_6$]Desmosterol |
| 8(14)-Dehydrocholesterol | 1.84 | 0.58 | 1.76 | 0.52 | 2.02 | 0.82 | 2.19 | 0.20 | 9.25 | [26,26,26,27,27,27-$^2H_6$]Desmosterol |
| 8-Dehydrocholesterol | 2.21 | 1.08 | 2.18 | 0.82 | 3.42 | 3.38 | 2.83 | 0.24 | 8.33 | [26,26,26,27,27,27-$^2H_6$]Desmosterol |
| 6-Dehydrocholesterol[d] | 1.37 | 0.50 | 1.35 | 0.45 | 1.48 | 0.57 | 1.39 | 0.42 | 30.31 | [26,26,26,27,27,27-$^2H_6$]Desmosterol |
| Brassicasterol | 0.85 | 0.37 | 0.83 | 0.35 | 0.84 | 0.35 | 0.90 | 0.32 | 35.94 | [26,26,26,27,27,27-$^2H_6$]Desmosterol |
| Lanosterol | 1.08 | 1.16 | 1.32 | 2.37 | 1.49 | 2.89 | 0.87 | 0.42 | 48.66 | [26,26,26,27,27,27-$^2H_6$]Lanosterol |
| | Concentration (μg/mL) | | | | | | | | | |
| Cholesterol (μg/mL) | 3.82 | 1.07 | 3.67 | 1.04 | 3.96 | 1.03 | 4.45 | 0.26 | 5.74 | [25,26,26,27,27,27-$^2H_7$]Cholesterol |

[a]6β-HC is derived from the dehydration of cholestane-3β,5α,6β-triol which can be formed via the hydration of 3β-hydroxycholestan-5,6-epoxide.
[b]Measured as the dehydrated molecule.
[c]Authentic standard not available. Identification based on exact mass and $MS^3$ spectrum.
[d]Likely autoxidation product.
