## Supplemental Table S4 for "24S-Hydroxycholesterol: A potential brain-derived biomarker of Huntington’s Disease"

**Supplementary Table S4 Consensus ranking lists of seven binary classification techniques for each of three pairwise comparisons amongst control, premanifest HD and manifest HD**

| | | | | |
|---|---|---|---|---|
| **Manifest HD V Premanifest HD** | **Manifest HD V Control** | **Premanifest HD V Control** | **Overall Consensus** | ***P*-value** |
| Age | BMI | Age | Age | 0.000240 |
| (plasma) 24S-HC | (plasma) 24SHC | BMI | BMI | 0.002152 |
| (plasma) 7-DHC | (CSF) 8-DHC | (CSF) 25-HC | (plasma) 24S-HC | 0.002645 |
| (plasma) Cholesterol | (plasma) 8(14)-DHC | (CSF) 3βHCA | (plasma) 7-DHC | 0.007519 |
| (CSF) 24S-HC | (plasma) 7-DHC | (CSF) 7αH,3O-CA | (plasma) Cholesterol | 0.009328 |
| (CSF) 3β-HCA | (plasma) 3β,22-diHCA | (plasma) 3β,7β-diH$\Delta^5$-BA | (CSF) 8-DHC | 0.015363 |
| (CSF) 8-DHC | Sex | (plasma) 3β,7β-diHCA | (CSF) 3β-HCA | 0.018560 |
| (plasma) 3β-HCA | (CSF) 24S-HC | (plasma) 3βHCA | (CSF) 24S-HC | 0.019775 |
| (CSF) 7αH,3O-CA | (CSF) 25-HC | (plasma) 3β,22-diHCA | (plasma) 8(14)-DHC | 0.031471 |
| (CSF) 7-DHC | (plasma) Cholesterol | (CSF) 22-DHC | (CSF) 7β-HC | 0.034302 |
| (CSF) 7α,26-diHC | (CSF) 7-DHC | (CSF) 7α,25-diHC | (CSF) 7αH,3O-CA | 0.034470 |
| (CSF) 6-DHC | (CSF) 6-DHC | (plasma) 3β,7α-diH-$\Delta^5$-BA | (CSF) 25-HC | 0.035540 |
| (plasma) 22-DHC | (CSF) 7β-HC | (plasma) 3β,7α-diHCA(25S) | (plasma) 3β,7β-diHCA | 0.038287 |
| (plasma) 3β,7β-diHCA | Age | (plasma) Cholesterol | (plasma) 3β,22-diHCA | 0.039273 |
| (plasma) 3β,7α-diHCA | (CSF) Brassicasterol | (plasma) 3β,7α-diHCA | (plasma) 3β-HCA | 0.044622 |
| (plasma) 8-DHC | (plasma) 6β-HC | Sex | Sex | 0.054948 |

Variables are listed in order of importance and colour coded according to their position in an overall consensus of the three lists obtained using the Stuart rank aggregation method. Only the most important 16 variables are shown. A significant *P*-value occurs for variables consistently higher in all three lists. Variables not colour coded fall below sixteenth place in the consensus.
Note. In CSF 7αH,3O-CA is measured as the dehydrated molecule.
If not listed in the main text abbreviations can be found in Supplementary Table S2.
