## Supplemental Text 1 for "24S-Hydroxycholesterol: A potential brain-derived biomarker of Huntington’s Disease"

### Supplementary Material 1

### Methods

#### LC-MS Gradients

##### Oxysterol 17 min gradient

Samples were injected in 60% methanol. Chromatographic separation was achieved with a reversed-phase Hypersil Gold $C_{18}$ column (1.9 µm particle size, 50 x 2.1 mm, Thermo Fisher) using an Ultimate 3000 LC system (Thermo Fisher Scientific). The mobile phase composition was initially at 80% A (33.3% methanol, 16.7% acetonitrile, 0.1% formic acid) and 20% B (63.3% methanol, 31.7% acetonitrile, 0.1% formic acid) and held at this composition for 1 min then changed over 7 min to 20% A, 80% B and maintained at this composition for 4 min. The mobile phase was returned to 80% A, 20% B in 6 s and the column reconditioned for a further 4 min 54 s. The flow-rate was 200 µL/min and the eluate introduced to the Orbitrap Elite mass spectrometer (Thermo Fisher Scientific) via an electrospray ionisation (ESI) source.

##### Oxysterol 37 min gradient

Samples were injected in 60% methanol. Chromatographic separation was achieved with a reversed-phase Hypersil Gold $C_{18}$ column (1.9 µm particle size, 50 x 2.1 mm, Thermo Fisher) using an Ultimate 3000 LC system (Thermo Fisher Scientific). The mobile phase composition was initially at 80% A (33.3% methanol, 16.7% acetonitrile, 0.1% formic acid) and 20% B (63.3% methanol, 31.7% acetonitrile, 0.1% formic acid) and held at this composition for 10 min then changed over 10 min to 50% A, 50% B and maintained at this composition for 6 min. The proportion of B increased to 80% over 3 minutes and maintained for a further 3 min. The mobile phase was returned to 80% A, 20% B in 6 s and the column reconditioned for a further 4 min 54 s. The flow-rate was 200 µL/min and the eluate introduced to the Orbitrap Elite mass spectrometer (Thermo Fisher Scientific) via an ESI source.

##### Sterols 24 min gradient

Samples were injected in 75% methanol. Chromatographic separation was achieved with a reversed-phase ACE $C_{18}$ column (10 cm x 2.1 mm, 2 µm, Advanced Chromatography Technologies Ltd) using an Ultimate 3000 LC system (Thermo Fisher Scientific). The mobile phase composition was initially at 22% A (33.3% methanol, 16.7% acetonitrile, 0.1% formic acid) and 78% B (63.3% methanol, 31.7% acetonitrile, 0.1% formic acid) and changed over 8 min to 20% A, 80% B then changed to 15% A, 85 %B over 3 min and maintained at this composition for a further 6 min. The mobile phase was returned to 22% A, 78% B in 6 s and the column reconditioned for a further 6 min 54 s. The flow-rate was 200 µL/min and the eluate introduced to the Orbitrap Elite mass spectrometer (Thermo Fisher Scientific) via an ESI source.

##### Lanosterol 17 min gradient

Samples were injected in 80% ethanol 0.1% formic acid. Chromatographic separation was achieved with a reversed-phase Hypersil Gold $C_{18}$ column (1.9 µm particle size, 20 x 2.1 mm, Thermo Fisher) using an Ultimate 3000 LC system (Thermo Fisher Scientific). The mobile phase composition was initially at 22% A (50% methanol, 0.1% formic acid) and 78% B (90% methanol, 5% propan-2-ol, 0.1% formic acid) and changed to 100%B in 1 min, kept at this composition for 12 min, then returned to 22% A, 78% B in 6 s and the column reconditioned for a further 3 min 54 s. The flow-rate was 200 µL/min and the eluate introduced to the Orbitrap IQX mass spectrometer (Thermo Fisher Scientific) via an ESI source.

### Scan Parameters

#### Oxysterol Analysis - Plasma

1. 17 min. (1) $MS^1$ Orbitrap 400 – 610, Resolution 120,000; (2) $MS^3$ LIT 506.34(30%)→427.30(35%)→; (3) $MS^3$ LIT 511.37(30%)→427.30(35%)→; (4) $MS^3$ LIT 546.37(30%)→467.33(35%)→; (5) $MS^3$ LIT 551.40(30%)→467.33(35%)→.
2. 17 min. (1) $MS^1$ Orbitrap 400 – 610, Resolution 120,000; (2) $MS^3$ LIT 534.41(30%)→455.36(35%)→; (3) $MS^3$ LIT 539.44(30%)→455.36(35%)→; (4) $MS^3$ LIT 541.45(30%)→462.41(35%)→; (5) $MS^3$ LIT 545.47(30%)→461.40(35%); (6) $MS^3$ LIT 546.48(30%)→462.42(35%)→.
3. 37 min. (1) $MS^1$ Orbitrap 400 – 610, Resolution 240,000; (2) $MS^3$ LIT 534.41(30%)→455.36(35%)→; (3) $MS^3$ LIT 539.44(30%)→455.36(35%)→; (4) $MS^3$ LIT 541.45(30%)→462.41(35%)→; (5) $MS^3$ LIT 545.47(30%)→461.40(35%); (6) $MS^3$ LIT 546.48(30%)→462.42(35%)→.
4. 17 min. (1) $MS^1$ Orbitrap 400 – 610, Resolution 120,000; (2) $MS^3$ LIT 550.40(30%)→471.36(35%)→; (3) $MS^3$ LIT 555.43(30%)→471.36(35%)→; (4) $MS^3$ LIT 561.47(30%)→477.40(35%)→.
5. 17 min. (1) $MS^1$ Orbitrap 400 – 610, Resolution 120,000; (2) $MS^3$ LIT 564.38(30%)→485.34(35%)→; (3) $MS^3$ LIT 567.40(30%)→488.36(35%)→; (4) $MS^3$ LIT 569.41(30%)→485.34(35%)→; (5) $MS^3$ LIT 572.43(30%)→488.36(35%)→.
6. 17 min. (1) $MS^1$ Orbitrap 400 – 610, Resolution 120,000; (2) $MS^3$ LIT 548.38(30%)→469.34(35%)→; (3) $MS^3$ LIT 553.42(30%)→469.34(35%); (4) $MS^3$ LIT 553.42(30%)→474.4(35%)→; (5) $MS^3$ LIT 558.45(30%)→474.4(35%)→.
7. 17 min. (1) $MS^1$ Orbitrap 300 – 800, Resolution 120,000; (2) $MS^2$ LIT 614.36(30%)→; (3) $MS^2$ LIT 619.39(30%)→; (4) $MS^2$ LIT 710.44(30%)→; (5) $MS^2$ LIT 715.47(30%)→.
8. 17 min. (1) $MS^1$ Orbitrap 400 – 610, Resolution 120,000; (2) $MS^3$ LIT 522.33(30%)→443.29(35%)→; (3) $MS^3$ LIT 527.36(30%)→443.29(35%)→; (4) $MS^3$ LIT 532.39(30%)→453.35(35%)→; (5) $MS^3$ LIT 537.42(30%)→453.35(35%)→.
9. 17 min. (1) $MS^1$ Orbitrap 400 – 610, Resolution 120,000; (2) $MS^3$ LIT 562.36(30%)→483.32(35%)→; (3) $MS^3$ LIT 567.40(30%)→483.32(35%)→; (4) $MS^3$ LIT 578.36(30%)→499.32(35%)→; (5) $MS^3$ LIT 583.39(30%)→499.32(35%)→.

10. 17 min. (1) MS[1] Orbitrap 400 – 610, Resolution 120,000; (2) MS[3] LIT 566.40(30%)→487.35(35%)→; (3) MS[3] LIT 571.43(30%)→487.35(35%)→; (4) MS[3] LIT 580.37(30%)→501.33(35%)→; (5) MS[3] LIT 585.41(30%)→501.33(35%)→.

Orbitrap Elite. Scan range given in amu. Resolution selected at *m/z* 400. MS[3] transitions are described by the precursor-ion and product-ion selected for fragmentation with CID collision energy given as a % in parenthesis. Isolation width was 1 amu. Identity of precursor ion *m/z* can be found in Supplementary Table S2. If not listed in Table S2 the precursor is either an $[M-H_2O]^+$ ion or the $[^2H_0]$GP derivative of a 3-oxo-4-ene sterol not generally observed.

#### Oxysterol Analysis – CSF

1. 17 min. (1) MS[1] Orbitrap 400 – 610, Resolution 120,000; (2) MS[3] LIT 546.37(30%)→467.33(35%)→; (2) MS[3] LIT 549.39(30%)→470.35(35%)→; (3) 551.40(30%)→467.33(35%)→; (4) 554.42(30%)→470.35(35%)→.
2. 37 min. (1) MS[1] Orbitrap 400 – 610, Resolution 240,000; (2) MS[3] LIT 534.41(30%)→455.35(35%)→; (3) MS[3] LIT 539.44(30%)→455.36(35%)→; (4) MS[3] LIT 541.45(30%)→462.41(35%)→; (5) MS[3] LIT 545.47(30%)→461.40(35%)→; (6) MS[3] LIT 546.48 (30%)→462.41(35%)→.
3. 17 min. (1) MS[1] Orbitrap 400 – 610, Resolution 120,000; (2) MS[3] LIT 534.41(30%)→455.35(35%)→; (3) MS[3] LIT 539.44(30%)→455.36(35%)→; (4) MS[3] LIT 541.45(30%)→462.41(35%)→; (5) MS[3] LIT 545.47(30%)→461.40(35%)→; (6) MS[3] LIT 546.48 (30%)→462.41(35%)→.
4. 17 min. (1) MS[1] Orbitrap 400 – 610, Resolution 120,000; (2) MS[3] LIT 550.40(30%)→471.36(35%); (3) MS[3] LIT 555.43(30%)→471.36(35%)→; (4) MS[3] LIT 561.47(30%)→477.40(35%)→.
5. 17 min. (1) MS[1] Orbitrap 400 – 610, Resolution 120,000; (2) MS[3] LIT 548.38(30%)→469.34(35%)→; MS[3] LIT 553.42(30%)→469.34(35%)→; MS[3] LIT 553.42(30%)→474.4(35%)→; MS[3] LIT 558.45(30%)→474.37(35%)→.
6. 17 min. (1) MS[1] Orbitrap 400 – 610, Resolution 120,000; (2) MS[3] LIT 564.38(30%)→485.34(35%); (3) MS[3] LIT 567.40(30%)→488.36(35%); (4) MS[3] LIT 569.41(30%)→485.34(35%); (5) 572.43(30%)→488.36(35%)→.

Orbitrap Elite. Scan range given in amu. Resolution selected at *m/z* 400. MS[3] transitions are described by the precursor-ion and product-ion selected for fragmentation with CID collision energy given as a % in parenthesis. Isolation width was 1 amu. Identity of precursor ion *m/z* can be found in Supplementary Table S2. If not listed in Table S2 the precursor is either an $[M-H_2O]^+$ ion or the $[^2H_0]$GP derivative of a 3-oxo-4-ene sterol not generally observed.

#### Sterol Analysis - Plasma

1. 24 min. (1) MS[1] Orbitrap 400 – 610, Resolution 120,000; (2) MS[3] LIT 516.39(30%)→437.35(35%)→; (3) MS[3] LIT 521.43(30%)→437.35(35%)→; (4) MS[3] LIT 522.43(30%)→443.39(35%)→; (5) MS[3] LIT 527.46(30%)→443.39(35%); (6) MS[3] LIT 530.41(30%)→451.37(35%)→; (7) MS[3] LIT 535.44(30%)→451.37(35%)→.

2. 24 min. (1) MS$^1$ Orbitrap 400 – 610, Resolution 120,000; (2) MS$^3$ LIT 514.38(30%)→435.34(35%)→; (3) MS$^3$ LIT 518.41(30%)→439.37(35%)→; (4) MS$^3$ LIT 519.41(30%)→435.34(35%)→; (5) MS$^3$ LIT 523.44(30%)→439.37(35%)→; (6) 525.45(30%)→446.41(35%); (7) 530.49(30%)→446.41(35%)→.

Orbitrap Elite. Scan range given in amu. Resolution selected at *m/z* 400. MS$^3$ transitions are described by the precursor-ion and product-ion selected for fragmentation with CID collision energy given as a % in parenthesis. Isolation width was 1 amu. If not listed in Table S2 the precursor is the [$^2H_0$]GP derivative of a 3-oxo-4-ene sterol not generally observed.

#### Sterol Analysis - CSF

1. 24 min. (1) MS$^1$ Orbitrap 400 – 610, Resolution 120,000; (2) MS$^3$ LIT 516.39(30%)→437.35(35%)→; (3) MS$^3$ LIT 521.43(30%)→437.35(35%)→; (4) MS$^3$ LIT 522.43(30%)→443.39(35%)→; (5) MS$^3$ LIT 527.46(30%)→443.39(35%); (6) MS$^3$ LIT 530.41(30%)→451.37(35%)→; (7) MS$^3$ LIT 535.44(30%)→451.37(35%)→.
2. 24 min. (1) MS$^1$ Orbitrap 400 – 610, Resolution 120,000; (2) MS$^3$ LIT 514.38(30%)→435.34(35%)→; (3) MS$^3$ LIT 518.41(30%)→439.37(35%)→; (4) MS$^3$ LIT 519.41(30%)→435.34(35%)→; (5) MS$^3$ LIT 523.44(30%)→439.37(35%)→; (6) 525.45(30%)→446.41(35%); (7) 530.49(30%)→446.41(35%)→.

Orbitrap Elite. Scan range given in amu. Resolution selected at *m/z* 400. MS$^3$ transitions are described by the precursor-ion and product-ion selected for fragmentation with CID collision energy given as a % in parenthesis. Isolation width was 1 amu. If not listed in Table S2 the precursor is the [$^2H_0$]GP derivative of a 3-oxo-4-ene sterol not generally observed.

##### Lanosterol Analysis – CSF

1. 17 min. (1) MS$^1$ Orbitrap 350 – 450, Resolution 120,000; (2) MS$^2$ LIT 409.38(45%)→; (3) MS$^2$ LIT 415.42(45%)→; (4) MS$^2$ LIT 369.35(45%)→; MS$^2$ LIT 376.40(45%)→; (5) MS$^2$ LIT 367.34(45%)→; (6) MS$^2$ LIT 373.37(45%)→.

Orbitrap IQX. Scan range given in amu. Resolution selected at *m/z* 400. MS$^2$ scans are described by the precursor-ion selected for fragmentation with HCD and collision energy given as a % in parenthesis. Isolation window was 1.6 amu.

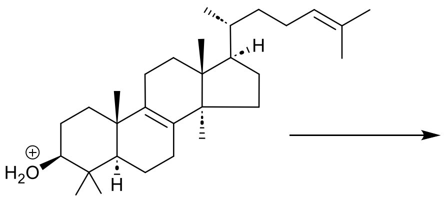


Chemical Formula: $C_{30}H_{51}O^+$
Exact Mass: 427.3934
Lanosterol $[M+H]^+$

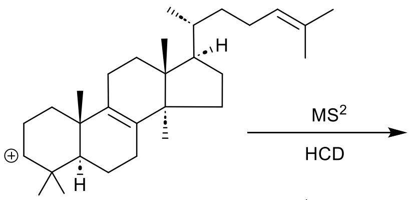


Chemical Formula: $C_{30}H_{49}^+$
Exact Mass: 409.3829
Lanosterol $[M-H_2O+H]^+$

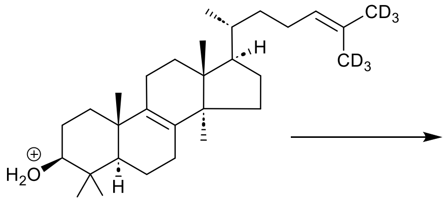


Chemical Formula: $C_{30}H_{45}D_6O^+$
Exact Mass: 433.4311
$[^2H_6]$Lanosterol $[M+H]^+$

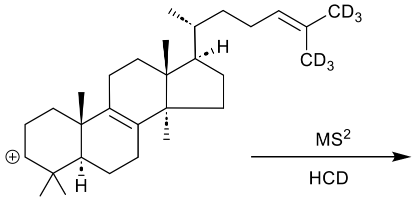


Chemical Formula: $C_{30}H_{43}D_6^+$
Exact Mass: 415.4205
$[^2H_6]$Lanosterol $[M-H_2O+H]^+$

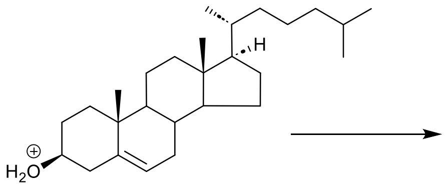


Chemical Formula: $C_{27}H_{47}O^+$
Exact Mass: 387.3621
Cholesterol $[M+H]^+$

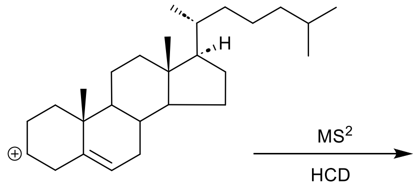


Chemical Formula: $C_{27}H_{45}^+$
Exact Mass: 369.3516
Cholesterol $[M-H_2O+H]^+$

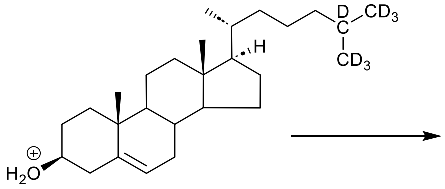


Chemical Formula: $C_{27}H_{40}D_7O^+$
Exact Mass: 394.4061
$[^2H_7]$Cholesterol $[M+H]^+$

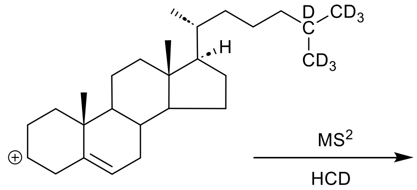


Chemical Formula: $C_{27}H_{38}D_7^+$
Exact Mass: 376.3955
$[^2H_7]$Cholesterol $[M-H_2O+H]^+$

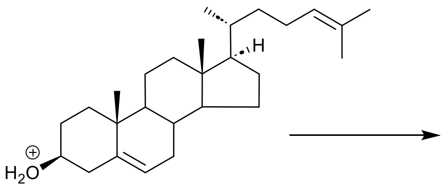


Chemical Formula: $C_{27}H_{45}O^+$
Exact Mass: 385.3465
Desmosterol $[M+H]^+$

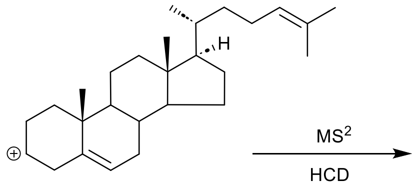


Chemical Formula: $C_{27}H_{43}^+$
Exact Mass: 367.3359
Desmoesterol $[M-H_2O+H]^+$

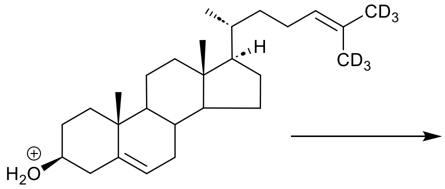


Chemical Formula: $C_{27}H_{39}D_6O^+$
Exact Mass: 391.3842
$[^2H_6]$Desmosterol $[M+H]^+$

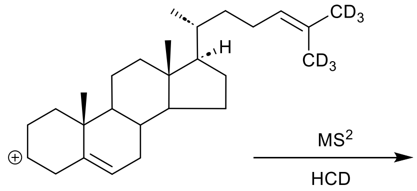


Chemical Formula: $C_{27}H_{37}D_6^+$
Exact Mass: 373.3736
$[^2H_6]$Desmoesterol $[M-H_2O+H]^+$
