## Supplemental Text 2 for "24S-Hydroxycholesterol: A potential brain-derived biomarker of Huntington’s Disease"

### Supplementary Material 2

#### Pairwise Comparison of three groups (Control, PreManifest and Manifest) using Binary Classification Techniques

#### Summary

Binary classification results for the comparative analysis of seven classification methods each applied separately to the three binary comparisons are given in Supplementary_Workbook_1 with a summary in Supplementary_Workbook_2 (both Workbooks available to view on figshare https://doi.org/10.6084/m9.figshare.30510515). A list of independent variables retained in the models is given together with the receiver operating characteristic (ROC) curve and area under the curve (AUC), histogram of probability of group membership, 2x2 confusion table, permutation analysis results, and associated statistics. Note that a list of abbreviations used in this section for metabolites can be found in Supplementary Table S2 where the lowercase letters p and c are used to indicate plasma and CSF, respectively. For the final five-fold cross-validation, summary statistics together with ROC curves and chi-square values for the confusion tables for each fold are shown. ROC curves are widely used in binary classification and show clinical sensitivity and specificity for a test where the AUC is a measure of the discriminative ability of a test. Tests with good diagnostic performance have a ROC curve that is close to the top-left corner, with an area under the curve that is close to 1, cf. Supplementary Figure S5 showing ROC curves for the PreManifest and Manifest comparison.

The AUC values for this comparison for the full dataset are consistently high (0.802 – 0.933). Similar AUC values were obtained for the five test samples from cross-validation (0.802 – 0.848). For the two binary comparisons involving Control, the full dataset also gives high values of AUC and significant values for the confusion table. However, for the cross-validation test samples the AUC values are lower and many of the p-values for the confusion tables are not significant. Overfitting in the larger full data set might overestimate significance whereas the smaller sample sizes of the cross-validation test samples might underestimate significance.

The values of AUC for forwards and backwards regression are similar to those for the regularisation techniques (lasso, ridge and elastic net). For the machine learning methods, gradient boosting has the highest AUC values for the full dataset of all methods but does not perform better in cross-validation. Random Forest has the lowest AUC values for the full dataset similar in magnitude to the averages of folds. The estimated p-values for AUC for the permutation analysis (worksheet AUC_2x2 section (A) in Supplementary_Workbook_2) are uniformly highly significant except for gradient boosting and random forest for the two comparisons with control. These differences might be the result of the machine learning methods being less prone to overfitting.

Ranked lists of independent variables retained by the seven different classification techniques are given for the full data set in Supplementary_Workbook_2 for the three binary group comparisons (worksheets PreMan, ConPre, and ConMan). These lists provide information on which of the independent variables are most important both for the binary classification and as variables (features) that are of importance in terms of disease mechanism.

For all three pairwise comparisons, the different classification methods demonstrate overlap in the variables of greatest importance. At the same time the methods also show some distinct differences from each other. The machine learning techniques (random forest and gradient boosting) possibly have slightly different rankings from the regression methods. At the same time the basic regression methods (forward and backward) do not produce results which deviate markedly from the other methods. The results underline the utility of testing different methods but also that, at least for the current data, grave errors might be unlikely when relying on a single technique for classification or feature selection.

For each of the three pairwise comparisons a single consensus ranking list for the seven different classification techniques was made using the Stuart aggregation method (Supplementary_Workbook_2, worksheet ClassListsCompared (A)). There is overlap in the variables at the top of the three lists. However, the important variables in the comparison of Control and PreManifest show differences from the other two comparisons involving Manifest which resemble each other more closely. This suggests that there might be qualitative differences in the most influential variables between the three comparisons.

A consensus was made of the ranking lists for the three pairwise comparisons (Supplementary Table S4 and Supplementary_Workbook_2, ClassListsCompared (B)). Consideration of these lists allowed the identification of the most important candidate variables overall for classification and feature selection. The highest ranked metabolites are plasma 24S-HC followed by plasma 7-DHC, plasma cholesterol, CSF 3β-hydroxycholest-5-en-(25R)26-oic acid (3β-HCA), CSF 24S-HC and CSF 8-DHC. The metabolites CSF 25-HC, CSF 7αH,3O-CA, plasma 8(14)-dehydrocholesterol (8(14)-DHC), CSF 7β-HC, plasma 3β,22-diHCA, plasma 3β,7β-diHCA, and plasma 3β-HCA also ranked highly but less so than the ones above. The highest-ranking metabolite overall is plasma 24S-HC. CSF 24S-HC also ranks highly.

### Introduction, Method, Results and Discussion

#### Introduction

Several techniques are available for the classification of individuals into two alternative classes based on values for a set of independent variables (features). These techniques could have two distinct objectives. One is to identify a small subset of the independent variables which are the most useful for classification, ignoring the rest. This subset could represent targets for downstream analysis, for example in work to identify therapeutic targets. Another objective is to use all available variables to maximise the ability of the statistical model to correctly assign individuals to the correct classes. This could be useful in a clinical context. If many variables do not provide information, then classification with a smaller subset of variables would be preferable.

Stepwise Logistic Regression has been widely used in the past to identify a small subset of the most useful and significant variables from those available. It relates the logarithm of the odds of individuals belonging to the two classes to the values of the independent variables. In forward regression, variables are added sequentially to the model until no further significant variables can be added. In backward regression all variables are initially in the model and removed one at a time until only significant variables remain.

For a significant variable retained in the model, this significance is assessed when holding all other variables at a constant value. Thus, the technique gives the unique contribution of the significant variable adjusted for and controlling for the influence of the other variables. Stepwise logistic regression does have several disadvantages. There is a tendency towards overfitting where the model tends to fit patterns specific to the data at hand (including effects of random noise), and hence might not fit new unseen data well. The technique is sensitive to non-linearity and interactions between variables which might require additional terms in the regression equation. Also, the technique is sensitive to correlations between the independent variables (multicollinearity). Stepwise regression will work well when there is no non-linearity and complex interactions between the variables are absent, and correlations between the variables are not high.

Regularisation techniques (Lasso, Ridge and Elastic Net) help mitigate the effect of multicollinearity and overfitting by penalising large regression coefficients. In Lasso, the magnitude of the penalty is proportional to the absolute values of the regression coefficients. This results in some regression coefficients being shrunk to zero and being eliminated from the model. This produces a simpler less complex model useful for feature selection. In Ridge, the magnitude of the penalty is proportional to the squares of the regression coefficients. This also shrinks regression coefficients but not to zero, so all variables are retained in the model. This is useful for classification assuming all variables contribute some small amount of information to the classification but is less useful for feature selection as all independent variables are retained. Elastic net balances the penalties of Lasso and Ridge and might handle multicollinearity better than lasso or ridge alone.

The machine learning techniques Random Forest and Gradient Boosting use decision trees to predict class membership. Random Forest is an ensemble technique which creates a consensus model derived from a large sample of trees each built from a different random training sample of the data, each sample being taken with replacement as in bootstrapping. The individuals excluded from the sample, called the out of bag (OOB) testing sample, can be used to assess the classification utility of the tree obtained from the corresponding training sample. In addition, random samples of variables are used to split groups of individuals at nodes in the tree. Unlike Random Forest, in Gradient Boosting a succession of trees are built sequentially each attempting to correct errors in prior trees to arrive at a final model.

Random forest produces a model which is usually robust to overfitting because the predictions derive from the averaging of many trees, with the impact of trees that overfit the data being reduced by trees that do not overfit. It can take account of non-linear relationships between dependent and independent variables and interactions between the independent variables, whereas in Stepwise Logistic Regression these might need to be modelled. Random Forest is also robust to multicollinearity, and the presence of outliers which in Stepwise Logistic Regression require modification of the dataset. Gradient Boosting generally shares the benefits of Random Forest but needs to be tuned carefully to avoid overfitting because error in earlier trees might propagate through later trees. In both the techniques more important and influential independent variables show a greater decline in impurity when nodes are split. Impurity reflects the difference in frequency of the two classes between the subnodes, the bigger the difference the greater the decline in impurity from the parent node.

Both the regression techniques (Stepwise Logistic Regression, Lasso, Ridge, and Elastic Net) and the machine learning techniques (Random Forest and Gradient Boosting) provide an estimate of the importance of each variable for prediction in the model. In regression techniques the contribution of individual variables might be gauged clearly from the absolute magnitude of the regression coefficients. In the machine learning techniques importance scores are based on how often an independent variable is used for splitting trees and the effectiveness of partitioning groups of individuals at nodes in the trees. Ridge, Random Forest, and Gradient Boosting include all variables in the final model which is useful for classification. Stepwise Logistic Regression, Lasso and Elastic Net produce a shortened list of only the most important variables which is more useful for feature selection.

#### Method

Pairwise comparisons were made between three groups of individuals, Control (N=81), PreManifest (N=124) and Manifest (N=183). There are 55 independent variables. Of these, 51 are the measured levels of metabolites in either blood plasma or CSF as well as Age, BMI, Sex (M or F) or taking Statins (Yes or No). The latter two are categorical variables, all the others treated as continuous. In the output the prefixes c and p are added to the variables names to indicate their measurement in CSF and plasma respectively.

For analysis carried out in R, seven classification techniques were compared. These are five regression techniques, Forward Stepwise Logistic Regression, Backwards Stepwise Logistic Regression, and the regularisation techniques Lasso, Ridge and Elastic Net, and two machine learning techniques Random Forest and Gradient Boosting. For the regression techniques variables values were standardized to mean=0 and variance =1, so that variables could be ranked and compared according to the absolute value of the regression coefficients. None of the seven techniques require that the variables are normally distributed. The R packages MASS, pRoc, performance, glmnet, randomForest, gbm, caret and RobustRankAggreg were used in the analyses.

For Stepwise Logistic Regression the favoured model was that having the lowest value for the Akaike information criterion (AIC). The AIC is calculated as AIC = k x (number of parameters) – 2 ln(L) where L is the likelihood of the model. By default, AIC uses a penalty of k=2. However, in the current analyses k=4 was used. This resulted in the number of parameters (independent variables) retained in the model being in line with the commonly used heuristic that this number be roughly equal to the smallest sample size divided by 10 or to its square root. For the favoured model, p-values were also calculated for the retained variables. The predictions of probability of group membership were made using the regression equations for the retained model. For the current data there is satisfactory concordance with the assumptions of Stepwise Logistic Regression[1]. Meeting these assumptions is of less importance for the other techniques described below.

Regularisation is achieved with two parameters, alpha which determines the balance between Lasso and Ridge, and lambda, which determines the strength of the regularisation. If alpha = 1, the technique is Lasso. If alpha = 0, the technique is Ridge. In both cases, lambda was tuned to an optimal value using 10-fold cross-validation, with the optimal value being determined by minimizing the cross-validated penalized deviance, which is a function of the likelihood of the model and the alpha and tuned lambda values. For Elastic Net, alpha was also tuned using a grid search method, testing alpha values from 0.1 to 0.9 in increments of 0.2. The optimal combination of alpha and lambda was chosen based on the highest 10-fold cross-validated area under the ROC curve (AUC). The predictions of probability of group membership were made using the regression equation for the best model, based on the optimal values of alpha and lambda obtained from the tuning process.

To optimize the random forest model, tuning was used to determine the best value for the mtry parameter, which specifies the number of variables randomly sampled as candidates at each node split when building the trees. The tuning process was carried out with the following settings: number of trees (ntreeTry) = 500 trees were used in each trial of the tuning process, step factor (stepFactor) = 1.5 which means that the mtry value was multiplied by 1.5 in each step to get the next candidate value, improvement threshold (improve) = 0.01, indicating that the tuning process continued as long as the relative improvement in out of bag (OOB) error was at least 1%. The tuning process identified the optimal value for mtry which was used in the final model with 500 decision trees. To assess the importance and contribution of each independent variable (feature) in the final random forest model, the Gini coefficient measure of impurity reduction, was used. The greater the difference between the two groups of individuals in the sub nodes descended from a node the better the separation and the greater the reduction in Gini impurity.

To optimize the gradient boosting model, tuning with grid search was employed to determine the best combination of four key parameters: the number of trees (n.trees, 500, 1000, 1500), the minimum number of observations in a terminal node (minobs, 5, 10, 15), the shrinkage rate which controls the learning rate (shrink, 0.01, 0.001), and the interaction depth which controls the complexity of the trees (depth, 1, 2, 3). For each combination of the above parameters values a gradient boosting model was fitted with a Bernoulli distribution for binary classification and five-fold cross-validation to assess performance. The combination of parameters that resulted in the lowest cross-validated error was selected as the optimal set, and the best model stored for making predictions on the data. The relative influence of each independent variable in this final model was estimated by the decrease in the Gini coefficient, across all trees in the ensemble, the more influential variables showing larger overall decreases. The tuning process also outputs the best iteration in which the model achieves its lowest cross-validated error. The values of the parameters obtained in tuning for all techniques are given in the Appendix at the end of this section. The seven classification techniques described above were carried out on the full dataset for each of the three comparisons to develop classification models and for feature selection.

In addition, five-fold cross-validation was used as a final step for each of the 7 (techniques) x 3 (comparisons). In this approach the data was allocated randomly to five testing samples (test1 to test5) each consisting of a different 20% of the data. In each fold the remaining 80% of the data was used as a training sample (train1 to train5) in which models were developed for the seven classification techniques in the same way as for the full dataset. The models were then used to predict class membership on the five testing samples, that is the model from train1 was tested on test1 and so on. The same random seeds were used for all classification techniques when constructing the individuals assigned to the 5 folds. Thus, the cross-validation was carried out on identical datasets for each classification technique which is good for comparative purposes. However, three different seeds were used for each comparison (123 for PreManifest and Manifest, 112 for Control and PreManifest, and 113 for Control and Manifest).

The performance of the models on the full dataset and on the test datasets in the cross-validation were assessed using standard methods. This included the ROC curve and AUC. All seven techniques allow prediction of group membership for individual subjects. Confusion tables (2x2) were constructed from the predicted probability of group membership using the prevalence method whereby the probability cutoff ensured that the numbers predicted for the two groups in the confusion table matched actual numbers observed. Statistics deriving from the confusion tables including sensitivity and specificity were calculated.

A permutation analysis was also carried out on the full dataset for each of the classification techniques for each of the three comparisons by shuffling the labels (Control, PreManifest and Manifest) across individuals. One thousand permuted samples were used for each permutation analysis again using the same random seed for each classification technique. This allowed computation of p-values for the individual features which are not provided by the regularisation and machine learning techniques. It also allowed computation of 95% confidence intervals for AUC based on quantiles against which the observed value for the full dataset could be compared.

The lists of the ranked independent variables retained in the different classification techniques were compared to determine whether there was a consensus on which variables occur consistently at high positions in the different lists (Supplementary_Workbook_2). This was carried out using the Stuart aggregation method which minimises disagreement between lists and gives more weight to higher ranked items. The method returns a p-value score for each variable in the list, those with lower p-values being consistently higher in ranking across all the lists. The different classification techniques produce lists of different lengths. Thus ridge, random forest and gradient boosting include all independent variables, the other techniques have lists of reduced length only for variables which are considered important or influential. The aggregation method can deal with this situation but clearly cannot use information from variables that are missing from a particular list. Most of the variables identified as important are included in all lists.

#### Results and Discussion

Details of the analyses are given in Supplementary_Workbook_1 (available to view on figshare https://doi.org/10.6084/m9.figshare.30510515) where there is a separate worksheet for each of the 7 (techniques) x 3 (comparisons) analyses in identical format.

Each of the results worksheets gives information on the analysis on the full data set. This comprises: (1) Significance levels or importance levels for the individual independent variables retained in the model including permutation p-values. (2) A ROC curve with associated statistics including area under the ROC curve (AUC). (3) A predicted probability histogram of membership of the two groups in the comparison. (4) A 2x2 confusion table showing actual versus predicted group membership. (5) A histogram of AUC from permutation analysis with 95% confidence intervals and observed AUC value. (6) The results of the five-fold cross validation analysis with ROC curves and associated statistics.

Summary tables are given in Supplementary_Workbook_2 (available to view on figshare https://doi.org/10.6084/m9.figshare.30510515). Worksheet AUC_2x2 section (A) show that AUC is consistently high for the PreManifest and Manifest comparison supported in worksheet AUC_2x2 (B) by many significant values for the 2x2 confusion tables. This is observed for the full dataset and the five test samples from cross-validation. This gives good confidence of a real difference between these two groups. For the other two group comparisons involving Control, the full dataset also gives high values of AUC and significant values for the confusion table. However, the values for the test samples of the cross validation are depressed with many non-significant p-values.

Successful classification on the full data set is expected because the model is developed on this same dataset. It might in part be due to capturing the many patterns in a larger sample and in part due to overfitting. Conversely the cross-validation test samples are unseen data for each fold and are smaller. This limits the power of the models developed on the training samples to capture patterns in the data and reduces the chance of statistically significant p-values in the test sample confusion tables. Thus, interpretation of the good classification results for the full data set needs to be balanced against poorer results for the smaller cross-validation test samples. One reflection of this difference is that the AUC values for the full sample are almost always greater than the mean value over folds in worksheet AUC_2x2 section (A).

The values of AUC for forwards and backwards logistic regression are similar to those obtained for the regularisation techniques (lasso, ridge and elastic net). The lambda values for the regularisation techniques are also low, mainly less than 0.1 (Appendix). This is consistent with all these techniques fitting the data well without the need for strong regularisation to avoid overfitting, and that substantial multicollinearity does not exist in the data. Although it is useful to test and compare different classification techniques, the results for the current data indicate that linear models such as logistic regression may perform adequately in classification and in the selection of the most important independent variables (features) for further studies.

For the machine learning techniques, gradient boosting produced the highest AUC values for the full dataset of all the classification techniques but does not seem to have performed better in the cross-validation. Random Forest gave the lowest AUC values for the full dataset. These AUC values were similar in magnitude to the averages across folds. This may be the result of this method being trained on multiple random subsets of the data producing a robust result free of overfitting at the cost of a reduced AUC.

The significance values (estimated p-value) for AUC in the permutation analysis are given in worksheet AUC_2x2 section (A). A value of 0.000 indicates that none of the 1000 permutation samples gave an AUC value higher than that for the full dataset. The values are uniformly very low for all the techniques except for gradient boosting and random forest for the two comparisons with control. This hints that these techniques might be relatively free of overfitting compared with the others. The permutation results for AUC are also given in histograms on the individual worksheets (Supplementary_workbook_1). The AUC values for the permuted data tend to be shifted above 0.5, the fitting of the model being unlikely to generate either a value lower than 0.5 or higher values for the off-diagonal in the 2x2 confusion tables. These results suggest that high values of AUC must be interpreted with caution without either permutation analysis or the five-fold cross validation.

A model can be said to be stable if it produces similar results when tested on multiple samples of unseen data. This can be examined by looking at the variation (standard deviation) of the performance metric AUC across the five folds in cross-validation. These values are given in worksheet AUC_2x2 section (A). The overall mean value of the standard deviation across all methods is lower for the comparison of PreManifest and Manifest than for the other two comparisons. This is an indication that for this comparison real differences between the two groups are being captured by the models on the training samples which can be extrapolated to the unseen test samples for each fold. There are not any marked easily interpretable differences between the seven classification techniques. However standard deviation values for logistic regression are as low as if not lower than for the other classification techniques, an indication of comparable stability.

Stepwise regression provides significance levels as p-values for the independent variables that are retained by the model. The other classification techniques provide relative measures of importance.

The permutation analysis provides p-values for all the techniques which supplements the importance values and allows cross-comparison of all the classification techniques (Supplementary_Workbook_1). (Backward stepwise logistic regression was excluded from the permutation analysis because of excessive computational time requirements, given the available computing resources.) For forward stepwise regression the permutation p-values, with a few exceptions, match or exceed the significance of those p-values obtained from the regression analysis on the full dataset. For the regularisation regression techniques (lasso, ridge and elastic net) most of the variables judged to be of greatest importance are also statistically significant in the permutation test. This might be expected and is reassuring. Similar results are obtained for the machine learning techniques random forest and gradient boosting, except that the number of significant variables is reduced particularly for gradient boosting. This could be because the regression techniques are more prone to overfitting thus more easily generating significant p-values. This underlines the importance of paying attention to the final five-fold cross validation analysis. Variables which are not significant should however not be disregarded. Particularly in ridge, random forest and gradient boosting these variables may possess information which contributes to the utility of the classification model.

The ranked lists of independent variables retained by the seven different classification techniques for the full data set are given in Supplementary_Workbook_2 for the three binary group comparisons (worksheets PreMan, ConPre, and ConMan). A consensus ranking over the seven lists is given in section (B). The associated p-value relates to whether a variable is higher in all the seven lists than would be expected by chance rather than to the importance of the variable in the classification models. The top variables in the consensus ranking are coded with colour fill, and these colours mapped back onto the lists for the seven techniques to facilitate comparison between them.

Several key points might be made. For each of the three pairwise comparisons between groups there is overlap between the classification techniques in the variables that rank at the top of the seven lists. This is not surprising as the different classification techniques are expected to capture some similar patterns in the data. The different classification techniques also show some differences in the ranking of the independent variables consistent with their operating on the data in different ways. This highlights the utility of testing different approaches and not relying on a single approach. Particularly for the variables that are high but not at the top of the lists, the machine learning techniques (Random Forest and Gradient Boosting) produce rankings which are somewhat different from the other techniques. On the other hand, the basic regression techniques give results which are not so different from the others overall. This suggests that they may be effective for this dataset in identifying the most important variables in relation to classification and feature selection.

The consensus list over the seven techniques for each of the three pairwise comparisons between groups are given and aligned side by side in worksheet ClassListsCompared (A). An overall consensus of these is provided in (B). Colour coding in (B) is mapped back to the three individual lists in (A) to facilitate comparison between them. The lists have both overlaps and differences between them. It is hoped that the current analysis using seven different classification techniques underlines that different methods can lead to deeper understanding of patterns in the data.

The three lists for the three pairwise comparisons are compared in worksheet ClassListsCompared (C) for the top 16 variables in each list. This number of 16 is a little greater than the number of significant variables in the regression techniques but corresponds with the number of variables significant in the consensus obtained with the Stuart aggregation method. There is overlap in the variables at the top of the three lists. However, the comparison between Control and PreManifest appears different in relation to the important variables, compared with the other two comparisons which resemble each other more closely. This is particularly evident in the variables near the top of the list (ClassListsCompared (C)). The two more similar comparisons reflect a contrast between Manifest and the other two groups, leaving a somewhat different selection of variables highlighted as important in the comparison of Control and PreManifest. This pattern of variation might reflect qualitative differences between the three groups which cannot easily be captured as a linear trend from Control to PreManifest to Manifest. Two variables pCholesterol and Age are highlighted as important as in all three comparisons between groups. These might be the best candidates for showing an additive linear trend between the groups.

Focusing on the overall consensus (ClassListsCompared (B)) and on the comparisons with Manifest (ClassListsCompared (C)) allows identification of the oxysterols that might be important generally for classification and feature selection. In the overall consensus list the first set comprise p24SHC (plasma 24S-HC), p7Dehydrocholesterol (plasma 7-DHC), pCholesterol (plasma cholesterol), c3βHCA (CSF 3β-HCA), c24SHC (CSF 24S-HC) and c89Dehydrocholesterol (CSF 8-DHC), in order of importance. The second set comprises c25HC (CSF 25-HC), c7αH3OCA18 (CSF 7αH,3O-CA), p814Dehydrocholesterol (plasma 8(14)-DHC), c7βHC (CSF 7β-HC), p3β22diHCA (plasma 3β,22-diHCA), p3β7βdiHCA (plasma 3β,7β-diHCA), and p3βHCA (plasma 3β-HCA). Of the individual variables that are not metabolites, Age and BMI stand out at the top of the overall consensus list as being influential. Of all the oxysterols 24S-HC might be identified as the most important which fits with a priori expectations.

#### Conclusion

The binary classification techniques investigated provide a means of both effectively classifying HD subjects and identifying the most important independent variables (features) to be used for classification. Regression techniques including regularisation and machine learning show both similarities and differences in relation to the variables important in the classifications and may complement each other. The variables important in the comparisons between Manifest and Control and between Manifest and PreManifest are more similar to each other than to the comparison of PreManifest and Control. The binary classification of individual subjects into PreManifest and Manifest groups appears to have been the most successful. Of all the oxysterols implicated as potentially good classifiers the 24S-HC stands out as potentially being the most important.

### Appendix – Values of tuned parameters

Lasso

Optimal lambda

PreManifest and Manifest: 0.02457366

Control and PreManifest: 0.02717733

Control and Manifest: 0.0191604

Ridge

Optimal lambda

PreManifest and Manifest: 0.05294235

Control and PreManifest: 0.1352622

Control and Manifest: 0.1046594

Elastic Net

Best alpha

PreManifest and Manifest: 0.7

Control and PreManifest: 0.5

Control and Manifest: 0.5

Best lambda

PreManifest and Manifest: 0.02914498

Control and PreManifest: 0.05965418

Control and Manifest: 0.03181459

Random Forest

Optimal mtry

PreManifest and Manifest: 7

Control and PreManifest: 7

Control and Manifest: 7

Gradient Boosting

PreManifest and Manifest

n.trees: 1000

minobs: 15

shrink: 0.01

depth: 1

best iteration: 651

Control and PreManifest

n.trees: 500

minobs: 15

shrink: 0.01

depth:1

best iteration: 344

Control and Manifest

n.trees: 500

minobs: 15

shrink: 0.01

depth: 3

best iteration: 228
